# WHOLE GENOME SEQUENCING ANALYSIS OF BODY MASS INDEX IDENTIFIES NOVEL AFRICAN ANCESTRY-SPECIFIC RISK ALLELE

**DOI:** 10.1101/2023.08.21.23293271

**Authors:** Xinruo Zhang, Jennifer A. Brody, Mariaelisa Graff, Heather M. Highland, Nathalie Chami, Hanfei Xu, Zhe Wang, Kendra Ferrier, Geetha Chittoor, Navya S. Josyula, Xihao Li, Zilin Li, Matthew A. Allison, Diane M. Becker, Lawrence F. Bielak, Joshua C. Bis, Meher Preethi Boorgula, Donald W. Bowden, Jai G. Broome, Erin J. Buth, Christopher S. Carlson, Kyong-Mi Chang, Sameer Chavan, Yen-Feng Chiu, Lee-Ming Chuang, Matthew P. Conomos, Dawn L. DeMeo, Margaret Du, Ravindranath Duggirala, Celeste Eng, Alison E. Fohner, Barry I. Freedman, Melanie E. Garrett, Xiuqing Guo, Chris Haiman, Benjamin D. Heavner, Bertha Hidalgo, James E. Hixson, Yuk-Lam Ho, Brian D. Hobbs, Donglei Hu, Qin Hui, Chii-Min Hwu, Rebecca D. Jackson, Deepti Jain, Rita R. Kalyani, Sharon L.R. Kardia, Tanika N. Kelly, Ethan M. Lange, Michael LeNoir, Changwei Li, Loic Le. Marchand, Merry-Lynn N. McDonald, Caitlin P. McHugh, Alanna C. Morrison, Take Naseri, NHLBI Trans-Omics for Precision Medicine (TOPMed) Consortium, Jeffrey O’Connell, Christopher J. O’Donnell, Nicholette D. Palmer, James S. Pankow, James A. Perry, Ulrike Peters, Michael H. Preuss, D.C. Rao, Elizabeth A. Regan, Sefuiva M. Reupena, Dan M. Roden, Jose Rodriguez-Santana, Colleen M. Sitlani, Jennifer A. Smith, Hemant K. Tiwari, Ramachandran S. Vasan, Zeyuan Wang, Daniel E. Weeks, Jennifer Wessel, Kerri L. Wiggins, Lynne R. Wilkens, Peter W.F. Wilson, Lisa R. Yanek, Zachary T. Yoneda, Wei Zhao, Sebastian Zöllner, Donna K. Arnett, Allison E. Ashley-Koch, Kathleen C. Barnes, John Blangero, Eric Boerwinkle, Esteban G. Burchard, April P. Carson, Daniel I. Chasman, Yii-Der Ida Chen, Joanne E. Curran, Myriam Fornage, Victor R. Gordeuk, Jiang He, Susan R. Heckbert, Lifang Hou, Marguerite R. Irvin, Charles Kooperberg, Ryan L. Minster, Braxton D. Mitchell, Mehdi Nouraie, Bruce M. Psaty, Laura M. Raffield, Alexander P. Reiner, Stephen S. Rich, Jerome I. Rotter, M. Benjamin Shoemaker, Nicholas L. Smith, Kent D. Taylor, Marilyn J. Telen, Scott T. Weiss, Yingze Zhang, Nancy Heard- Costa, Yan V. Sun, Xihong Lin, L. Adrienne Cupples, Leslie A. Lange, Ching-Ti Liu, Ruth J.F. Loos, Kari E. North, Anne E. Justice

## Abstract

Obesity is a major public health crisis associated with high mortality rates. Previous genome-wide association studies (GWAS) investigating body mass index (BMI) have largely relied on imputed data from European individuals. This study leveraged whole-genome sequencing (WGS) data from 88,873 participants from the Trans-Omics for Precision Medicine (TOPMed) Program, of which 51% were of non-European population groups. We discovered 18 BMI-associated signals (*P* < 5 × 10^-9^). Notably, we identified and replicated a novel low frequency single nucleotide polymorphism (SNP) in *MTMR3* that was common in individuals of African descent. Using a diverse study population, we further identified two novel secondary signals in known BMI loci and pinpointed two likely causal variants in the *POC5* and *DMD* loci. Our work demonstrates the benefits of combining WGS and diverse cohorts in expanding current catalog of variants and genes confer risk for obesity, bringing us one step closer to personalized medicine.

## INTRODUCTION

In 2015, approximately 12% of adults worldwide had obesity ^1^, and four years later, the global obesity-related deaths amounted to five million, translating to an age-standardized mortality rate of 62.6 per 100,000 individuals in 2019 ^2^. Previous genome-wide association studies (GWAS) have identified hundreds of loci associated with obesity-related traits, primarily with body mass index (BMI) – a practical and widely used proxy of overall adiposity. However, most of these genome-wide screens relied on meta-analyses of imputed data, predominantly from individuals of European ancestry ^3,4^.

Despite making some advancements toward improving ancestral diversity in GWAS, ancestry-stratified analyses and multi-ancestry analyses leveraged for discovery and fine-mapping are uncommon and largely underpowered by comparison. Furthermore, follow-up investigations for known BMI loci identified in European ancestry populations are insufficiently conducted to evaluate the generalizability of these loci. As such, the majority of BMI risk variants are common variants (minor allele frequency [MAF] > 5%) in primarily European ancestry populations, most of which exhibit small effect sizes. While these index variants collectively explain less than 5% of the total phenotypic variation in BMI ^5^, it is estimated that as much as 1/5 of the phenotypic variance can be captured by common variants across the entire genome ^5^, leaving low and rare variants (MAF ≤ 5%) with potentially large effects to be explored ^6^.

Whole-genome sequencing (WGS) outperforms genotyping arrays in capturing low and rare frequency variants, as demonstrated in a recent study where researchers revealed that the heritability of BMI estimated using WGS data was comparable to the pedigree-based estimates, h^2^ ≈ 0.40 ^7^. Thus, the discrepancy between phenotypic variance explained by genetic variations in GWAS compared to the expected heritability may be due to the use of imputed genotypes rather than directly sequenced variations. Causal variants or SNPs in known loci may not be represented on 1000 Genomes panels, or not well imputed from reference data because of differences in linkage disequilibrium (LD) across populations. To address this limitation, we conducted WGS association analyses to identify rare, low-frequency, and ancestry-specific genetic variants associated with BMI, using data from the Trans-Omics for Precision Medicine (TOPMed) Program ^8^, which is the most racially and ethnically diverse WGS program to date, as well as the Centers for Common Disease Genomics (CCDG) Program ^9^.

## METHODS

### Study Population and Phenotype

Our study population was racially, ethnically, geographically, and ancestrally diverse. We analyzed a multi-population sample of 88,873 adults from 36 studies in the freeze 8 TOPMed and CCDG programs (**Figure 1, Supplementary Data 1)**. They belonged to 15 population groups, reflecting the way participants self-identified in each study. For individuals who had unreported or non-specific population memberships (e.g., “Multiple” or “Other”), we applied the Harmonized Ancestry and Race/Ethnicity (HARE) method ^10^ to infer their group memberships using genetic data. This imputation was applied to 8,015 participants (9% of the overall population), assigning each to one of the existing population groups. In this way, our study population groups were defined based on a combination of self-reported identity and the first nine genetic principal components (PCs) **(Figure 1, Supplementary Fig 1**, and **Supplementary Data 1)**.

**Figure 1.**
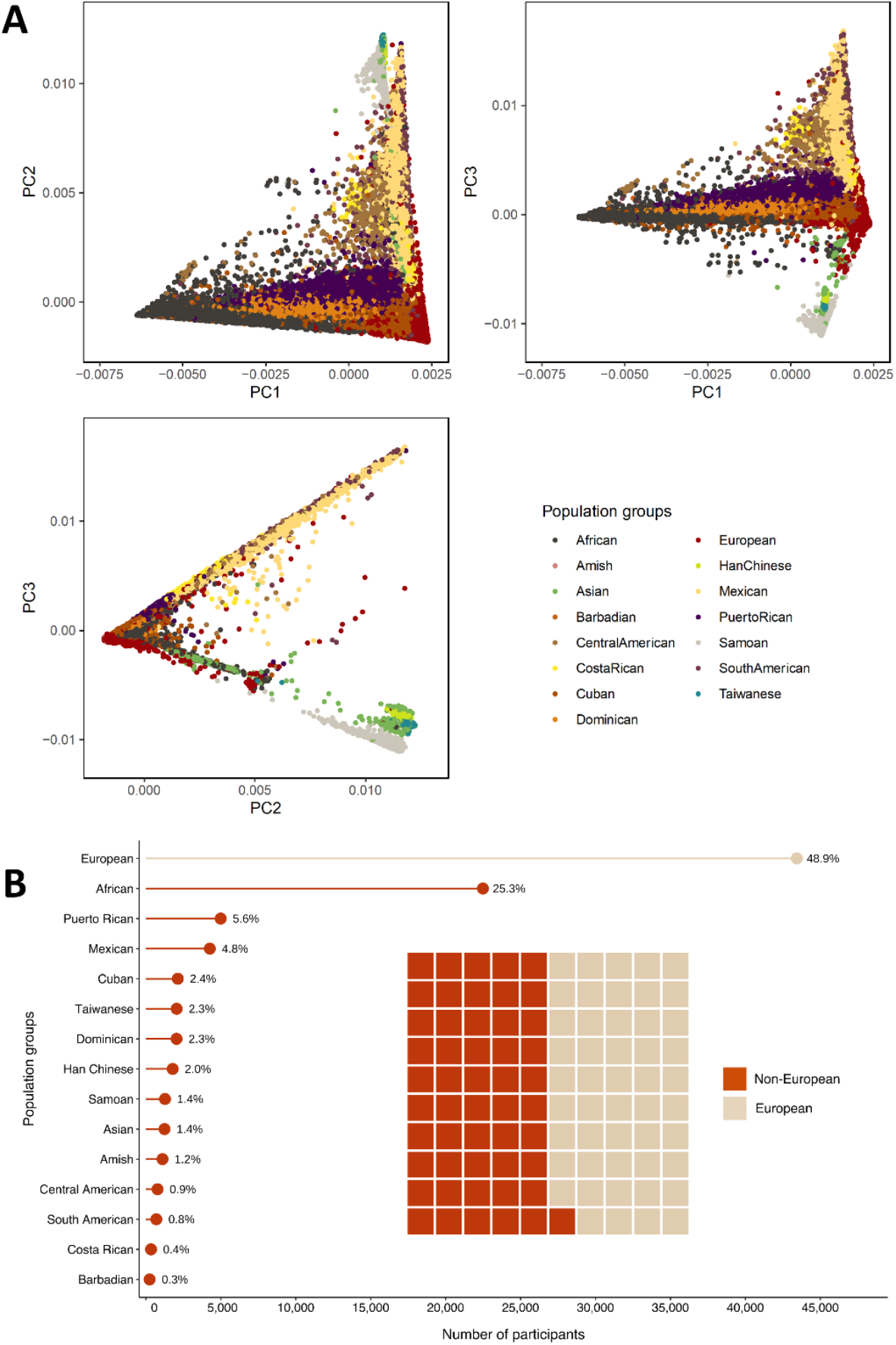
Study population group composition. A) Pairwise scatter plots of the first three principal components (PCs) by population group. B) This image contains a lollipop chart and a waffle plot illustrating the number and proportion of participants by population group. Our study population was composed of 88,873 participants from 15 population groups, 51% of which are non-European.

The 15 population groups were labeled by their self-identified or primary inferred population group (e.g., predominantly African ancestry/admixed African/Black were labeled as “African”). Sample sizes for these groups ranged from 341 to over 43,000 as follows: African (N = 22,488), Amish (N = 1,106), Asian (N = 1,241), Barbadian (N = 248), Central American (N = 776), Costa Rican (N = 341), Cuban (N = 2,128), Dominican (N = 2,046), European (N = 43,434), Han Chinese (N = 1,787), Mexican (N = 4,265), Puerto Rican (N = 4,991), Samoan (N = 1,274), South American (N = 695), and Taiwanese (N = 2,053). We refer to analyses involving all 15 population groups as multi-population analysis and group-specific analyses by their primary population group.

Among the 88,873 participants, 53,109 (60%) were female and 45,439 (51%) were non-European. The mean (SD) age of the participants was 53.5 (15.1) years. Additional descriptive tables of the participants are presented in **Supplementary Data 2 – 4**. BMI was calculated by dividing weight in kilograms by the square of height in meters. Participants were excluded from analyses if less than 18 years of age, had known pregnancy at the time of BMI measurement, had implausible BMI values (above 100 kg/m^2^ without corroborating evidence), or did not provide appropriate consent. The mean (SD) of BMI varied by study, ranging from 23.4 (3.1) in GenSALT to 32.7 (6.8) in VAFAR (**Supplementary Data 2**), and by population group, ranging from 23.4 (3.1) in Han Chinese to 33.7 (6.8) in Samoans (**Supplementary Data 3**).

### TOPMed WGS

A detailed description of WGS methods has been reported previously ^11^. Details regarding the laboratory methods, data processing, and quality control are described on the TOPMed website (https://www.nhlbiwgs.org/methods). Briefly, ∼30X WGS was conducted using Illumina HiSeq X Ten instruments at six sequencing centers. At the Center for Statistical Genetics at University of Michigan, TOPMed sequence data were mapped to the GRCh38 human genome reference sequence in a manner consistent with the joint CCDG/TOPMed functionally equivalent read mapping pipeline ^12^. Joint genotype calling on all samples in Freeze 8 used the GotCloud pipeline ^13^. Variants were filtered using a Support Vector Machine (SVM) implemented in the libsvm software package. Sample-level quality assurance steps included concordance between annotated and genetic sex, between prior SNP array genotyping and WGS-derived genotypes, and between observed and expected relatedness and pedigree information from cleaned sequence data.

### Common Variant Association Analysis

We performed multi-population WGS association analysis of BMI using GENESIS ^14^ on the Analysis Commons (http://analysiscommons.com) ^15^ computation platform. Analyses were performed using linear mixed models (LMM). To improve power and control for false positives with a non-normal phenotype distribution, we implemented a fully adjusted two-stage procedure for rank-normalization when fitting the null model ^16^. The first model was fit by adjusting BMI for age, age squared, sex, study, population group, ancestry-representative PCs generated using PC-AiR^17^, sequencing center, sequencing phase and project. A 4^th^ degree sparse empirical kinship matrix (KM) computed with PC-Relate ^18^ was included to account for genetic relatedness among participants. We also allowed for heterogeneous residual variances across sex by population group (e.g., female European), as it has previously been shown to improve control of genomic inflation ^19^. Residuals from the first model were rank normal transformed within population group and sex strata. The resulting transformed residuals were used to fit the second stage null model allowing for heterogeneous variances by the population group and sex strata and accounting for relatedness using the kinship matrix. Variants with a MAF of at least 0.5% were then tested individually.

Due to the large number of variants tested (N = 90,142,062) in the multi-population analysis, we adopted a significance threshold of 5 × 10^-9^ as has been used previously ^20^. Group-specific analyses were conducted in the two largest population groups, European and African.

### Replication Analyses

For the novel single-variant association identified in our discovery analyses, we requested replication from five independent cohorts of similar race, ethnicity, and continental ancestry to our discovery populations (N_total_ = 109,748): Multiethnic Cohort (MEC) ^21^, Million Veteran Program (MVP) ^22,23^, BioMe BioBank Program ^24^, UK Biobank (UKBB) ^25,26^, and the REasons for Geographic And Racial Differences in Stroke (REGARDS) study ^27^. Phenotypes were developed and analyses were carried out under the same protocol as outlined above. We subsequently conducted inverse variance weighted fixed effects meta-analysis in METAL^28^, using study-specific summary results. Additional details on the parent study design for each replication study are included in the **Supplementary Note**.

### Conditional Analysis

To identify loci harboring multiple independent signals, we performed stepwise conditional analyses on the most significant signal within 500kb of our index variant. The significance threshold for secondary signals was determined by Bonferroni correction for the number of variants across all regions tested, *P* = 5.96 × 10^-7^ (*P* < 0.05/83,928 SNPs with MAF > 0.5% within 500kb of the 16 index SNPs). Variants passing the significance threshold after the first round were further conditioned on the top variant in the locus after the first round of conditioning, to identify potential third signals within each locus using the same threshold.

To determine whether association signals in known loci were independent of known signals, we performed conditional analyses using previously published index variants ^5,29-48^. Specifically, we analyzed all genome-wide significant variants that were not the previously reported index variants but located within 500 kb of a known GWAS SNP. Given that these are potential new signals in regions known to influence BMI, index variants were considered independent if the estimated effect (β) value remained ≥ 90% of the unconditioned β value and *P* < 6.25 × 10^-3^ (0.05/8 loci tested). LDlink was used to calculate pairwise LD between potentially independent signals in known loci and produce LD heatmaps using the 1000 Genomes Global reference panel ^49^.

### Rare Variant Aggregate Association Analysis

Rare variants with a MAF ≤ 1% were tested in aggregate by gene unit across studies in the multi-population analysis. Variants were grouped into gene units in reference to GENCODE v28, including both coding variants and variants falling within gene-associated non-coding elements. Coding variants included high-confidence loss of function variants (Ensembl Variant Effect Predictor [VEP] LoF = HC), missense variants (MetaSVM score > 0) and in-frame insertion/deletions or synonymous variants (FATHNMM-XF coding score > 0.5). In addition to coding variants, we included variants falling within the promoter of each transcript tested. Promoter regions were defined as falling in the 5 kb region 5’ of the transcript and also overlaying a FANTOM5 Cage Peak ^50^. In order to identify regulatory elements likely to be acting through the tested gene, we leveraged the GeneHancer database ^51^. GeneHancer identifies enhancer regions and associates them with the specific genes they are likely to regulate, allowing us to aggregate regulatory regions by the likely target gene. GeneHancer regions were limited to the top 50% scored regions and variants falling in these regulatory elements were further filtered to those most likely to have a functional impact (FATHMM-MKL noncoding score > 0.75). Variants aggregated to gene units were tested using variant set mixed model association tests (SMMAT) ^52^. Variants were weighted inversely to their MAF using a beta distribution density function with parameters 1 and 25. Genes were considered significantly associated after Bonferroni correction for the number of genes analyzed (*P* < 5 × 10^-7^).

### Fine-Mapping

In order to identify candidate functional variants underlying association regions, we performed fine-mapping analyses in our multi-population GWAS single variant association summary statistics, using the program PAINTOR ^53^ which integrates the association strength and genomic functional annotation. We used the annotation file from aggregate-based testing described above under “Rare Variant Aggregate Association Analysis” to identify deleterious coding variants, variants within GeneHancer regions, and variants within gene promoter regions. We restricted this analysis to variants located within 100 kb of the locus index variants. We calculated LD using our analysis subset of the TOPMed data, assuming one causal variant per locus, unless evidence of independent secondary signals was identified, in which case we allowed for two causal variants per locus.

### PheWAS

To identify potential novel phenotypic associations with newly discovered variants, we performed a phenome-wide association (PheWAS) in the MyCode Community Health Initiative Study (MyCode), a hospital-based population study in central and northeastern Pennsylvania ^54^, and in the Charles Bronfman Institute for Personalized Medicine’s BioMe BioBank Program (BioMe) located in New York City^55^; both studies had genetic data linked to electronic health records (EHR). ICD-10-CM and ICD-9-CM codes were mapped to unique PheCodes using the Phecode Map v1.2 ^56^ from the EHR. Cases were defined if individuals had two or more PheCodes on separate dates, while controls had zero instances of the relevant PheCode. We performed association analyses on PheCodes with N ≥ 20 cases and 20 controls using logistic regressions, adjusting for current age, sex (for non-sex-specific PheCodes), and the first 15 PCs calculated from genome-wide data, and assuming an additive genetic model using the PheWAS package ^57^ in R. Analyses were conducted in the overall population as well as in individuals of genetically-informed African ancestry alone (as inferred from k-means clustering of the PCs ^58^), given the potential population-specific association of our novel locus. We restricted our analyses to unrelated individuals up to 2^nd^ degree. Association analyses were conducted within each study, followed by inverse variance weighted fixed effects meta-analysis in METAL^28^. PheCodes were deemed statistically significant after Bonferroni correction for the number of PheCodes analyzed (*P* < 0.05/538 = 9.3 × 10^-5^).

## RESULTS

### Single-variant Analyses

Among the 90 million SNPs included in the multi-population analysis, 86% (N = 77,178,487) were rare SNPs with a study-wide MAF of 0.5% < MAF ≤ 1%, and 6% (N = 5,542,150) were low-frequency (1% < MAF ≤ 5%) SNPs. In the multi-population unconditional analysis, we identified 16 loci that reached the prespecified genome-wide significance threshold of *P* < 5 × 10^-9^ (**Table 1, Figure 2, Supplementary Figs. 2 – 3**), including one low-frequency (MAF = 4%) and 15 common (MAF 14% – 50%) tag SNPs. In general, the low-frequency variant in our primary discovery showed a stronger effect than the common variants, with an estimated effect 2.14 times larger than the average common variants (0.078 *vs*. 0.037 on average). Of these 16 loci, 15 were in known BMI-associated regions, and one novel locus was identified on chromosome 22 harboring a low-frequency index SNP near *MTMR3* (rs111490516; MAF = 4%, β = 0.078, SE = 0.013, *P* = 4.52 × 10^-9^; **Table 1**). The MAF of this *MTMR3* locus varied widely across population groups, with the highest MAF observed in the African (13%) and Barbadian (13%) population groups, while it ranged from 0% to 5% in other population groups (**Supplementary Data 5**).

**Table 1.**
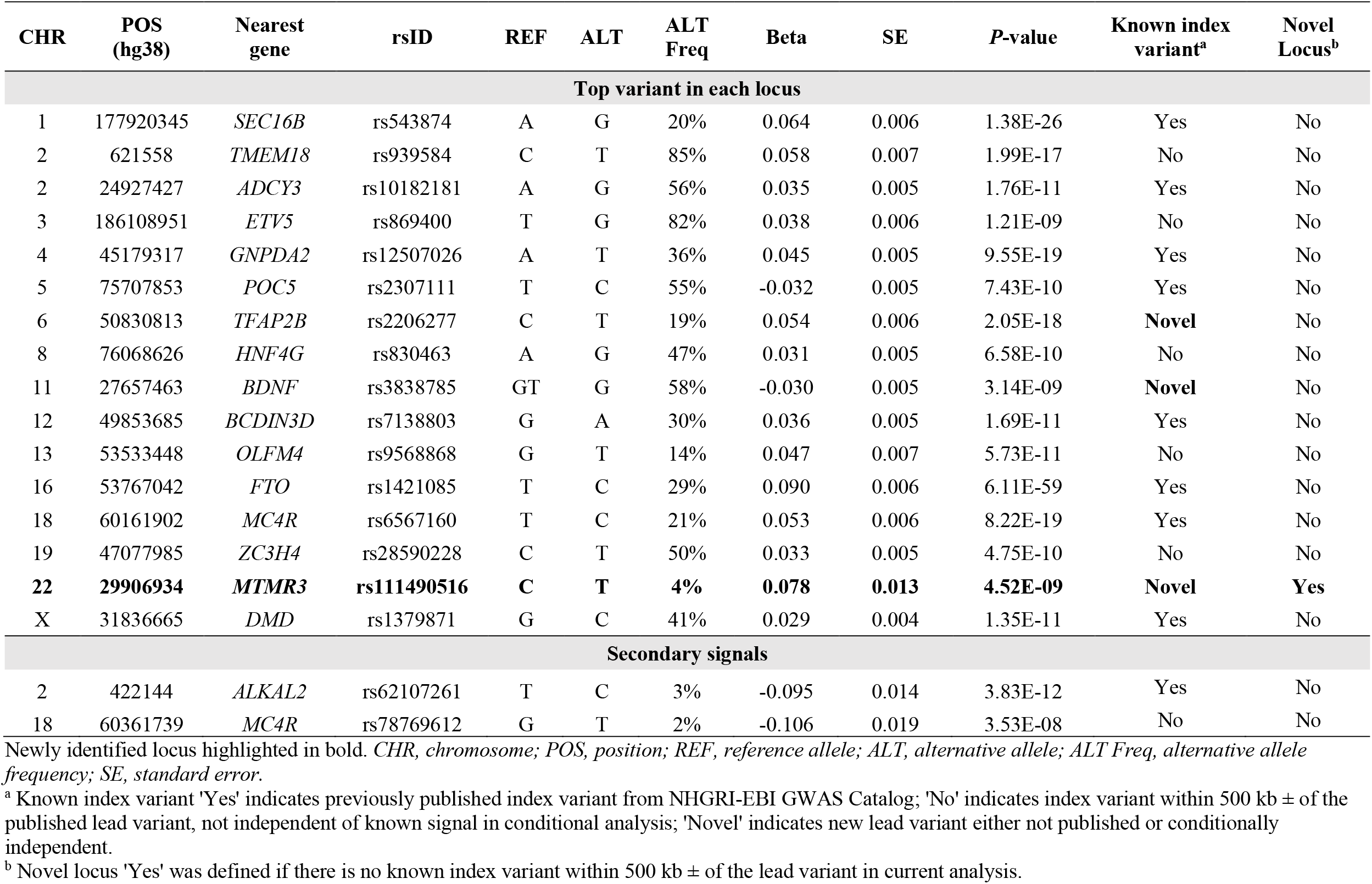
Summary of independent loci reaching genome-wide significance (*P* < 5 × 10^-9^) in single variant and internal conditional analyses.

| CHR | POS<br>(hg38) | Nearest<br>gene | rsID | REF | ALT | ALT<br>Freq | Beta | SE | P-value | Known index<br>variant<sup>a</sup> | Novel<br>Locus<sup>b</sup> |
| --- | --- | --- | --- | --- | --- | --- | --- | --- | --- | --- | --- |
| Top variant in each locus |  |  |  |  |  |  |  |  |  |  |  |
| 1 | 177920345 | *SEC16B* | rs543874 | A | G | 20% | 0.064 | 0.006 | 1.38E-26 | Yes | No |
| 2 | 621558 | *TMEM18* | rs939584 | C | T | 85% | 0.058 | 0.007 | 1.99E-17 | No | No |
| 2 | 24927427 | *ADCY3* | rs10182181 | A | G | 56% | 0.035 | 0.005 | 1.76E-11 | Yes | No |
| 3 | 186108951 | *ETV5* | rs869400 | T | G | 82% | 0.038 | 0.006 | 1.21E-09 | No | No |
| 4 | 45179317 | *GNPDA2* | rs12507026 | A | T | 36% | 0.045 | 0.005 | 9.55E-19 | Yes | No |
| 5 | 75707853 | *POC5* | rs2307111 | T | C | 55% | -0.032 | 0.005 | 7.43E-10 | Yes | No |
| 6 | 50830813 | *TFAP2B* | rs2206277 | C | T | 19% | 0.054 | 0.006 | 2.05E-18 | **Novel** | No |
| 8 | 76068626 | *HNF4G* | rs830463 | A | G | 47% | 0.031 | 0.005 | 6.58E-10 | No | No |
| 11 | 27657463 | *BDNF* | rs3838785 | GT | G | 58% | -0.030 | 0.005 | 3.14E-09 | **Novel** | No |
| 12 | 49853685 | *BCDIN3D* | rs7138803 | G | A | 30% | 0.036 | 0.005 | 1.69E-11 | Yes | No |
| 13 | 53533448 | *OLFM4* | rs9568868 | G | T | 14% | 0.047 | 0.007 | 5.73E-11 | No | No |
| 16 | 53767042 | *FTO* | rs1421085 | T | C | 29% | 0.090 | 0.006 | 6.11E-59 | Yes | No |
| 18 | 60161902 | *MC4R* | rs6567160 | T | C | 21% | 0.053 | 0.006 | 8.22E-19 | Yes | No |
| 19 | 47077985 | *ZC3H4* | rs28590228 | C | T | 50% | 0.033 | 0.005 | 4.75E-10 | No | No |
| **22** | **29906934** | ***MTMR3*** | **rs111490516** | **C** | **T** | **4%** | **0.078** | **0.013** | **4.52E-09** | **Novel** | **Yes** |
| X | 31836665 | *DMD* | rs1379871 | G | C | 41% | 0.029 | 0.004 | 1.35E-11 | Yes | No |
| Secondary signals |  |  |  |  |  |  |  |  |  |  |  |
| 2 | 422144 | *ALKAL2* | rs62107261 | T | C | 3% | -0.095 | 0.014 | 3.83E-12 | Yes | No |
| 18 | 60361739 | *MC4R* | rs78769612 | G | T | 2% | -0.106 | 0.019 | 3.53E-08 | No | No |

Newly identified locus highlighted in bold. CHR, chromosome; POS, position; REF, reference allele; ALT, alternative allele; ALT Freq, alternative allele frequency; SE, standard error.
<sup>a</sup> Known index variant 'Yes' indicates previously published index variant from NHGRI-EBI GWAS Catalog; 'No' indicates index variant within 500 kb ± of the published lead variant, not independent of known signal in conditional analysis; 'Novel' indicates new lead variant either not published or conditionally independent.
<sup>b</sup> Novel locus 'Yes' was defined if there is no known index variant within 500 kb ± of the lead variant in current analysis.

**Figure 2.**
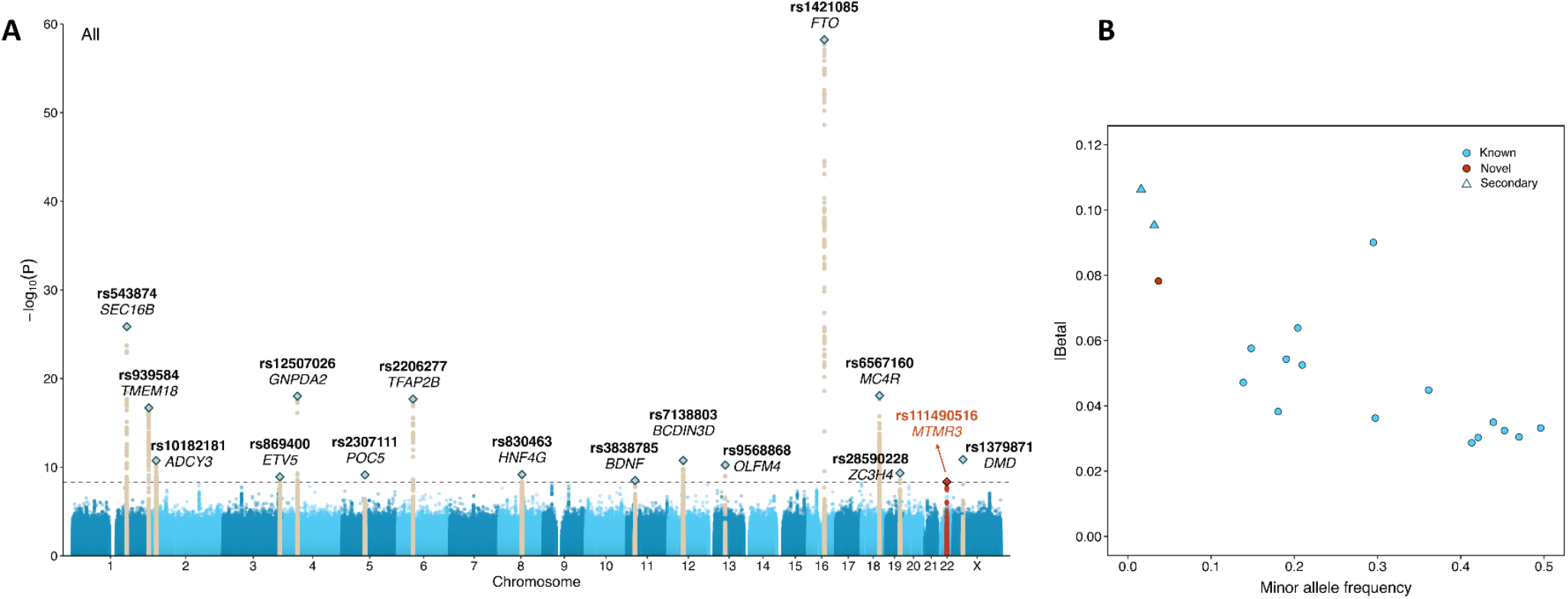
Summary of significant association findings. A) Manhattan plot of multi-population, single variant analysis (N = 88,873 individuals). The novel locus *(MTMR3)* is highlighted in red. Previously reported BMI loci are in dark beige. The horizontal dashed line indicates genome-wide significance threshold *P* = 5 × 10^-9^. B) Scatterplot showing the minor allele frequency compared to the absolute value of the estimate effect of the index variant at each significant locus. All effect estimates are from the primary analysis conducted across all population groups. Previously reported loci are highlighted in blue, while the novel locus is in red; circles represent the most significant variant at each locus, and triangles show newly reported secondary signals within known loci.

In the two population-specific analyses, 10 association signals reached genome-wide significance (**Supplementary Data 6, Supplementary Figs 4 – 7**). Two of these signals were also detected in the multi-population analysis. For two loci, *SEC16B* and *FTO*, each population-specific analysis revealed a distinct lead variant compared to the multi-population analysis; however, they were in high LD with (R^2^ = 0.95 and R^2^ = 1.00, according to TOP-LD ^59^; **Supplementary Data 6**) and within 30 kb of the multi-population lead SNPs. Notably, the novel locus in *MTMR3* achieved significance exclusively in the African group. While the most significant SNP in the African population group (rs73396827) differed from that in the multi-population analysis (rs111490516), the two were in strong LD in the TOPMed African population (R^2^ = 1.00). Both of these SNPs were fixed in the European group (MAF = 0%). In the European group analysis, one SNP in the *ALKAL2* locus on chromosome 2 (rs62107261, β = -0.102, SE = 0.016, *P* = 2.08 × 10^-10^, MAF = 5%) was not in LD with the corresponding lead variant in the multi-population analysis (R^2^ = 0.00, as calculated in the analysis subset), but was a known independent secondary signal at this locus ^40^. The remaining SNPs were in the proximity to the index SNPs in the corresponding loci from the multi-population analysis.

### Replication

The replication sample sizes ranged from 4,413 in BioMe to 79,889 in MVP (**Supplementary Data 7**). In the five replication studies of Blacks, Africans, and African Americans, the MAF of rs111490516 in *MTMR3* ranged from 11% to 13%, aligning with the 13% observed in our African and Barbadian groups and contrasting to the 0% to 5% range in our non-African discovery groups (**Supplementary Data 7**). We replicated the novel variant rs111490516, demonstrating directionally consistent associations with BMI across the replication studies and a 68% reduction in the estimated effect when meta-analyzing across replication studies (β = 0.025, SE = 0.007, *P* = 4.76 × 10^-4^, MAF = 11%) compared to the discovery analysis (**Figure 3, Supplementary Data 7**). In the meta-analysis of 198,621 individuals from both discovery and replication studies, the estimated effect was 0.037 with a SE of 0.006 and a *P*-value of 4.19 × 10^-9^ (**Figure 3, Supplementary Data 7**).

**Figure 3.**
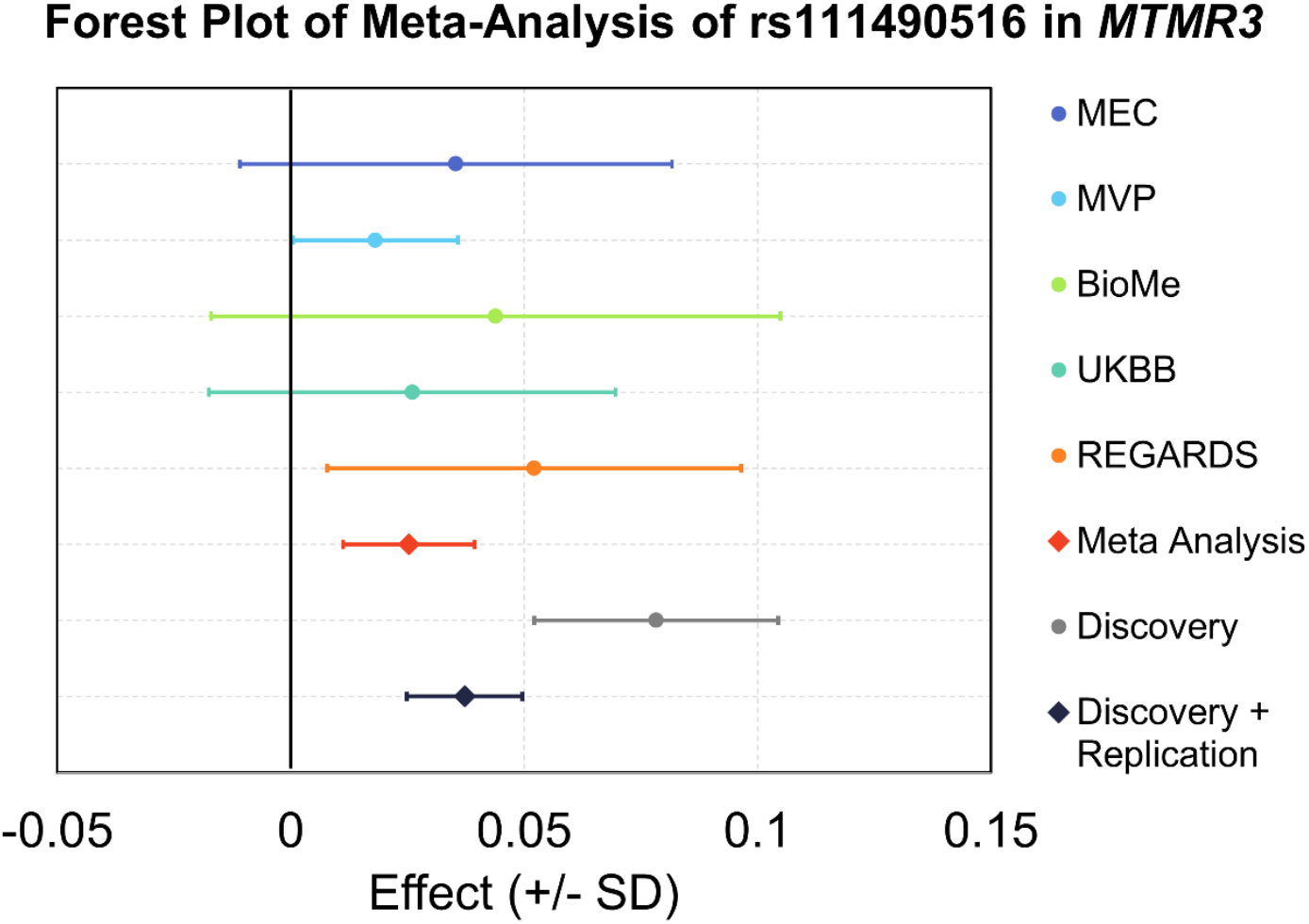
Forest plot of rs111490516 replication. All effect estimates (95% confidence interval) are oriented on the BMI increasing allele and are provided as standard deviation per allele. Actual beta values and *P*-values are in Supplementary Data 7.

To gain a better understanding of the potential functional consequence of the *MTMR3* locus, we used Ensembl VEP ^60^ to annotate all variants in high LD with our top SNP (R^2^ > 0.8 in the African population group using TOP-LD ^59^). Of the 54 variants in high LD, most were intronic or nearby *MTMR3* (**Supplementary Data 8**). Of these, four variants had a moderate CADD (combined annotation dependent depletion) score (scaled CADD > 10) with rs73394881 having the highest relative CADD score^61^, three of which lay within a possible enhancer (rs73396896, R^2^ = 0.884; rs73394881, R^2^ = 0.889; rs74832232, R^2^ = 0.889).

### Conditional Analyses

Conditional analysis using the most associated variant at each locus revealed two significant secondary signals after multiple testing correction (**Tables 1, Supplementary Data 9, Supplementary Figure 8**). These included a known BMI-associated index variant on chromosome 2 (rs62107261, β = -0.097, SE = 0.014, *P* = 2.06 × 10^-12^, near *ALKAL2*) ^40^, which was also the most significant variant at this locus in the European group analysis (**Supplementary Data 6**). We further identified rs78769612 on chromosome 18 (β = -0.100, SE = 0.019, *P* = 2.17 × 10^-7^, near *MC4R*). Although both secondary SNPs were in known BMI-associated loci, rs78769612 near *MC4R* was a new index variant.

We additionally assessed independence for the top variants in known loci, by conditioning on all previously-reported index variants ^5,29-48^. Two SNPs, rs2206277 in *TFAP2B* and rs3838785 in *BDNF*, remained significant after multiple test correction, indicating potentially novel signals in known loci (**Supplementary Data 10)**. The novel index variant from the internal conditional analysis, rs78769612 near *MC4R*, was not robust to this treatment, suggesting that this novel variant was not independent of known BMI variants. The LD matrix plots highlighted low LD (R^2^ range 0.018 – 0.342) between our top SNP at the *BDNF* locus, rs3838785, and previously published lead variants within 500 kb (**Supplementary Fig. 9**). Although our top SNP, rs2206277, in the *TFAP2B* locus was conditionally independent of previously published BMI-risk SNPs (β ≥ 90% of the unconditioned β and *P* < 6.25 × 10^-3^), this SNP was in high LD with two nearby published SNPs (R^2^ = 0.822 for rs987237 and R^2^ = 0.793 for rs72892910).

### Aggregate-based testing

We did not identify any novel gene regions through association tests at the genome-wide level (*P* < 5 × 10^-7^) when aggregating variants with MAF ≤ 1%. Nevertheless, we successfully replicated previous gene-based associations with the well-known melanocortin 4 receptor (*MC4R*) gene (*P* = 8.47 × 10^-8^), with 111 alleles across 37 sites within coding regions, enhancers, and promoters for *MC4R*. The *MC4R* locus was also identified in single-variant analyses.

### Fine-mapping

To pinpoint the most probable causal variant(s) underlying each of the 16 loci, we subsequently performed fine-mapping using PAINTOR ^53^. Assuming one causal variant per locus, the index variants were the most likely causal variants in 14 loci, with posterior probabilities (PP) ranging from 0.02 and 1.00, and seven of them had a PP above 0.50 (**Supplementary Data 11, Supplementary Fig 10**). Two intronic index variants, rs2307111 in *POC5* and rs1379871 in *DMD*, were particularly noteworthy with PP exceeding 0.98. In contrast, variants with the highest PP in *ADCY3* and *ZC3H4* were not the reported index variants, although the highest PP for the *ADCY3* locus was below 0.50, and thus not likely the causal variant underlying this signal. In the *ZC3H4* locus, the highest PP variant (rs55731973, PP = 0.77) was intronic, located in the 5’ UTR or upstream of *ZC3H4* depending on alternative transcripts, and resided in probable enhancer regions. Additionally, this variant was a significant cis-eQTL for *SAE1*^62^, another nearby downstream gene.

### PheWAS

To explore potential novel pleiotropy, we conducted association tests between the tag variant from our novel locus, rs111490516, and 538 PheCodes available in the MyCode and BioMe studies. No PheCode was significantly associated with rs111490516 following multiple test correction (*P* < 9.3 × 10^-5^). However, PheCode 327.3 (Sleep Apnea) and 327.32 (Obstructive Sleep Apnea) ranked among the top associated PheCodes (*P* < 0.001) (**Supplementary Data 12, Supplementary Fig 11**). Perhaps not coincidentally, obesity is one of the strongest risk factors for sleep apnea ^63^.

## DISCUSSION

By leveraging WGS data from a large multi-population study, we identified and replicated one novel low-frequency BMI variant in *MTMR3*, specific to the diversity of our sample. We also identified two common secondary signals in known BMI loci, supported gene-based associations for *MC4R*, and refined resolution in multiple loci by prioritizing candidate SNPs with high PP. Our discovery of the novel BMI-associated variant emphasizes the importance of studying diverse populations, which could further refine and expand the catalog of genes and variants that confer risk for obesity and potentially other disease traits.

The novel *MTMR3* variant, rs111490516, was most common in our African and Barbadian population groups (MAF = 13%) and of moderate frequency in our Dominican population group (MAF = 5%). We further replicated this association in study samples of similar population background. Yet, previous GWAS of BMI focusing on African ancestry individuals failed to identify a significant association in this region. It is not available for lookup in the most recent MVP BMI GWAS ^23^, although included in our replication. In one of the largest GWAS meta-analyses of imputed genotype data in African ancestry individuals with summary data available publicly, which was conducted by the African Ancestry Anthropometry Genetics Consortium (AAAGC, N up to 42,751)^37^, this variant was directionally consistent and suggestively associated (β = 0.042, *P* = 1.80 × 10^-4^, MAF = 12%)^37^. Similarly, in our replication analysis of 109,748 individuals with imputed genotypes, *MTMR3* (rs111490516) was suggestively significant (β = 0.025, *P* = 4.76 × 10^-4^, MAF = 11%). Therefore, the lack of discovery in prior publications is likely not due to insufficient power. As indicated by our fine-mapping results for this novel locus, our index SNP is likely not causal but could be in LD with a causal SNP and also poorly captured in studies relying on imputation. In other words, the causal variant underlying this locus may be nearby, less frequent, and on an LD block more frequent in a population poorly represented in other imputation reference panels, but well represented in our WGS and highly diverse sample (e.g., Caribbean admixed individuals). In this case, one would require sequencing data in a large sample size with the relevant haplotype to detect a significant association that was not able to be identified with imputation in a similar number of people.

The SNP rs111490516 lies in an intron of the *MTMR3* (myotubularin related protein 3) gene, with limited evidence of involvement in regulatory or functional protein activity. Other variants mapped to *MTMR3* have been associated with obesity-related traits in GWAS. In a study of 155,961 healthy and medication-free UKBB participants, rs5752989 near *MTMR3* was associated with fat-free mass (β = 0.115, *P* = 8.00 × 10^-9^, allele G frequency = 43%)^64^. In a meta-analysis of up to 628,000 BioBank Japan (BBJ), UKBB, and FinnGen (FG) participants, the same SNP was associated with body weight (β = - 0.010, *P* = 3.86 × 10^-8^, allele A frequency ranged from 51% in FG to 86% in BBJ) ^65^.

The primary cellular function of *MTMR3* relates to regulation of autophagy ^66^. Although there is no direct evidence linking *MTMR3* to obesity, previous studies have established a connection between *MTMR3* and related cardiometabolic traits. *MTMR3* was associated with LDL cholesterol (*P* = 1 × 10^-8^) in a GWAS meta-analysis of European, East Asian, South Asian, and African ancestry individuals ^67^. A potential mechanism was proposed later suggesting *MTMR3* may mediate the association between miRNA-4513 and total cholesterol ^68^. Furthermore, pyruvate dehydrogenase complex-specific knockout mice with high-fat diet induced obesity also exhibited increased blood glucose and higher expression levels of *MTMR3* ^69^. We utilized the Ensembl VEP database to explore predicted functional consequences of our novel locus. While there is limited knowledge on the biological implications of the lead variant and those in high LD, there are multiple lines of evidence supporting a role in obesity at this locus.

The use of WGS coupled with inclusion of non-European populations improved fine-mapping resolution, as has been shown previously ^47^. While there have been multiple attempts to fine-map previously identified BMI loci ^5,33,48^, no previous study has successfully identified BMI risk variants of high confidence at the *POC5* and *DMD* loci. By applying a Bayesian fine-mapping approach, we reduced associated signals to 95% credible sets of two likely causal SNPs. Functional annotation revealed that one of them, rs2307111, was a benign missense variant in *POC5* (NP_001092741.1:p.His36Arg) according to ClinVar ^70,71^, while the other is an intron variant in the promotor region of *DMD*. These two variants were also considered high-confidence causal variants (PP_rs2307111_ = 0.96, PP_rs1379871_ = 0.99) in a recent joint analysis of three biobanks (UKBB, FG, BBJ) ^72^. Notably, unlike in Kanai et al. where the PP appeared to be driven by the Europeans (for rs2307111: PP_UKBB_ = 0.96, PP_BBJ_ = 0.12, PP_FG_ = 0.01; for rs1379871: PP_UKBB_ = 1.00, PP_FG_ = 1.00), the effect alleles in our study were observed in high proportions across many non-European population groups (**Supplementary Data 5**).

In addition to our novel findings, 17 of the 18 identified variants reside in previously reported BMI-associated loci, highlighting the generalizability of the genes underlying BMI across populations, including *SEC16B, TMEM18, ETV5, GNPDA2, BDFN*, and *MC4R* ^5,34,48,73^. Three of the loci harbor genes implicated in severe and early-onset obesity – *ADCY3, BDNF*, and *MC4R* ^4^. We also consistently identified multiple association signals of high effect in *MC4R*, which is a well-established monogenic obesity gene, through our discovery analysis, internal conditional analysis, and rare variant aggregate analysis.

While our study included a large sample size of diverse populations and leveraged high quality WGS data from well-characterized and harmonized cohorts, our results should also be interpreted with the following limitations. First, although our study is large compared to other harmonized and sequenced data samples, the total study size is relatively modest compared to existing GWAS meta-analyses of common variants using imputed genotype data. Moreover, rare variants, such as those analyzed in our study, may require even larger sample sizes for novel discoveries. Even though our study is among the most racially, ethnically, and ancestrally diverse yet conducted, the European population group still represented 49% of our participants. On the other hand, diversity can contribute to added heterogeneity of effect sizes, potentially limiting discovery in the multi-population analysis. We sought to overcome this limitation by allowing for heterogeneous residual variances across population groups and examining population stratified results when samples sizes were adequate. Notably, all our genome-wide significant loci from population stratified analyses were also captured in the multi-population analysis, likely owing to our considerations of heterogenous effects, self-identity (population groups), and ancestry (genotype PCs). As has been shown by others ^47^, this underscores the importance of conducting multi-population analysis using appropriate methods that account for heterogeneity and minimize the risk of inflation or missed detection of loci that may vary in MAF or phenotypic effects across populations.

In summary, our study demonstrates the power of leveraging WGS data from diverse populations for new discoveries associated with BMI. As we enter the era of incorporating GWAS-based risk models in clinical practice, it is critical that we continue to diversify the data collected and analyzed in genomic research. Failure to do so risks further exacerbating health disparities for public health crisis such as obesity. Ultimately, our study brings us one step closer to understanding the complex genetic underpinnings of obesity, translating these leads into mechanistic insights, and developing targeted preventions and interventions to address this global public health challenge.

## Data Availability

The data used in our discovery analyses are available to applicants through dbGaP (NHLBI Trans-Omics for Precision Medicine WGS-TOPMed Data Access for the Scientific Community (https://topmed.nhlbi.nih.gov/topmed-data-access-scientific-community)).

## ACKNOWLEDGEMENTS

This work was funded in part by National Institutes of Health (NIH) grants (R01 DK122503, T32 HL007055, T32 HL129982, R01 HL142825, I01-BX003362, U01 HL120393, R01 HL68959, U01 HL072507, K08 HL136928, R01 HL119443, R01 HL055673-18S1, R01 HL92301, R01 HL67348, R01 NS058700, R0 AR48797, R01 DK071891, R01 AG058921, F32 HL085989, U01 HL089897, U01 HL089856, R01 HL093093, R01 HL133040, I01 BX003340, I01 BX004821, U01 HL072524, R01 HL104135-04S1, U01 HL054472, U01 HL054473, U01 HL054495, U01 HL054509, R01 HL055673 with supplement -18S1, R01 HL104608, R01 AI132476, R01 AI114555, R01 HL104608-S1, U01 HL072507, P20 GM109036, KL2 TR002490 and T32 HL129982, P01 HL132825, R35 CA197449, P01 CA134294, U19 CA203654, R01 HL113338, U01 HG009088, R01 HL142302, R01 DK124097, R01 DK110113, R01 DK107786, X01 HL134588, R01 HG010297, U01 HG007416, R01 HL105756) and contracts (HHSN268201800001I, HHSN268201500014C,) American Diabetes Association (ADA) grant #1-19-PDF-045, the General Clinical Research Center of the Wake Forest University School of Medicine (M01 RR07122), and a pilot grant from the Claude Pepper Older Americans Independence Center of Wake Forest University Health Sciences (P60 AG10484). A full list of study acknowledgements is detailed in the **Supplementary Note**.

