## Supplemental Files (Notes, Figures, and Tables) for "WHOLE GENOME SEQUENCING ANALYSIS OF BODY MASS INDEX IDENTIFIES NOVEL AFRICAN ANCESTRY-SPECIFIC RISK ALLELE"

**SUPPLEMENTARY ONLINE MATERIAL**

**Table of Contents**


| Supplementary Notes | | |
|---|---|---|
| Supplementary Note 1. | Replication cohort descriptions | 2-3 |
| Supplementary Note 2. | Investigator acknowledgements | 3-4 |
| Supplementary Note 3. | Study Acknowledgements | 4-16 |
| Supplementary Note 4 | NHLBI TOPMed: NHLBI Trans-Omics for Precision Medicine (TOPMed) consortium banner authorship | 16-23 |
| Supplementary Note 5 | References | 23 |
| **Supplementary Figures** | | |
| Supplementary Figure 1 | Scatter plots of PC1 vs. PC2 by population group | 24-26 |
| Supplementary Figure 2 | QQ Plots primary BMI GWAS | 27 |
| Supplementary Figure 3 | Regional association plots for primary BMI GWAS | 28-36 |
| Supplementary Figure 4 | Manhattan plot of African HARE group BMI GWAS | 37 |
| Supplementary Figure 5 | QQ Plot of African HARE group BMI GWAS | 38 |
| Supplementary Figure 6 | Manhattan plot of European HARE group BMI GWAS | 39 |
| Supplementary Figure 7 | QQ Plots of European HARE group BMI GWAS | 40 |
| Supplementary Figure 8 | Regional association plots of secondary signals in discovery | 41 |
| Supplementary Figure 9 | LD matrix heatmap for conditionally independent SNPs in known BMI-risk loci | 42-43 |
| Supplementary Figure 10 | Fine-mapping regional plots | 44-47 |
| Supplementary Figure 11 | PheWAS meta-analysis Manhattan plot | 48 |
| **Supplementary Data Tables** | | |
| Supplementary Data 1 | Participant counts by study and population group | 49 |
| Supplementary Data 2 | BMI and percent female by study | 50 |
| Supplementary Data 3 | BMI and percent female by population group | 51 |
| Supplementary Data 4 | Study-specific descriptive statistics of age and BMI | 52-55 |
| Supplementary Data 5 | Genome-wide significant (GWS) variants by locus with frequency by population group | 56 |
| Supplementary Data 6 | GWS variants by locus from African and European population groups | 57 |
| Supplementary Data 7 | Replication of rs111490516 | 58 |
| Supplementary Data 8 | Variant annotation from Variant Effect Predictor (VEP) for *MTMR3* locus | 59-61 |
| Supplementary Data 9 | Association results after conditioning on top index variant | 62 |
| Supplementary Data 10 | Association results after conditioning on known index variants | 63 |
| Supplementary Data 11 | PAINTOR fine-mapping results assuming a single causal variant | 64 |
| Supplementary Data 12 | PheWAS meta-analysis | 65 |

### SUPPLEMENTARY NOTE

#### Supplementary Note 1. REPLICATION COHORTS DESCRIPTIONS

***MEC*** is a population-based prospective cohort study including approximately 215,000 men and women from Hawaii and California [Kolonel, L. N. et al. A multiethnic cohort in Hawaii and Los Angeles: baseline characteristics. Am. J. 869 Epidemiol. 151, 346–357 (2000)]. All participants were 45-75 years of age at baseline, and primarily of five ancestries: Japanese Americans, African Americans, European Americans, Hispanic/Latinos, and Native Hawaiians. MEC was funded by the National Cancer Institute in 1993 to examine lifestyle risk factors and genetic susceptibility to cancer. All eligible cohort members completed baseline and follow-up questionnaires. Participants from the MEC sample in the current analyses included 3,825 women and 3,281 men who self-reported African American background, had measured height and weight available, and had genetic data available. Of these, 4,593 were genotyped on the MEGA chip and 2,513 were genotyped on the Illumina Human1M-Duo chip.

***MVP*** participants were recruited from over 60 Veterans Health Administration medical centers nationwide since 2011. The design of MVP has been previously described [1]. A unique feature of MVP is the linkage of a large biobank to an extensive, national, database from 2003 onward that integrates multiple elements such as diagnosis codes, procedure codes, laboratory values, and imaging reports, which permits detailed phenotyping of this large cohort. MVP has received ethical and study protocol approval by the Veterans Affairs Central Institutional Review Board in accordance with the principles outlined in the Declaration of Helsinki. DNA extracted from participants' blood was genotyped using a customized Affymetrix Axiom® biobank array, the MVP 1.0 Genotyping Array. The array was enriched for both common and rare genetic variants of clinical significance in different ethnic backgrounds. Quality-control procedures used to assign ancestry, remove low-quality samples and variants, and perform genotype imputation were previously described [2]. We excluded: duplicate samples, samples with more heterozygosity than expected an excess (>2.5%) of missing genotype calls, or discordance between genetically inferred sex and phenotypic gender. In addition, one individual from each pair of related individuals (more than second degree relatedness as measured by the KING software) were removed. The MVP participants were assigned to mutually exclusive racial/ethnic groups using HARE (Harmonized Ancestry and Race/Ethnicity), a machine learning algorithm that integrates genetically inferred ancestry with self-identified race/ethnicity [3]. The present study included non-Hispanic African Americans with both genotypic and phenotypic data for genetic association analyses. The phenotyping and analytical details of body mass index in the MVP were previously described [4]. SNP rs111490516 was imputed with quality score of 0.7083.

***The UK Biobank*** is a prospective cohort study with genetic and phenotypic data on more than 500,000 individuals, aged between 39–69 years. Study design, protocols, sample handling and quality control have been described in detail elsewhere (PMID: 25826379 and PMID: 30305743). African ancestry was determined using k-means clustering (PMID: 32692746). Briefly, clustering was performed by projecting the 1000 genomes reference panel dataset based on the PCA loadings from the UK Biobank. We performed k-means clustering with a pre-specified number of 4 clusters. Individuals from the UK Biobank that clustered with the AFR 1000G cluster were assigned African ancestry.

***REGARDS***: The Reasons for Geographic and Racial Differences in Stroke (REGARDS) project, sponsored by the National Institutes of Health (NIH), is a national study focusing on learning more about the factors that increase a person's risk of having a stroke. REGARDS is an observational study of risk factors for stroke in unrelated adults 45 years or older. 30,239 African American and European American participants were recruited between January 2003 and October 2007. The study design and objectives have been previously described [5]. MEGAEX genotype data is available for 8,837 African American and 1,716 European American REGARDS participants. The study is ongoing and will follow participants for many years.

***BioMe*** is an ongoing electronic medical record-linked biobank with more than 60,000 patients enrolled through the Mount Sinai Health System in New York. BioMe is a multiethnic biobank comprising individuals of African, Hispanic, European, Asian, and other ancestries [6]. Genotyping data is available on 32,595 individuals and was done using the Global Screening Array (GSA-24v1-0_A1). The data was cleaned for duplicate samples, discordant sex, heterozygosity rate that exceeded 6 SD from the population mean, call rate<95% at the site and individual level, and deviation from Hardy Weinberg equilibrium. Replication was conducted within self-reported African ancestry.

### Supplementary Note 2. INVESTIGATOR ACKNOWLEDGEMENTS


- Heather M. Highland was funded in part by NHLBI training grants (T32 HL007055, T32 HL129982) ADA Grant #1-19-PDF-045, and R01HL142825.
- Kyong-Mi Chang was funded in part by I01-BX003362.
- Matthew P. Conomos was funded in part by NIH U01HL-120393; contract HHSN268201800001I.
- Melanie E. Garrett was funded in part by NIH R01HL68959.
- Benjamin D. Heavner was funded in part by NIH U01HL-120393; contract HHSN268201800001I.
- James E. Hixson was funded in part by NIH U01HL072507.
- Brian D. Hobbs was funded in part by NIH K08 HL136928.
- Deepti Jain was funded in part by NIH R01HL-120393; U01HL-120393; contract HHSN268201800001I.
- Sharon LR Kardia was funded in part by NIH R01 HL119443, 3R01HL055673-18S1, HHSN268201500014C.
- Nicholette D. Palmer was funded in part by R01HL92301, R01HL67348, R01NS058700, R0 AR48797, R01DK071891, R01AG058921, the General Clinical Research Center of the Wake Forest University School of
  Medicine (M01 RR07122, F32 HL085989), the American Diabetes Association, and a pilot grant from the Claude Pepper Older Americans Independence Center of Wake Forest University Health Sciences (P60 AG10484).
  Elizabeth A. Regan was funded in part by NIH U01 HL089897 and U01 HL089856.
- Daniel E. Weeks was funded in part by NIH R01-HL093093, R01-HL133040.
- Peter W.F. Wilson was funded in part by NIH I01-BX003340, I01-BX004821.
- Donna K. Arnett was funded in part by NIH U01 HL072524 and R01 HL104135-04S1, U01 HL054472, U01 HL054473, U01 HL054495, and U01 HL054509,R01 HL055673 with supplement -18S1.
- Allison E. Ashley-Koch was funded in part by NIH R01HL68959.
- Kathleen C. Barnes was funded in part by NIH 5R01HL104608-09, R01AI132476, 1R01 AI114555, R01 HL104608-S1.
- Jiang He was funded in part by NIH U01HL072507, P20GM109036.
- Stephen T. McGarvey was funded in part by NIH R01-HL093093, R01-HL133040
- Laura M. Raffield was funded in part by NIH KL2TR002490 and T32HL129982.
- Marilyn J. Telen was funded in part by NIH R01HL68959.
- Scott T. Weiss was funded in part by P01 HL132825.
- Xihong Lin was funded in part by R35-CA197449, P01-CA134294, U19-CA203654, R01-HL113338, and U01-HG009088.
- Ching-Ti Liu was funded in part by R01DK122503.

- Ruth J.F. Loos was funded in part by NIH R01HL142302; R01DK124097; R01 DK110113; R01DK107786; X01HL134588
- Kari E. North was funded in part by R01HG010297; U01HG007416, DK122503.
- Anne E Justice, Geetha Chittoor, and Navya Josyula were funded in part by NIH NIDDK R01 DK 122503.
- Bruce M. Psaty was funded in part by HL105756.

### Supplementary Note 3. STUDY ACKNOWLEDGEMENTS

**NHLBI TOPMed Analysis Commons:** The TOPMed Analysis Commons was funded in part by NIH NHLBI Grant R01HL131136.

**NHLBI TOPMed: Genetics of Cardiometabolic Health in the Amish (Amish)**
The TOPMed component of the Amish Research Program was supported by NIH grants R01 HL121007, U01 HL072515, and R01 AG18728.

**NHLBI TOPMed: Atherosclerosis Risk in Communities Study VTE cohort (ARIC)**
The Atherosclerosis Risk in Communities study has been funded in whole or in part with Federal funds from the National Heart, Lung, and Blood Institute, National Institutes of Health, Department of Health and Human Services (contract numbers HHSN268201700001I, HHSN268201700002I, HHSN268201700003I, HHSN268201700004I and HHSN268201700005I). The authors thank the staff and participants of the ARIC study for their important contributions.

**NHLBI TOPMed: New Approaches for Empowering Studies of Asthma in Populations of African Descent - Barbados Asthma Genetics Study (BAGS)**
We gratefully acknowledge the contributions of Pissamai and Trevor Maul, Paul Levett, Anselm Hennis, P. Michele Lashley, Raana Naidu, Malcolm Howitt and Timothy Roach, and the numerous health care providers, and community clinics and co-investigators who assisted in the phenotyping and collection of DNA samples, and the families and patients for generously donating DNA samples to the Barbados Asthma Genetics Study (BAGS). Funding for BAGS was provided by National Institutes of Health (NIH) R01HL104608, R01HL087699, and HL104608 S1.

**NHLBI TOPMed: Mount Sinai BioMe Biobank (BioMe)**
The Mount Sinai BioMe Biobank has been supported by The Andrea and Charles Bronfman Philanthropies and in part by Federal funds from the NHLBI and NHGRI (U01HG00638001; U01HG007417; X01HL134588). We thank all participants in the Mount Sinai Biobank. We also thank all our recruiters who have assisted and continue to assist in data collection and management and are grateful for the computational resources and staff expertise provided by Scientific Computing at the Icahn School of Medicine at Mount Sinai.

**NHLBI TOPMed: Coronary Artery Risk Development in Young Adults (CARDIA)**
The Coronary Artery Risk Development in Young Adults Study (CARDIA) is conducted and supported by the National Heart, Lung, and Blood Institute (NHLBI) in collaboration with the University of Alabama at Birmingham (HHSN268201800005I & HHSN268201800007I), Northwestern University (HHSN268201800003I), University of Minnesota (HHSN268201800006I), and Kaiser Foundation Research Institute (HHSN268201800004I). CARDIA was also partially supported by the Intramural Research Program of the National Institute on Aging (NIA) and an intra-agency agreement between NIA and NHLBI (AG0005).

**NHLBI TOPMed: Cleveland Clinic Atrial Fibrillation Study (CCAF)**

This study was supported by the National Institutes of Health (NIH) grants R01 HL 090620 and R01 HL 111314, the NIH National Center for Research Resources for Case Western Reserve University and Cleveland Clinic Clinical and Translational Science Award UL1-RR024989, the Cleveland Clinic Department of Cardiovascular Medicine philanthropy research funds, and the Tomsich Atrial Fibrillation Research Fund.

**NHLBI TOPMed: Cleveland Family Study - WGS Collaboration (CFS)**
The Cleveland Family Study has been supported in part by National Institutes of Health grants [R01-HL046380, KL2-RR024990, R35-HL135818, and R01-HL113338].

**NHLBI TOPMed: Cardiovascular Health Study (CHS)**
Cardiovascular Health Study: This research was supported by contracts HHSN268201200036C, HHSN268200800007C, HHSN268201800001C, N01HC55222, N01HC85079, N01HC85080, N01HC85081, N01HC85082, N01HC85083, N01HC85086, 75N92021D00006, and grants U01HL080295 and U01HL130114 from the National Heart, Lung, and Blood Institute (NHLBI), with additional contribution from the National Institute of Neurological Disorders and Stroke (NINDS). Additional support was provided by R01AG023629 from the National Institute on Aging (NIA). A full list of principal CHS investigators and institutions can be found at CHS-NHLBI.org. The content is solely the responsibility of the authors and does not necessarily represent the official views of the National Institutes of Health.

**NHLBI TOPMed: Genetic Epidemiology of COPD Study (COPDGene)**
The COPDGene project described was supported by Award Number U01 HL089897 and Award Number U01 HL089856 from the National Heart, Lung, and Blood Institute. The content is solely the responsibility of the authors and does not necessarily represent the official views of the National Heart, Lung, and Blood Institute or the National Institutes of Health. The COPDGene project is also supported by the COPD Foundation through contributions made to an Industry Advisory Board comprised of AstraZeneca, Boehringer Ingelheim, GlaxoSmithKline, Novartis, Pfizer, Siemens and Sunovion. A full listing of COPDGene investigators can be found at: http://www.copdgene.org/directory

**COPDGene® Investigators – Core Units:** Administrative Center: James D. Crapo, MD (PI); Edwin K. Silverman, MD, PhD (PI); Barry J. Make, MD; Elizabeth A. Regan, MD, PhD. ***Genetic Analysis Center:*** Terri Beaty, PhD; Ferdouse Begum, PhD; Peter J. Castaldi, MD, MSc; Michael Cho, MD; Dawn L. DeMeo, MD, MPH; Adel R. Boueiz, MD; Marilyn G. Foreman, MD, MS; Eitan Halper-Stromberg; Lystra P. Hayden, MD, MMSc; Craig P. Hersh, MD, MPH; Jacqueline Hetmanski, MS, MPH; Brian D. Hobbs, MD; John E. Hokanson, MPH, PhD; Nan Laird, PhD; Christoph Lange, PhD; Sharon M. Lutz, PhD; Merry-Lynn McDonald, PhD; Margaret M. Parker, PhD; Dmitry Prokopenko, Ph.D; Dandi Qiao, PhD; Elizabeth A. Regan, MD, PhD; Phuwanat Sakornsakolpat, MD; Edwin K. Silverman, MD, PhD; Emily S. Wan, MD; Sungho Won, PhD. ***Imaging Center:*** Juan Pablo Centeno; Jean-Paul Charbonnier, PhD; Harvey O. Coxson, PhD; Craig J. Galban, PhD; MeiLan K. Han, MD, MS; Eric A. Hoffman, Stephen Humphries, PhD; Francine L. Jacobson, MD, MPH; Philip F. Judy, PhD; Ella A. Kazerooni, MD; Alex Kluiber; David A. Lynch, MB; Pietro Nardelli, PhD; John D. Newell, Jr., MD; Aleena Notary; Andrea Oh, MD; Elizabeth A. Regan, MD, PhD; James C. Ross, PhD; Raul San Jose Estepar, PhD; Joyce Schroeder, MD; Jered Sieren; Berend C. Stoel, PhD; Juerg Tschirren, PhD; Edwin Van Beek, MD, PhD; Bram van Ginneken, PhD; Eva van Rikxoort, PhD; Gonzalo Vegas Sanchez-Ferrero, PhD; Lucas Veitel; George R. Washko, MD; Carla G. Wilson, MS; ***PFT QA Center, Salt Lake City, UT:*** Robert Jensen, PhD. **Data Coordinating Center and *Biostatistics, National Jewish Health, Denver, CO:*** Douglas Everett, PhD; Jim Crooks, PhD; Katherine Pratte, PhD; Matt Strand, PhD; Carla G. Wilson, MS. **Epidemiology Core, University of Colorado *Anschutz Medical Campus, Aurora, CO:*** John E. Hokanson, MPH, PhD; Gregory Kinney, MPH, PhD; Sharon M. Lutz, PhD; Kendra A. Young, PhD. ***Mortality Adjudication Core:*** Surya P. Bhatt, MD; Jessica Bon, MD; Alejandro A. Diaz, MD, MPH;

MeiLan K. Han, MD, MS; Barry Make, MD; Susan Murray, ScD; Elizabeth Regan, MD; Xavier Soler, MD; Carla G. Wilson, MS. ***Biomarker Core:*** Russell P. Bowler, MD, PhD; Katerina Kechris, PhD; Farnoush Banaei-Kashani, Ph.D. **COPDGene® Investigators – Clinical Centers:** ***Ann Arbor VA:*** Jeffrey L. Curtis, MD; Perry G. Pernicano, MD. ***Baylor College of Medicine, Houston, TX:*** Nicola Hanania, MD, MS; Mustafa Atik, MD; Aladin Boriek, PhD; Kalpatha Guntupalli, MD; Elizabeth Guy, MD; Amit Parulekar, MD. ***Brigham and Women's Hospital, Boston, MA:*** Dawn L. DeMeo, MD, MPH; Alejandro A. Diaz, MD, MPH; Lystra P. Hayden, MD; Brian D. Hobbs, MD; Craig Hersh, MD, MPH; Francine L. Jacobson, MD, MPH; George Washko, MD. ***Columbia University, New York, NY:*** R. Graham Barr, MD, DrPH; John Austin, MD; Belinda D'Souza, MD; Byron Thomashow, MD. ***Duke University Medical Center, Durham, NC:*** Neil MacIntyre, Jr., MD; H. Page McAdams, MD; Lacey Washington, MD. ***Grady Memorial Hospital, Atlanta, GA:*** Eric Flenaugh, MD; Silanth Terpenning, MD. ***HealthPartners Research Institute, Minneapolis, MN:*** Charlene McEvoy, MD, MPH; Joseph Tashjian, MD. ***Johns Hopkins University, Baltimore, MD:*** Robert Wise, MD; Robert Brown, MD; Nadia N. Hansel, MD, MPH; Karen Horton, MD; Allison Lambert, MD, MHS; Nirupama Putcha, MD, MHS. ***Lundquist Institute for Biomedical Innovationat Harbor UCLA Medical Center, Torrance, CA:*** Richard Casaburi, PhD, MD; Alessandra Adami, PhD; Matthew Budoff, MD; Hans Fischer, MD; Janos Porszasz, MD, PhD; Harry Rossiter, PhD; William Stringer, MD. ***Michael E. DeBakey VAMC, Houston, TX:*** Amir Sharafkhaneh, MD, PhD; Charlie Lan, DO. ***Minneapolis VA:*** Christine Wendt, MD; Brian Bell, MD; Ken M. Kunisaki, MD, MS. ***National Jewish Health, Denver, CO:*** Russell Bowler, MD, PhD; David A. Lynch, MB. ***Reliant Medical Group, Worcester, MA:*** Richard Rosiello, MD; David Pace, MD. ***Temple University, Philadelphia, PA:*** Gerard Criner, MD; David Ciccolella, MD; Francis Cordova, MD; Chandra Dass, MD; Gilbert D'Alonzo, DO; Parag Desai, MD; Michael Jacobs, PharmD; Steven Kelsen, MD, PhD; Victor Kim, MD; A. James Mamary, MD; Nathaniel Marchetti, DO; Aditi Satti, MD; Kartik Shenoy, MD; Robert M. Steiner, MD; Alex Swift, MD; Irene Swift, MD; Maria Elena Vega-Sanchez, MD. ***University of Alabama, Birmingham, AL:*** Mark Dransfield, MD; William Bailey, MD; Surya P. Bhatt, MD; Anand Iyer, MD; Hrudaya Nath, MD; J. Michael Wells, MD. ***University of California, San Diego, CA:*** Douglas Conrad, MD; Xavier Soler, MD, PhD; Andrew Yen, MD. ***University of Iowa, Iowa City, IA:*** Alejandro P. Comellas, MD; Karin F. Hoth, PhD; John Newell, Jr., MD; Brad Thompson, MD. ***University of Michigan, Ann Arbor, MI:*** MeiLan K. Han, MD MS; Ella Kazerooni, MD MS; Wassim Labaki, MD MS; Craig Galban, PhD; Dharshan Vummidi, MD. ***University of Minnesota, Minneapolis, MN:*** Joanne Billings, MD; Abbie Begnaud, MD; Tadashi Allen, MD. ***University of Pittsburgh, Pittsburgh, PA:*** Frank Sciurba, MD; Jessica Bon, MD; Divay Chandra, MD, MSc; Joel Weissfeld, MD, MPH . ***University of Texas Health, San Antonio, San Antonio, TX:*** Antonio Anzueto, MD; Sandra Adams, MD; Diego Maselli-Caceres, MD; Mario E. Ruiz, MD; Harjinder Singh.

**NHLBI TOPMed: The Genetic Epidemiology of Asthma in Costa Rica - Asthma in Costa Rica cohort (CRA)**

**NHLBI TOPMed: Diabetes Heart Study (DHS)**
This work was supported by R01 HL92301, R01 HL67348, R01 NS058700, R01 AR48797, R01 DK071891, R01 AG058921, the General Clinical Research Center of the Wake Forest University School of Medicine (M01 RR07122, F32 HL085989), the American Diabetes Association, and a pilot grant from the Claude Pepper Older Americans Independence Center of Wake Forest University Health Sciences (P60 AG10484).

**NHLBI TOPMed: Framingham Heart Study (FHS)**
The Framingham Heart Study (FHS) acknowledges the support of contracts NO1-HC-25195, HHSN268201500001I and 75N92019D00031 from the National Heart, Lung and Blood Institute and grant supplement R01 HL092577-06S1 for this research. We also acknowledge the dedication of the FHS study participants without whom this research would not be possible. Dr. Vasan is supported in part by

the Evans Medical Foundation and the Jay and Louis Coffman Endowment from the Department of Medicine, Boston University School of Medicine.

**NHLBI TOPMed: Gene-Environment, Admixture and Latino Asthmatics Study (GALAII)**
The Genes-environments and Admixture in Latino Americans (GALA II) Study was supported by the National Heart, Lung, and Blood Institute of the National Institute of Health (NIH) grants R01HL117004 and X01HL134589; study enrollment supported by the Sandler Family Foundation, the American Asthma Foundation, the RWJF Amos Medical Faculty Development Program, Harry Wm. and Diana V. Hind Distinguished Professor in Pharmaceutical Sciences II and the National Institute of Environmental Health Sciences grant R01ES015794 . The GALA II study collaborators include Shannon Thyne, UCSF; Harold J. Farber, Texas Children's Hospital; Denise Serebrisky, Jacobi Medical Center; Rajesh Kumar, Lurie Children's Hospital of Chicago; Emerita Brigino-Buenaventura, Kaiser Permanente; Michael A. LeNoir, Bay Area Pediatrics; Kelley Meade, UCSF Benioff Children’s Hospital, Oakland; William Rodriguez-Cintron, VA Hospital, Puerto Rico; Pedro C. Avila, Northwestern University; Jose R. Rodriguez-Santana, Centro de Neumologia Pediatrica; Luisa N. Borrell, City University of New York; Adam Davis, UCSF Benioff Children's Hospital, Oakland; Saunak Sen, University of Tennessee and Fred Lurmann, Sonoma Technologies, Inc. The authors acknowledge the families and patients for their participation and thank the numerous health care providers and community clinics for their support and participation in GALA II. In particular, the authors thank study coordinator Sandra Salazar; the recruiters who obtained the data: Duanny Alva, MD, Gaby Ayala-Rodriguez, Lisa Caine, Elizabeth Castellanos, Jaime Colon, Denise DeJesus, Blanca Lopez, Brenda Lopez, MD, Louis Martos, Vivian Medina, Juana Olivo, Mario Peralta, Esther Pomares, MD, Jihan Quraishi, Johanna Rodriguez, Shahdad Saeedi, Dean Soto, Ana Taveras; and the lab researcher Celeste Eng who processed the biospecimens.

**NHLBI TOPMed: Genetic Studies of Atherosclerosis Risk (GeneSTAR)**
The Johns Hopkins Genetic Study of Atherosclerosis Risk (GeneSTAR) was supported by grants from the National Institutes of Health through the National Heart, Lung, and Blood Institute (HL49762, HL071025, U01HL72518, HL087698, HL092165, HL099747, K23HL105897, HL112064) and the National Institute of Nursing Research (NR0224103), by a grant from the National Center for Research Resources (M01-RR000052) to the Johns Hopkins General Clinical Research Center, and by a grant from the National Center for Research Resources and the National Center for Advancing Translational Sciences (UL1 RR 025005) to the Johns Hopkins Institute for Clinical and Translational Research.

**NHLBI TOPMed: Genetic Epidemiology Network of Arteriopathy (GENOA)**
Support for GENOA was provided by the National Heart, Lung and Blood Institute (U01 HL054457, U01 HL054464, U01 HL054481, R01 HL119443, and R01 HL087660) of the National Institutes of Health. WGS for “NHLBI TOPMed: Genetic Epidemiology Network of Arteriopathy” (phs001345) was performed at the Mayo Clinic Genotyping Core, the DNA Sequencing and Gene Analysis Center at the University of Washington (3R01HL055673-18S1), and the Broad Institute (HHSN268201500014C) for their genotyping and sequencing services. We would like to thank the GENOA participants.

**NHLBI TOPMed: Genetic Epidemiology Network of Salt Sensitivity (GenSalt)**
The Genetic Epidemiology Network of Salt-Sensitivity (GenSalt) was supported by research grants (U01HL072507, R01HL087263, and R01HL090682) from the National Heart, Lung and Blood Institute, National Institutes of Health, Bethesda, MD.

**NHLBI TOPMed: Genetics of Lipid Lowering Drugs and Diet Network (GOLDN)**
GOLDN biospecimens, baseline phenotype data, and intervention phenotype data were collected with funding from National Heart, Lung and Blood Institute (NHLBI) grant U01 HL072524. Whole-genome sequencing in GOLDN was funded by NHLBI grant R01 HL104135 and supplement R01 HL104135-04S1.

**NHLBI TOPMed: Hispanic Community Health Study - Study of Latinos (HCHS_SOL)**
The Hispanic Community Health Study/Study of Latinos is a collaborative study supported by contracts from the National Heart, Lung, and Blood Institute (NHLBI) to the University of North Carolina (HHSN268201300001I / N01-HC-65233), University of Miami (HHSN268201300004I / N01-HC-65234), Albert Einstein College of Medicine (HHSN268201300002I / N01-HC-65235), University of Illinois at Chicago – HHSN268201300003I / N01-HC-65236 Northwestern Univ), and San Diego State University (HHSN268201300005I / N01-HC-65237). The following Institutes/Centers/Offices have contributed to the HCHS/SOL through a transfer of funds to the NHLBI: National Institute on Minority Health and Health Disparities, National Institute on Deafness and Other Communication Disorders, National Institute of Dental and Craniofacial Research, National Institute of Diabetes and Digestive and Kidney Diseases, National Institute of Neurological Disorders and Stroke, NIH Institution-Office of Dietary Supplements. All HCHS/SOL participants provided informed consent, and the study was approved by the Institutional Review Board of local field centers, coordinating center and laboratories.

**NHLBI TOPMed: Heart and Vascular Health Study (HVH)**
The Heart and Vascular Health Study was supported by grants HL068986, HL085251, HL095080, and HL073410 from the National Heart, Lung, and Blood Institute.

**NHLBI TOPMed: Hypertension Genetic Epidemiology Network (HyperGEN)**
The HyperGEN Study is part of the National Heart, Lung, and Blood Institute (NHLBI) Family Blood Pressure Program; collection of the data represented here was supported by grants U01 HL054472 (MN Lab), U01 HL054473 (DCC), U01 HL054495 (AL FC), and U01 HL054509 (NC FC). The HyperGEN: Genetics of Left Ventricular Hypertrophy Study was supported by NHLBI grant R01 HL055673 with whole-genome sequencing made possible by supplement -18S1.

**NHLBI TOPMed: Jackson Heart Study (JHS)**
The Jackson Heart Study (JHS) is supported and conducted in collaboration with Jackson State University (HHSN268201800013I), Tougaloo College (HHSN268201800014I), the Mississippi State Department of Health (HHSN268201800015I) and the University of Mississippi Medical Center (HHSN268201800010I, HHSN268201800011I and HHSN268201800012I) contracts from the National Heart, Lung, and Blood Institute (NHLBI) and the National Institute on Minority Health and Health Disparities (NIMHD). The authors also wish to thank the staffs and participants of the JHS. JHS disclaimer- The views expressed in this manuscript are those of the authors and do not necessarily represent the views of the National Heart, Lung, and Blood Institute; the National Institutes of Health; or the U.S. Department of Health and Human Services.

**NHLBI TOPMed: Lung Tissue Research Consortium (LTRC)**

**NHLBI TOPMed: Mayo Clinic Venous Thromboembolism Study (Mayo_VTE)**
Funded, in part, by grants from the National Institutes of Health, National Heart, Lung and Blood Institute (HL66216 and HL83141). the National Human Genome Research Institute (HG04735, HG06379), and research support provided by Mayo Foundation.

**NHLBI TOPMed: Multi-Ethnic Study of Atherosclerosis (MESA)**
Whole genome sequencing (WGS) for the Trans-Omics in Precision Medicine (TOPMed) program was supported by the National Heart, Lung and Blood Institute (NHLBI). WGS for "NHLBI TOPMed: Multi-Ethnic Study of Atherosclerosis (MESA)" (phs001416.v3.p1) was performed at the Broad Institute of MIT and Harvard (3U54HG003067-13S1). Centralized read mapping and genotype calling, along with variant quality metrics and filtering were provided by the TOPMed Informatics Research Center (3R01HL-117626-02S1). Phenotype harmonization, data management, sample-identity QC, and general

study coordination, were provided by the TOPMed Data Coordinating Center (3R01HL-120393-02S1), and TOPMed MESA Multi-Omics (HHSN2682015000031/HSN26800004). The MESA projects are conducted and supported by the National Heart, Lung, and Blood Institute (NHLBI) in collaboration with MESA investigators. Support for the Multi-Ethnic Study of Atherosclerosis (MESA) projects are conducted and supported by the National Heart, Lung, and Blood Institute (NHLBI) in collaboration with MESA investigators. Support for MESA is provided by contracts 75N92020D00001, HHSN268201500003I, N01-HC-95159, 75N92020D00005, N01-HC-95160, 75N92020D00002, N01-HC-95161, 75N92020D00003, N01-HC-95162, 75N92020D00006, N01-HC-95163, 75N92020D00004, N01-HC-95164, 75N92020D00007, N01-HC-95165, N01-HC-95166, N01-HC-95167, N01-HC-95168, N01-HC-95169, UL1-TR-000040, UL1-TR-001079, UL1-TR-001420, UL1TR001881, DK063491, and R01HL105756. The authors thank the other investigators, the staff, and the participants of the MESA study for their valuable contributions.  A full list of participating MESA investigators and institutes can be found at http://www.mesa-nhlbi.org.

**NHLBI TOPMed: Massachusetts General Hospital Atrial Fibrillation Study (MGH_AF)**

**NHLBI TOPMed: Outcome Modifying Genes in Sickle Cell Disease (OMG_SCD)**
The OMG-SCD study was administrated by Marilyn J. Telen, M.D. and Allison E. Ashley-Koch, Ph.D. from Duke University Medical Center and collection of the data set was supported by grants HL068959 and HL079915 from the National Heart, Lung, and Blood Institute (NHLBI) of the National Institute of Health (NIH).

**NHLBI TOPMed: Partners Healthcare Biorepository (Partners)**

**NHLBI TOPMed: Whole Genome Sequencing to Identify Causal Genetic Variants Influencing CVD Risk - San Antonio Family Studies (SAFS)**
Collection of the San Antonio Family Study data was supported in part by National Institutes of Health (NIH) grants R01 HL045522, MH078143, MH078111 and MH083824; and whole genome sequencing of SAFS subjects was supported by U01 DK085524 and R01 HL113323. We are very grateful to the participants of the San Antonio Family Study for their continued involvement in our research programs.

**NHLBI TOPMed: Study of African Americans, Asthma, Genes and Environment (SAGE)**
The Study of African Americans, Asthma, Genes and Environments (SAGE) was supported by by the National Heart, Lung, and Blood Institute of the National Institute of Health (NIH) grants R01HL117004 and X01HL134589; study enrollment supported by the Sandler Family Foundation, the American Asthma Foundation, the RWJF Amos Medical Faculty Development Program, Harry Wm. and Diana V. Hind Distinguished Professor in Pharmaceutical Sciences II. The SAGE study collaborators include Harold J. Farber, Texas Children's Hospital; Emerita Brigino-Buenaventura, Kaiser Permanente; Michael A. LeNoir, Bay Area Pediatrics; Kelley Meade, UCSF Benioff Children's Hospital, Oakland; Luisa N. Borrell, City University of New York; Adam Davis, UCSF Benioff Children's Hospital, Oakland and Fred Lurmann, Sonoma Technologies, Inc. The authors acknowledge the families and patients for their participation and thank the numerous health care providers and community clinics for their support and participation in SAGE. In particular, the authors thank study coordinator Sandra Salazar; the recruiters who obtained the data: Lisa Caine, Elizabeth Castellanos, Brenda Lopez, MD, Shahdad Saeedi; and the lab researcher Celeste Eng who processed the biospecimens.

**NHLBI TOPMed: Samoan Adiposity Study (Samoan)**
Date collection was funded by NIH grant R01-HL093093. We thank the Samoan participants of the study and local village authorities. We acknowledge the support of the Samoan Ministry of Health and the Samoa Bureau of Statistics for their support of this research.

We would also like to acknowledge the Samoan Obesity, Lifestyle and Genetic Adaptations Study (OLaGA) Group: Ranjan Deka, Dept. of Environmental Health, University of Cincinnati; Nicola L. Hawley, Dept. of Chronic Disease Epidemiology, Yale University; Stephen T McGarvey, Dept. of Epidemiology and International Health Institute, and Dept. of Anthropology, Brown University; Ryan L Minster, Dept. of Human Genetics, University of Pittsburgh; Take Naseri, Ministry of Health, Government of Samoa; Muagututi'a Sefuiva Reupena, Lutia I Puava Ae Mapu I Fagalele; Daniel E. Weeks, Depts. of Human Genetics and Biostatistics, University of Pittsburgh.

**NHLBI TOPMed: Taiwan Study of Hypertension using Rare Variants (THRV)**
The Rare Variants for Hypertension in Taiwan Chinese (THRV) is supported by the National Heart, Lung, and Blood Institute (NHLBI) grant (R01HL111249) and its participation in TOPMed is supported by an NHLBI supplement (R01HL111249-04S1). THRV is a collaborative study between Washington University in St. Louis, LA BioMed at Harbor UCLA, University of Texas in Houston, Taichung Veterans General Hospital, Taipei Veterans General Hospital, Tri-Service General Hospital, National Health Research Institutes, National Taiwan University, and Baylor University. THRV is based (substantially) on the parent SAPPHIRe study, along with additional population-based and hospital-based cohorts. SAPPHIRe was supported by NHLBI grants (U01HL54527, U01HL54498) and Taiwan funds, and the other cohorts were supported by Taiwan funds.

**NHLBI TOPMed: Vanderbilt Atrial Fibrillation Ablation Registry (VAFAR)**

**NHLBI TOPMed: Vanderbilt Genetic Basis of Atrial Fibrillation (VU_AF)**

**NHLBI TOPMed: Treatment of Pulmonary Hypertension and Sickle Cell Disease with Sildenafil Therapy (walk_PHaSST)**
We thank Dr. Mark Gladwin and the investigators of the Walk-PHasst study and the patients who participated in the study. We also thanks the walk-PHaSST clinical site team: Albert Einstein College of Medicine: Jane Little and Verlene Davis; Columbia University: Robyn Barst, Erika Rosenzweig, Margaret Lee and Daniela Brady; UCSF Benioff Children's Hospital Oakland: Claudia Morris, Ward Hagar, Lisa Lavrisha, Howard Rosenfeld, and Elliott Vichinsky; Children's Hospital of Pittsburgh of UPMC: Regina McCollum; Hammersmith Hospital, London: Sally Davies, Gaia Mahalingam, Sharon Meehan, Ofelia Lebanto, and Ines Cabrita; Howard University: Victor Gordeuk, Oswaldo Castro, Onyinye Onyekwere, Vandana Sachdev, Alvin Thomas, Gladys Onojobi, Sharmin Diaz, Margaret Fadojutimi-Akinsiku, and Randa Aladdin; Johns Hopkins University: Reda Girgis, Sophie Lanzkron and Durrant Barasa; NHLBI: Mark Gladwin, Greg Kato, James Taylor, Wynona Coles, Catherine Seamon, Mary Hall, Amy Chi, Cynthia Brenneman, Wen Li, and Erin Smith; University of Colorado: Kathryn Hassell, David Badesch, Deb McCollister and Julie McAfee; University of Illinois at Chicago: Dean Schraufnagel, Robert Molokie, George Kondos, Patricia Cole-Saffold, and Lani Krauz; National Heart & Lung Institute, Imperial College London: Simon Gibbs. Thanks also to the data coordination center team from Rho, Inc.: Nancy Yovetich, Rob Woolson, Jamie Spencer, Christopher Woods, Karen Kesler, Vickie Coble, and Ronald W. Helms. We also thank Dr. Yingze Zhang for directing the Walk-PHasst repository and Dr. Mehdi Nouraie for maintaining the Walk-PHasst database and Dr. Jonathan Goldsmith as a NIH program director for this study. Special thanks to the volunteers who participated in the Walk-PHaSST study. This project was funded with federal funds from the NHLBI, NIH, Department of Health and Human Services, under contract HHSN268200617182C. This study is registered at www.clinicaltrials.gov as NCT00492531. Detail description of the study was published in Blood, 2011 118:855-864, Machado et al "Hospitalization for pain in patients with sickle cell disease treated with sildenafil for elevated TRV and low exercise capacity".

**NHLBI TOPMed: Women's Genome Health Study (WGHS)**

The WGHS is supported by the National Heart, Lung, and Blood Institute (HL043851 and HL080467) and the National Cancer Institute (CA047988 and UM1CA182913). The most recent cardiovascular endpoints were supported by ARRA funding HL099355.

**NHLBI TOPMed: Women's Health Initiative (WHI)**
The WHI program is funded by the National Heart, Lung, and Blood Institute, National Institutes of Health, U.S. Department of Health and Human Services through contracts 75N92021D00001, 75N92021D00002, 75N92021D00003, 75N92021D00004, 75N92021D00005.

**The Multiethnic Cohort (MEC)** is a population-based prospective cohort study including approximately 215,000 men and women from Hawaii and California. All participants were 45-75 years of age at baseline, and primarily of 5 ancestries: Japanese Americans, African Americans, European Americans, Hispanic/Latinos, and Native Hawaiians. (PMIDs: 10695593; 23449381) MEC was funded by the National Cancer Institute in 1993 to examine lifestyle risk factors and genetic susceptibility to cancer. All eligible cohort members completed baseline and follow-up questionnaires. Within the PAGE II investigation, MEC proposes to study: 1) diseases for which we have DNA available for large numbers of cases and controls (breast, prostate, and colorectal cancer, diabetes, and obesity); 2) common traits that are risk factors for these diseases (e.g., body mass index / weight, waist-to-hip ratio, height), and 3) relevant disease-associated biomarkers (e.g., fasting insulin and lipids, steroid hormones). The specific aims are: 1) to determine the population-based epidemiologic profile (allele frequency, main effect, heterogeneity by disease characteristics) of putative causal variants in the five racial/ethnic groups in MEC; 2) for variants displaying effect heterogeneity across ethnic/racial groups, we will utilize differences in LD to identify a more complete spectrum of associated variants at these loci; 3) investigate gene x gene and gene x environment interactions to identify modifiers; 4) examine the associations of putative causal variants with already measured intermediate phenotypes (e.g., plasma insulin, lipids, steroid hormones); and 5) for variants that do not fall within known genes, start to investigate their relationships with gene expression and epigenetic patterns in small genomic studies. The studies listed here are individuals of African and Laitno American ancestry/ethnicity who were part of the breast cancer or prostate cancer case/controls substuides. (dbGaP study accession number: phs000220).

**REGARDS:** This REGARDS research was supported by the National Institutes of Health (NIH) National Heart, Lung, and Blood Institute R01HL136666 and T32HL007457. The REGARDS study is supported by a cooperative agreement U01 NS041588 from the National Institute of Neurological Disorders and Stroke, National Institutes of Health, U.S. Department of Health and Human Services. The content is solely the responsibility of the authors and does not necessarily represent the official views of the National Institute of Neurological Disorders and Stroke or the National Institutes of Health. Representatives of the funding agency have been involved in the review of the manuscript but not directly involved in the collection, management, analysis, or interpretation of the data. The authors thank the other investigators, the staff, and the participants of the REGARDS study for their valuable contributions. A full list of participating REGARDS investigators and institutions can be found at http://www.regardsstudy.org.

**Million Veteran Program (MVP):** We are grateful to all the MVP investigators; a list of MVP investigators can be found in the appendix. This research is supported by funding from the Department of Veterans Affairs Office of Research and Development, Million Veteran Program Grant I01-BX003340 and I01-BX004821. This publication does not represent the views of the Department of Veterans Affairs or the United States Government.

*MVP Program Office*
- Sumitra Muralidhar, Ph.D., Program Director

US Department of Veterans Affairs, 810 Vermont Avenue NW, Washington, DC 20420
- Jennifer Moser, Ph.D., Associate Director, Scientific Programs
  US Department of Veterans Affairs, 810 Vermont Avenue NW, Washington, DC 20420
- Jennifer E. Deen, B.S., Associate Director, Cohort & Public Relations
  US Department of Veterans Affairs, 810 Vermont Avenue NW, Washington, DC 20420

*MVP Executive Committee*
- Co-Chair: Philip S. Tsao, Ph.D.
  VA Palo Alto Health Care System, 3801 Miranda Avenue, Palo Alto, CA 94304
- Co-Chair: Sumitra Muralidhar, Ph.D.
  US Department of Veterans Affairs, 810 Vermont Avenue NW, Washington, DC 20420
- J. Michael Gaziano, M.D., M.P.H.
  VA Boston Healthcare System, 150 S. Huntington Avenue, Boston, MA 02130
- Elizabeth Hauser, Ph.D.
  Durham VA Medical Center, 508 Fulton Street, Durham, NC 27705
- Amy Kilbourne, Ph.D., M.P.H.
  VA HSR&D, 2215 Fuller Road, Ann Arbor, MI 48105
- Shiuh-Wen Luoh, M.D., Ph.D.
  VA Portland Health Care System, 3710 SW US Veterans Hospital Rd, Portland, OR 97239
- Michael Matheny, M.D., M.S., M.P.H.
  VA Tennessee Valley Healthcare System, 1310 24$^{th}$ Ave. South, Nashville, TN 37212
- Dave Oslin, M.D.
  Philadelphia VA Medical Center, 3900 Woodland Avenue, Philadelphia, PA 19104

*MVP Co-Principal Investigators*
- J. Michael Gaziano, M.D., M.P.H.
  VA Boston Healthcare System, 150 S. Huntington Avenue, Boston, MA 02130
- Philip S. Tsao, Ph.D.
  VA Palo Alto Health Care System, 3801 Miranda Avenue, Palo Alto, CA 94304

*MVP Core Operations*
- Lori Churby, B.S., Director, MVP Regulatory Affairs
  VA Palo Alto Health Care System, 3801 Miranda Avenue, Palo Alto, CA 94304
- Stacey B. Whitbourne, Ph.D., Director, MVP Cohort Management
  VA Boston Healthcare System, 150 S. Huntington Avenue, Boston, MA 02130
- Jessica V. Brewer, M.P.H., Director, MVP Recruitment & Enrollment
  VA Boston Healthcare System, 150 S. Huntington Avenue, Boston, MA 02130
- Shahpoor (Alex) Shayan, M.S., Director, MVP Recruitment and Enrollment Informatics
  VA Boston Healthcare System, 150 S. Huntington Avenue, Boston, MA 02130
- Luis E. Selva, Ph.D., Executive Director, MVP Biorepositories
  VA Boston Healthcare System, 150 S. Huntington Avenue, Boston, MA 02130
- Saiju Pyarajan Ph.D., Director, Data and Computational Sciences
  VA Boston Healthcare System, 150 S. Huntington Avenue, Boston, MA 02130
- Kelly Cho, M.P.H, Ph.D., Director, MVP Phenomics Data Core
  VA Boston Healthcare System, 150 S. Huntington Avenue, Boston, MA 02130
- Scott L. DuVall, Ph.D., Director, VA Informatics and Computing Infrastructure (VINCI)
  VA Salt Lake City Health Care System, 500 Foothill Drive, Salt Lake City, UT 84148
- Mary T. Brophy M.D., M.P.H., Director, VA Central Biorepository
  VA Boston Healthcare System, 150 S. Huntington Avenue, Boston, MA 02130

- MVP Coordinating Centers
  - MVP Coordinating Center, Boston - J. Michael Gaziano, M.D., M.P.H.
    VA Boston Healthcare System, 150 S. Huntington Avenue, Boston, MA 02130
  - MVP Coordinating Center, Palo Alto – Philip S. Tsao, Ph.D.
    VA Palo Alto Health Care System, 3801 Miranda Avenue, Palo Alto, CA 94304
  - MVP Information Center, Canandaigua – Brady Stephens, M.S.
    Canandaigua VA Medical Center, 400 Fort Hill Avenue, Canandaigua, NY 14424
  - Cooperative Studies Program Clinical Research Pharmacy Coordinating Center, Albuquerque – Todd Connor, Pharm.D.; Dean P. Argyres, B.S., M.S.
    New Mexico VA Health Care System, 1501 San Pedro Drive SE, Albuquerque, NM 87108

*MVP Publications and Presentations Committee*

- Co-Chair: Tim Assimes, M.D.
  VA Palo Alto Health Care System, 3801 Miranda Avenue, Palo Alto, CA 94304
- Co-Chair: Adriana Hung, M.D.
  VA Tennessee Valley Healthcare System, 1310 24th Ave. South, Nashville, TN 37212
- Co-Chair: Henry Kranzler, M.D.
  Philadelphia VA Medical Center, 3900 Woodland Avenue, Philadelphia, PA 19104

*MVP Local Site Investigators*

- Samuel Aguayo, M.D., Phoenix VA Health Care System
  650 E. Indian School Road, Phoenix, AZ 85012
- Sunil Ahuja, M.D., South Texas Veterans Health Care System
  7400 Merton Minter Boulevard, San Antonio, TX 78229
- Kathrina Alexander, M.D., Veterans Health Care System of the Ozarks
  1100 North College Avenue, Fayetteville, AR 72703
- Xiao M. Androulakis, M.D., Columbia VA Health Care System
  6439 Garners Ferry Road, Columbia, SC 29209
- Prakash Balasubramanian, M.D., William S. Middleton Memorial Veterans Hospital
  2500 Overlook Terrace, Madison, WI 53705
- Zuhair Ballas, M.D., Iowa City VA Health Care System
  601 Highway 6 West, Iowa City, IA 52246-2208
- Jean Beckham, Ph.D., Durham VA Medical Center
  508 Fulton Street, Durham, NC 27705
- Sujata Bhushan, M.D., VA North Texas Health Care System
  4500 S. Lancaster Road, Dallas, TX 75216
- Edward Boyko, M.D., VA Puget Sound Health Care System
  1660 S. Columbian Way, Seattle, WA 98108-1597
- David Cohen, M.D., Portland VA Medical Center
  3710 SW U.S. Veterans Hospital Road, Portland, OR 97239
- Louis Dellitalia, M.D., Birmingham VA Medical Center
  700 S. 19th Street, Birmingham AL 35233
- L. Christine Faulk, M.D., Robert J. Dole VA Medical Center
  5500 East Kellogg Drive, Wichita, KS 67218-1607
- Joseph Fayad, M.D., VA Southern Nevada Healthcare System
  6900 North Pecos Road, North Las Vegas, NV 89086
- Daryl Fujii, Ph.D., VA Pacific Islands Health Care System
  459 Patterson Rd, Honolulu, HI 96819

- Saib Gappy, M.D., John D. Dingell VA Medical Center
  4646 John R Street, Detroit, MI 48201
- Frank Gesek, Ph.D., White River Junction VA Medical Center
  163 Veterans Drive, White River Junction, VT 05009
- Jennifer Greco, M.D., Sioux Falls VA Health Care System
  2501 W 22nd Street, Sioux Falls, SD 57105
- Michael Godschalk, M.D., Richmond VA Medical Center
  1201 Broad Rock Blvd., Richmond, VA 23249
- Todd W. Gress, M.D., Ph.D., Hershel "Woody" Williams VA Medical Center
  1540 Spring Valley Drive, Huntington, WV 25704
- Samir Gupta, M.D., M.S.C.S., VA San Diego Healthcare System
  3350 La Jolla Village Drive, San Diego, CA 92161
- Salvador Gutierrez, M.D., Edward Hines, Jr. VA Medical Center
  5000 South 5th Avenue, Hines, IL 60141
- John Harley, M.D., Ph.D., Cincinnati VA Medical Center
  3200 Vine Street, Cincinnati, OH 45220
- Kimberly Hammer, Ph.D., Fargo VA Health Care System
  2101 N. Elm, Fargo, ND 58102
- Mark Hamner, M.D., Ralph H. Johnson VA Medical Center
  109 Bee Street, Mental Health Research, Charleston, SC 29401
- Adriana Hung, M.D., M.P.H., VA Tennessee Valley Healthcare System
  1310 24th Avenue, South Nashville, TN 37212
- Robin Hurley, M.D., W.G. (Bill) Hefner VA Medical Center
  1601 Brenner Ave, Salisbury, NC 28144
- Pran Iruvanti, D.O., Ph.D., Hampton VA Medical Center
  100 Emancipation Drive, Hampton, VA 23667
- Frank Jacono, M.D., VA Northeast Ohio Healthcare System
  10701 East Boulevard, Cleveland, OH 44106
- Darshana Jhala, M.D., Philadelphia VA Medical Center
  3900 Woodland Avenue, Philadelphia, PA 19104
- Scott Kinlay, M.B.B.S., Ph.D., VA Boston Healthcare System
  150 S. Huntington Avenue, Boston, MA 02130
- Jon Klein, M.D., Ph.D., Louisville VA Medical Center
  800 Zorn Avenue, Louisville, KY 40206
- Michael Landry, Ph.D., Southeast Louisiana Veterans Health Care System
  2400 Canal Street, New Orleans, LA 70119
- Peter Liang, M.D., M.P.H., VA New York Harbor Healthcare System
  423 East 23rd Street, New York, NY 10010
- Suthat Liangpunsakul, M.D., M.P.H., Richard Roudebush VA Medical Center
  1481 West 10th Street, Indianapolis, IN 46202
- Jack Lichy, M.D., Ph.D., Washington DC VA Medical Center
  50 Irving St, Washington, D. C. 20422
- C. Scott Mahan, M.D., Charles George VA Medical Center
  1100 Tunnel Road, Asheville, NC 28805
- Ronnie Marrache, M.D., VA Maine Healthcare System
  1 VA Center, Augusta, ME 04330

- Stephen Mastorides, M.D., James A. Haley Veterans' Hospital
  13000 Bruce B. Downs Blvd, Tampa, FL 33612
- Elisabeth Mates M.D., Ph.D., VA Sierra Nevada Health Care System
  975 Kirman Avenue, Reno, NV 89502
- Kristin Mattocks, Ph.D., M.P.H., Central Western Massachusetts Healthcare System
  421 North Main Street, Leeds, MA 01053
- Paul Meyer, M.D., Ph.D., Southern Arizona VA Health Care System
  3601 S 6th Avenue, Tucson, AZ 85723
- Jonathan Moorman, M.D., Ph.D., James H. Quillen VA Medical Center
  Corner of Lamont & Veterans Way, Mountain Home, TN 37684
- Timothy Morgan, M.D., VA Long Beach Healthcare System
  5901 East 7th Street Long Beach, CA 90822
- Maureen Murdoch, M.D., M.P.H., Minneapolis VA Health Care System
  One Veterans Drive, Minneapolis, MN 55417
- James Norton, Ph.D., VA Health Care Upstate New York
  113 Holland Avenue, Albany, NY 12208
- Olaoluwa Okusaga, M.D., Michael E. DeBakey VA Medical Center
  2002 Holcombe Blvd, Houston, TX 77030
- Kris Ann Oursler, M.D., Salem VA Medical Center
  1970 Roanoke Blvd, Salem, VA 24153
- Ana Palacio, M.D., M.P.H., Miami VA Health Care System
  1201 NW 16th Street, 11 GRC, Miami FL 33125
- Samuel Poon, M.D., Manchester VA Medical Center
  718 Smyth Road, Manchester, NH 03104
- Emily Potter, Pharm.D., VA Eastern Kansas Health Care System
  4101 S 4th Street Trafficway, Leavenworth, KS 66048
- Michael Rauchman, M.D., St. Louis VA Health Care System
  915 North Grand Blvd, St. Louis, MO 63106
- Richard Servatius, Ph.D., Syracuse VA Medical Center
  800 Irving Avenue, Syracuse, NY 13210
- Satish Sharma, M.D., Providence VA Medical Center
  830 Chalkstone Avenue, Providence, RI 02908
- River Smith, Ph.D., Eastern Oklahoma VA Health Care System
  1011 Honor Heights Drive, Muskogee, OK 74401
- Peruvemba Sriram, M.D., N. FL/S. GA Veterans Health System
  1601 SW Archer Road, Gainesville, FL 32608
- Patrick Strollo, Jr., M.D., VA Pittsburgh Health Care System
  University Drive, Pittsburgh, PA 15240
- Neeraj Tandon, M.D., Overton Brooks VA Medical Center
  510 East Stoner Ave, Shreveport, LA 71101
- Philip Tsao, Ph.D., VA Palo Alto Health Care System
  3801 Miranda Avenue, Palo Alto, CA 94304-1290
- Gerardo Villareal, M.D., New Mexico VA Health Care System
  1501 San Pedro Drive, S.E. Albuquerque, NM 87108
- Agnes Wallbom, M.D., M.S., VA Greater Los Angeles Health Care System
  11301 Wilshire Blvd, Los Angeles, CA 90073

- Jessica Walsh, M.D., VA Salt Lake City Health Care System
  500 Foothill Drive, Salt Lake City, UT 84148
- John Wells, Ph.D., Edith Nourse Rogers Memorial Veterans Hospital
  200 Springs Road, Bedford, MA 01730
- Jeffrey Whittle, M.D., M.P.H., Clement J. Zablocki VA Medical Center
  5000 West National Avenue, Milwaukee, WI 53295
- Mary Whooley, M.D., San Francisco VA Health Care System
  4150 Clement Street, San Francisco, CA 94121
- Allison E. Williams, N.D., Ph.D., R.N, Bay Pines VA Healthcare System
  10,000 Bay Pines Blvd Bay Pines, FL 33744
- Peter Wilson, M.D., Atlanta VA Medical Center
  1670 Clairmont Road, Decatur, GA 30033
- Junzhe Xu, M.D., VA Western New York Healthcare System
  3495 Bailey Avenue, Buffalo, NY 14215-1199
- Shing Shing Yeh, Ph.D., M.D., Northport VA Medical Center
  79 Middleville Road, Northport, NY 11768

Supplementary Note 4. **NHLBI TOPMED: NHLBI TRANS-OMICS FOR PRECISION MEDICINE (TOPMED) CONSORTIUM BANNER AUTHORSHIP**

***Banner Authors (in alphabetical order by last name)***

Abe, Namiko, New York Genome Center, New York, New York, 10013, US; Abecasis, Gonçalo, University of Michigan, Ann Arbor, Michigan, 48109, US; Aguet, Francois, Broad Institute, Cambridge, Massachusetts, 02142, US; Albert, Christine, Cedars Sinai, Boston, Massachusetts, 02114, US; Almasy, Laura, Children's Hospital of Philadelphia, University of Pennsylvania, Philadelphia, Pennsylvania, 19104, US; Alonso, Alvaro, Emory University, Atlanta, Georgia, 30322, US; Ament, Seth, University of Maryland, Baltimore, Maryland, 21201, US; Anderson, Peter, University of Washington, Seattle, Washington, 98195, US; Anugu, Pramod, University of Mississippi, Jackson, Mississippi, 38677, US; Applebaum-Bowden, Deborah, National Institutes of Health, Bethesda, Maryland, 20892, US; Ardlie, Kristin, Broad Institute, Cambridge, Massachusetts, 02142, US; Arking, Dan, Johns Hopkins University, Baltimore, Maryland, 21218, US; Arnett, Donna K, University of Kentucky, Lexington, Kentucky, 40506, US; Ashley-Koch, Allison, Duke University, Durham, North Carolina, 27708, US; Aslibekyan, Stella, University of Alabama, Birmingham, Alabama, 35487, US; Assimes, Tim, Stanford University, Stanford, California, 94305, US; Auer, Paul, Medical College of Wisconsin, Milwaukee, Wisconsin, 53211, US; Avramopoulos, Dimitrios, Johns Hopkins University, Baltimore, Maryland, 21218, US; Ayas, Najib, Providence Health Care, Medicine, Vancouver, CA; Balasubramanian, Adithya, Baylor College of Medicine Human Genome Sequencing Center, Houston, Texas, 77030, US; Barnard, John, Cleveland Clinic, Cleveland, Ohio, 44195, US; Barnes, Kathleen, Tempus, University of Colorado Anschutz Medical Campus, Aurora, Colorado, 80045, US; Barr, R. Graham, Columbia University, New York, New York, 10032, US; Barron-Casella, Emily, Johns Hopkins University, Baltimore, Maryland, 21218, US; Barwick, Lucas, The Emmes Corporation, LTRC, Rockville, Maryland, 20850, US; Beaty, Terri, Johns Hopkins University, Baltimore, Maryland, 21218, US; Beck, Gerald, Cleveland Clinic, Quantitative Health Sciences, Cleveland, Ohio, 44195, US; Becker, Diane, Johns Hopkins University, Medicine, Baltimore, Maryland, 21218, US; Becker, Lewis, Johns Hopkins University, Baltimore, Maryland, 21218, US; Beer, Rebecca, National Heart, Lung, and Blood Institute, National Institutes of Health, Bethesda, Maryland, 20892, US; Beitelshees, Amber, University of Maryland, Baltimore, Maryland, 21201, US; Benjamin, Emelia, Boston University, Massachusetts General Hospital, Boston University School of Medicine, Boston, Massachusetts, 02114, US; Benos, Takis, University of Pittsburgh, Pittsburgh, Pennsylvania, 15260, US; Bezerra, Marcos, Fundação de Hematologia e Hemoterapia de Pernambuco -

Hemope, Recife, 52011-000, BR; Bielak, Larry, University of Michigan, Ann Arbor, Michigan, 48109, US; Bis, Joshua, University of Washington, Cardiovascular Health Research Unit, Department of Medicine, Seattle, Washington, 98195, US; Blackwell, Thomas, University of Michigan, Ann Arbor, Michigan, 48109, US; Blangero, John, University of Texas Rio Grande Valley School of Medicine, Human Genetics, Brownsville, Texas, 78520, US; Boerwinkle, Eric, University of Texas Health at Houston, Houston, Texas, 77225, US; Bowden, Donald W., Wake Forest Baptist Health, Department of Biochemistry, Winston-Salem, North Carolina, 27157, US; Bowler, Russell, National Jewish Health, National Jewish Health, Denver, Colorado, 80206, US; Brody, Jennifer, University of Washington, Seattle, Washington, 98195, US; Broeckel, Ulrich, Medical College of Wisconsin, Pediatrics, Milwaukee, Wisconsin, 53226, US; Broome, Jai, University of Washington, Seattle, Washington, 98195, US; Brown, Deborah, University of Texas Health at Houston, Pediatrics, Houston, Texas, 77030, US; Bunting, Karen, New York Genome Center, New York, New York, 10013, US; Burchard, Esteban, University of California, San Francisco, San Francisco, California, 94143, US; Bustamante, Carlos, Stanford University, Biomedical Data Science, Stanford, California, 94305, US; Buth, Erin, University of Washington, Biostatistics, Seattle, Washington, 98195, US; Cade, Brian, Brigham & Women's Hospital, Brigham and Women's Hospital, Boston, Massachusetts, 02115, US; Cardwell, Jonathan, University of Colorado at Denver, Denver, Colorado, 80204, US; Carey, Vincent, Brigham & Women's Hospital, Boston, Massachusetts, 02115, US; Carrier, Julie, University of Montreal, , US; Carson, April, University of Mississippi, Medicine, Jackson, Mississippi, 39213, US; Carty, Cara, Washington State University, Pullman, Washington, 99164, US; Casaburi, Richard, University of California, Los Angeles, Los Angeles, California, 90095, US; Casas Romero, Juan P, Brigham & Women's Hospital, , US; Casella, James, Johns Hopkins University, Baltimore, Maryland, 21218, US; Castaldi, Peter, Brigham & Women's Hospital, Medicine, Boston, Massachusetts, 02115, US; Chaffin, Mark, Broad Institute, Cambridge, Massachusetts, 02142, US; Chang, Christy, University of Maryland, Baltimore, Maryland, 21201, US; Chang, Yi-Cheng, National Taiwan University, Taipei, 10617, TW; Chasman, Daniel, Brigham & Women's Hospital, Division of Preventive Medicine, Boston, Massachusetts, 02215, US; Chavan, Sameer, University of Colorado at Denver, Denver, Colorado, 80204, US; Chen, Bo-Juen, New York Genome Center, New York, New York, 10013, US; Chen, Wei-Min, University of Virginia, Charlottesville, Virginia, 22903, US; Chen, Yii-Der Ida, Lundquist Institute, Torrance, California, 90502, US; Cho, Michael, Brigham & Women's Hospital, Boston, Massachusetts, 02115, US; Choi, Seung Hoan, Broad Institute, Cambridge, Massachusetts, 02142, US; Chuang, Lee-Ming, National Taiwan University, National Taiwan University Hospital, Taipei, 10617, TW; Chung, Mina, Cleveland Clinic, Cleveland Clinic, Cleveland, Ohio, 44195, US; Chung, Ren-Hua, National Health Research Institute Taiwan, Miaoli County, 350, TW; Clish, Clary, Broad Institute, Metabolomics Platform, Cambridge, Massachusetts, 02142, US; Comhair, Suzy, Cleveland Clinic, Immunity and Immunology, Cleveland, Ohio, 44195, US; Conomos, Matthew, University of Washington, Biostatistics, Seattle, Washington, 98195, US; Cornell, Elaine, University of Vermont, Burlington, Vermont, 05405, US; Correa, Adolfo, University of Mississippi, Population Health Science, Jackson, Mississippi, 39216, US; Crandall, Carolyn, University of California, Los Angeles, Los Angeles, California, 90095, US; Crapo, James, National Jewish Health, Denver, Colorado, 80206, US; Cupples, L. Adrienne, Boston University, Biostatistics, Boston, Massachusetts, 02115, US; Curran, Joanne, University of Texas Rio Grande Valley School of Medicine, Brownsville, Texas, 78520, US; Curtis, Jeffrey, University of Michigan, Internal Medicine, Ann Arbor, Michigan, 48109, US; Custer, Brian, Vitalant Research Institute, San Francisco, California, 94118, US; Damcott, Coleen, University of Maryland, Baltimore, Maryland, 21201, US; Darbar, Dawood, University of Illinois at Chicago, Chicago, Illinois, 60607, US; David, Sean, University of Chicago, Chicago, Illinois, 60637, US; Davis, Colleen, University of Washington, Seattle, Washington, 98195, US; Daya, Michelle, University of Colorado at Denver, Denver, Colorado, 80204, US; de Andrade, Mariza, Mayo Clinic, Health Quantitative Sciences Research , Rochester, Minnesota, 55905, US; de las Fuentes, Lisa, Washington University in St Louis, Department of Medicine, Cardiovascular Division, St. Louis, Missouri, 63110, US; de Vries, Paul, University of Texas Health at Houston, Human Genetics Center, Department of Epidemiology, Human Genetics, and Environmental Sciences, Houston, Texas, 77030,

US; DeBaun, Michael, Vanderbilt University, Nashville, Tennessee, 37235, US; Deka, Ranjan, University of Cincinnati, Cincinnati, Ohio, 45220, US; DeMeo, Dawn, Brigham & Women's Hospital, Boston, Massachusetts, 02115, US; Devine, Scott, University of Maryland, Baltimore, Maryland, 21201, US; Dinh, Huyen, Baylor College of Medicine Human Genome Sequencing Center, Houston, Texas, 77030, US; Doddapaneni, Harsha , Baylor College of Medicine Human Genome Sequencing Center, Houston, Texas, 77030, ; Duan, Qing, University of North Carolina, Chapel Hill, North Carolina, 27599, US; Dugan-Perez, Shannon, Baylor College of Medicine Human Genome Sequencing Center, Houston, Texas, 77030, US; Duggirala, Ravi, University of Texas Rio Grande Valley School of Medicine, Edinburg, Texas, 78539, US; Durda, Jon Peter, University of Vermont, Burlington, Vermont, 05405, US; Dutcher, Susan K., Washington University in St Louis, Genetics , St Louis, Missouri, 63110, US; Eaton, Charles, Brown University, Providence, Rhode Island, 02912, US; Ekunwe, Lynette, University of Mississippi, Jackson, Mississippi, 38677, US; El Boueiz, Adel, Harvard University, Channing Division of Network Medicine, Cambridge, Massachusetts, 02138, US; Ellinor, Patrick, Massachusetts General Hospital, Boston, Massachusetts, 02114, US; Emery, Leslie, University of Washington, Seattle, Washington, 98195, US; Erzurum, Serpil, Cleveland Clinic, Cleveland, Ohio, 44195, US; Farber, Charles, University of Virginia, Charlottesville, Virginia, 22903, US; Farek, Jesse, Baylor College of Medicine Human Genome Sequencing Center, Houston, Texas, 77030, US; Fingerlin, Tasha, National Jewish Health, Center for Genes, Environment and Health, Denver, Colorado, 80206, US; Flickinger, Matthew, University of Michigan, Ann Arbor, Michigan, 48109, US; Fornage, Myriam, University of Texas Health at Houston, Houston, Texas, 77225, US; Franceschini, Nora, University of North Carolina, Epidemiology, Chapel Hill, North Carolina, 27599, US; Frazar, Chris, University of Washington, Seattle, Washington, 98195, US; Fu, Mao, University of Maryland, Baltimore, Maryland, 21201, US; Fullerton, Stephanie M., University of Washington, Seattle, Washington, 98195, US; Fulton, Lucinda, Washington University in St Louis, St Louis, Missouri, 63130, US; Gabriel, Stacey, Broad Institute, Cambridge, Massachusetts, 02142, US; Gan, Weiniu, National Heart, Lung, and Blood Institute, National Institutes of Health, Bethesda, Maryland, 20892, US; Gao, Shanshan, University of Colorado at Denver, Denver, Colorado, 80204, US; Gao, Yan, University of Mississippi, Jackson, Mississippi, 38677, US; Gass, Margery, Fred Hutchinson Cancer Research Center, Seattle, Washington, 98109, US; Geiger, Heather, New York Genome Center, New York City, New York, 10013, US; Gelb, Bruce, Icahn School of Medicine at Mount Sinai, New York, New York, 10029, US; Geraci, Mark, University of Pittsburgh, Pittsburgh, Pennsylvania, US; Germer, Soren, New York Genome Center, New York, New York, 10013, US; Gerszten, Robert, Beth Israel Deaconess Medical Center, Boston, Massachusetts, 02215, US; Ghosh, Auyon, Brigham & Women's Hospital, Boston, Massachusetts, 02115, US; Gibbs, Richard, Baylor College of Medicine Human Genome Sequencing Center, Houston, Texas, 77030, US; Gignoux, Chris, Stanford University, Stanford, California, 94305, US; Gladwin, Mark, University of Pittsburgh, Pittsburgh, Pennsylvania, 15260, US; Glahn, David, Boston Children's Hospital, Harvard Medical School, Department of Psychiatry, Boston, Massachusetts, 02115, US; Gogarten, Stephanie, University of Washington, Seattle, Washington, 98195, US; Gong, Da-Wei, University of Maryland, Baltimore, Maryland, 21201, US; Goring, Harald, University of Texas Rio Grande Valley School of Medicine, San Antonio, Texas, 78229, US; Graw, Sharon, University of Colorado Anschutz Medical Campus, Aurora, Colorado, 80045, US; Gray, Kathryn J., Mass General Brigham, Obstetrics and Gynecology, Boston, Massachusetts, 02115, US; Grine, Daniel, University of Colorado at Denver, Denver, Colorado, 80204, US; Gross, Colin, University of Michigan, Ann Arbor, Michigan, 48109, US; Gu, C. Charles, Washington University in St Louis, St Louis, Missouri, 63130, US; Guan, Yue, University of Maryland, Baltimore, Maryland, 21201, US; Guo, Xiuqing, Lundquist Institute, Torrance, California, 90502, US; Gupta, Namrata, Broad Institute, Cambridge, Massachusetts, 02142, US; Haas, David M., Indiana University, OB/GYN, Indianapolis, Indiana, 46202, US; Haessler, Jeff, Fred Hutchinson Cancer Research Center, Seattle, Washington, 98109, US; Hall, Michael, University of Mississippi, Cardiology, Jackson, Mississippi, 39216, US; Han, Yi, Baylor College of Medicine Human Genome Sequencing Center, Houston, Texas, 77030, US; Hanly, Patrick, University of Calgary, Medicine, Calgary, CA; Harris, Daniel, University of Maryland, Genetics, Philadelphia, Pennsylvania, 19104, US; Hawley, Nicola L.,

Yale University, Department of Chronic Disease Epidemiology, New Haven, Connecticut, 06520, US; He, Jiang, Tulane University, New Orleans, Louisiana, 70118, US; Heavner, Ben, University of Washington, Biostatistics, Seattle, Washington, 98195, US; Heckbert, Susan, University of Washington, Epidemiology, Seattle, Washington, 98195, US; Hernandez, Ryan, University of California, San Francisco, San Francisco, California, 94143, US; Herrington, David, Wake Forest Baptist Health, Winston-Salem, North Carolina, 27157, US; Hersh, Craig, Brigham & Women's Hospital, Channing Division of Network Medicine, Boston, Massachusetts, 02115, US; Hidalgo, Bertha, University of Alabama, Birmingham, Alabama, 35487, US; Hixson, James, University of Texas Health at Houston, Houston, Texas, 77225, US; Hobbs, Brian, Brigham & Women's Hospital, Boston, Massachusetts, 02115, US; Hokanson, John, University of Colorado at Denver, Denver, Colorado, 80204, US; Hong, Elliott, University of Maryland, Baltimore, Maryland, 21201, US; Hoth, Karin, University of Iowa, Iowa City, Iowa, 52242, US; Hsiung, Chao (Agnes), National Health Research Institute Taiwan, Institute of Population Health Sciences, NHRI, Miaoli County, 350, TW; Hu, Jianhong, Baylor College of Medicine Human Genome Sequencing Center, Houston, Texas, 77030, US; Hung, Yi-Jen, Tri-Service General Hospital National Defense Medical Center, , TW; Huston, Haley, Blood Works Northwest, Seattle, Washington, 98104, US; Hwu, Chii Min, Taichung Veterans General Hospital Taiwan, Taichung City, 407, TW; Irvin, Marguerite Ryan, University of Alabama, Birmingham, Alabama, 35487, US; Jackson, Rebecca, Oklahoma State University Medical Center, Internal Medicine, DIvision of Endocrinology, Diabetes and Metabolism, Columbus, Ohio, 43210, US; Jain, Deepti, University of Washington, Seattle, Washington, 98195, US; Jaquish, Cashell, National Heart, Lung, and Blood Institute, National Institutes of Health, NHLBI, Bethesda, Maryland, 20892, US; Johnsen, Jill, Blood Works Northwest, Research Institute, Seattle, Washington, 98104, US; Johnson, Andrew, National Heart, Lung, and Blood Institute, National Institutes of Health, Bethesda, Maryland, 20892, US; Johnson, Craig, University of Washington, Seattle, Washington, 98195, US; Johnston, Rich, Emory University, Atlanta, Georgia, 30322, US; Jones, Kimberly, Johns Hopkins University, Baltimore, Maryland, 21218, US; Kang, Hyun Min, University of Michigan, Biostatistics, Ann Arbor, Michigan, 48109, US; Kaplan, Robert, Albert Einstein College of Medicine, New York, New York, 10461, US; Kardia, Sharon, University of Michigan, Ann Arbor, Michigan, 48109, US; Kelly, Shannon, University of California, San Francisco, San Francisco, California, 94118, US; Kenny, Eimear, Icahn School of Medicine at Mount Sinai, New York, New York, 10029, US; Kessler, Michael, University of Maryland, Baltimore, Maryland, 21201, US; Khan, Alyna, University of Washington, Seattle, Washington, 98195, US; Khan, Ziad, Baylor College of Medicine Human Genome Sequencing Center, Houston, Texas, 77030, US; Kim, Wonji, Harvard University, Cambridge, Massachusetts, 02138, US; Kimoff, John, McGill University, Montréal, QC H3A 0G4, CA; Kinney, Greg, University of Colorado at Denver, Epidemiology, Aurora, Colorado, 80045, US; Konkle, Barbara, Blood Works Northwest, Medicine, Seattle, Washington, 98104, US; Kooperberg, Charles, Fred Hutchinson Cancer Research Center, Seattle, Washington, 98109, US; Kramer, Holly, Loyola University, Public Health Sciences, Maywood, Illinois, 60153, US; Lange, Christoph, Harvard School of Public Health, Biostats, Boston, Massachusetts, 02115, US; Lange, Ethan, University of Colorado at Denver, Denver, Colorado, 80204, US; Lange, Leslie, University of Colorado at Denver, Medicine, Aurora, Colorado, 80048, US; Laurie, Cathy, University of Washington, Seattle, Washington, 98195, US; Laurie, Cecelia, University of Washington, Seattle, Washington, 98195, US; LeBoff, Meryl, Brigham & Women's Hospital, Boston, Massachusetts, 02115, US; Lee, Jiwon, Brigham & Women's Hospital, Boston, Massachusetts, 02115, US; Lee, Sandra, Baylor College of Medicine Human Genome Sequencing Center, Houston, Texas, 77030, US; Lee, Wen-Jane, Taichung Veterans General Hospital Taiwan, Taichung City, 407, TW; LeFaive, Jonathon, University of Michigan, Ann Arbor, Michigan, 48109, US; Levine, David, University of Washington, Seattle, Washington, 98195, US; Levy, Dan, National Heart, Lung, and Blood Institute, National Institutes of Health, Bethesda, Maryland, 20892, US; Lewis, Joshua, University of Maryland, Baltimore, Maryland, 21201, US; Li, Xiaohui, Lundquist Institute, Torrance, California, 90502, US; Li, Yun, University of North Carolina, Chapel Hill, North Carolina, 27599, US; Lin, Henry, Lundquist Institute, Torrance, California, 90502, US; Lin, Honghuang, Boston University, Boston, Massachusetts, 02215, US; Lin, Xihong, Harvard School of Public Health,

Boston, Massachusetts, 02115, US; Liu, Simin, Brown University, Epidemiology and Medicine, Providence, Rhode Island, 02912, US; Liu, Yongmei, Duke University, Cardiology, Durham, North Carolina, 27708, US; Liu, Yu, Stanford University, Cardiovascular Institute, Stanford, California, 94305, US; Loos, Ruth J.F., Icahn School of Medicine at Mount Sinai, The Charles Bronfman Institute for Personalized Medicine, New York, New York, 10029, US; Lubitz, Steven, Massachusetts General Hospital, Boston, Massachusetts, 02114, US; Lunetta, Kathryn, Boston University, Boston, Massachusetts, 02215, US; Luo, James, National Heart, Lung, and Blood Institute, National Institutes of Health, Bethesda, Maryland, 20892, US; Magalang, Ulysses, Ohio State University, Division of Pulmonary, Critical Care and Sleep Medicine, Columbus, Ohio, 43210, US; Mahaney, Michael, University of Texas Rio Grande Valley School of Medicine, Brownsville, Texas, 78520, US; Make, Barry, Johns Hopkins University, Baltimore, Maryland, 21218, US; Manichaikul, Ani, University of Virginia, Charlottesville, Virginia, 22903, US; Manning, Alisa, Broad Institute, Harvard University, Massachusetts General Hospital, , ; Manson, JoAnn, Brigham & Women's Hospital, Boston, Massachusetts, 02115, US; Martin, Lisa, George Washington University, cardiology, Washington, District of Columbia, 20037, US; Marton, Melissa, New York Genome Center, New York City, New York, 10013, US; Mathai, Susan, University of Colorado at Denver, Denver, Colorado, 80204, US; Mathias, Rasika, Johns Hopkins University, Baltimore, Maryland, 21218, US; May, Susanne, University of Washington, Biostatistics, Seattle, Washington, 98195, US; McArdle, Patrick, University of Maryland, Baltimore, Maryland, 21201, US; McDonald, Merry-Lynn, University of Alabama, University of Alabama at Birmingham, Birmingham, Alabama, 35487, US; McFarland, Sean, Harvard University, Cambridge, Massachusetts, 02138, US; McGarvey, Stephen, Brown University, Epidemiology, Providence, Rhode Island, 02912, US; McGoldrick, Daniel , University of Washington, Genome Sciences, Seattle, Washington, 98195, US; McHugh, Caitlin, University of Washington, Biostatistics, Seattle, Washington, 98195, US; McNeil, Becky, RTI International, , US; Mei, Hao, University of Mississippi, Jackson, Mississippi, 38677, US; Meigs, James, Massachusetts General Hospital, Medicine, Boston , Massachusetts, 02114, US; Menon, Vipin, Baylor College of Medicine Human Genome Sequencing Center, Houston, Texas, 77030, US; Mestroni, Luisa, University of Colorado Anschutz Medical Campus, Aurora, Colorado, 80045, US; Metcalf, Ginger, Baylor College of Medicine Human Genome Sequencing Center, Houston, Texas, 77030, US; Meyers, Deborah A, University of Arizona, Tucson, Arizona, 85721, US; Mignot, Emmanuel, Stanford University, Center For Sleep Sciences and Medicine, Palo Alto, California, 94304, US; Mikulla, Julie, National Heart, Lung, and Blood Institute, National Institutes of Health, Bethesda, Maryland, 20892, US; Min, Nancy, University of Mississippi, Jackson, Mississippi, 38677, US; Minear, Mollie, National Institute of Child Health and Human Development, National Institutes of Health, Bethesda, Maryland, 20892, US; Minster, Ryan L, University of Pittsburgh, Pittsburgh, Pennsylvania, 15260, US; Mitchell, Braxton D., University of Maryland, Baltimore, Maryland, 21201, US; Moll, Matt, Brigham & Women's Hospital, Medicine, Boston, Massachusetts, 02115, US; Momin, Zeineen, Baylor College of Medicine Human Genome Sequencing Center, Houston, Texas, 77030, US; Montasser, May E., University of Maryland, Baltimore, Maryland, 21201, US; Montgomery, Courtney, Oklahoma Medical Research Foundation, Genes and Human Disease, Oklahoma City, Oklahoma, 73104, US; Muzny, Donna, Baylor College of Medicine Human Genome Sequencing Center, Houston, Texas, 77030, US; Mychaleckyj, Josyf C, University of Virginia, Charlottesville, Virginia, 22903, US; Nadkarni, Girish, Icahn School of Medicine at Mount Sinai, New York, New York, 10029, US; Naik, Rakhi, Johns Hopkins University, Baltimore, Maryland, 21218, US; Naseri, Take, Ministry of Health, Government of Samoa, Apia, WS; Natarajan, Pradeep, Broad Institute, Cambridge, Massachusetts, 02142, US; Nekhai, Sergei, Howard University, Washington, District of Columbia, 20059, US; Nelson, Sarah C., University of Washington, Biostatistics, Seattle, Washington, 98195, US; Neltner, Bonnie, University of Colorado at Denver, Denver, Colorado, 80204, US; Nessner, Caitlin, Baylor College of Medicine Human Genome Sequencing Center, Houston, Texas, 77030, US; Nickerson, Deborah, University of Washington, Department of Genome Sciences, Seattle, Washington, 98195, US; Nkechinyere, Osuji, Baylor College of Medicine Human Genome Sequencing Center, Houston, Texas, 77030, US; North, Kari, University of North Carolina, Chapel Hill, North Carolina,

27599, US; O'Connell, Jeff, University of Maryland, Balitmore, Maryland, 21201, US; O'Connor, Tim, University of Maryland, Baltimore, Maryland, 21201, US; Ochs-Balcom, Heather, University at Buffalo, Buffalo, New York, 14260, US; Okwuonu, Geoffrey, Baylor College of Medicine Human Genome Sequencing Center, Houston, Texas, 77030, US; Pack, Allan , University of Pennsylvania, Division of Sleep Medicine/Department of Medicine, Philadelphia, Pennsylvania, 19104-3403, US; Paik, David T., Stanford University, Stanford Cardiovascular Institute, Stanford, California, 94305, US; Palmer, Nicholette, Wake Forest Baptist Health, Biochemistry, Winston-Salem, North Carolina, 27157, US; Pankow, James, University of Minnesota, Minneapolis, Minnesota, 55455, US; Papanicolaou, George, National Heart, Lung, and Blood Institute, National Institutes of Health, Bethesda, Maryland, 20892, US; Parker, Cora, RTI International, Biostatistics and Epidemiology Division, Research Triangle Park, North Carolina, 27709-2194, US; Peloso, Gina, Boston University, Department of Biostatistics, Boston, Massachusetts, 02118, US; Peralta, Juan Manuel, University of Texas Rio Grande Valley School of Medicine, Edinburg, Texas, 78539, US; Perez, Marco, Stanford University, Stanford, California, 94305, US; Perry, James, University of Maryland, Baltimore, Maryland, 21201, US; Peters, Ulrike, Fred Hutchinson Cancer Research Center, Fred Hutch and UW, Seattle, Washington, 98109, US; Peyser, Patricia, University of Michigan, Ann Arbor, Michigan, 48109, US; Phillips, Lawrence S, Emory University, Atlanta, Georgia, 30322, US; Pleiness, Jacob, University of Michigan, Ann Arbor, Michigan, 48109, US; Pollin, Toni, University of Maryland, Baltimore, Maryland, 21201, US; Post, Wendy, Johns Hopkins University, Cardiology/Medicine, Baltimore, Maryland, 21218, US; Powers Becker, Julia, University of Colorado at Denver, Medicine, Denver, Colorado, 80204, US; Preethi Boorgula, Meher, University of Colorado at Denver, Denver, Colorado, 80204, US; Preuss, Michael, Icahn School of Medicine at Mount Sinai, New York, New York, 10029, US; Psaty, Bruce, University of Washington, Seattle, Washington, 98195, US; Qasba, Pankaj, National Heart, Lung, and Blood Institute, National Institutes of Health, Bethesda, Maryland, 20892, US; Qiao, Dandi, Brigham & Women's Hospital, Boston, Massachusetts, 02115, US; Qin, Zhaohui, Emory University, Atlanta, Georgia, 30322, US; Rafaels, Nicholas, University of Colorado at Denver, CCPM, Denver, Colorado, 80045, US; Raffield, Laura, University of North Carolina, Genetics, Chapel Hill, North Carolina, 27599, US; Rajendran, Mahitha, Baylor College of Medicine Human Genome Sequencing Center, Houston, Texas, 77030, US; Ramachandran, Vasan S., Boston University, Boston, Massachusetts, 02215, US; Rao, D.C., Washington University in St Louis, St Louis, Missouri, 63130, US; Rasmussen-Torvik, Laura, Northwestern University, Chicago, Illinois, 60208, US; Ratan, Aakrosh, University of Virginia, Charlottesville, Virginia, 22903, US; Redline, Susan, Brigham & Women's Hospital, Medicine, Boston, Massachusetts, 02115, US; Reed, Robert, University of Maryland, Baltimore, Maryland, 21201, US; Reeves, Catherine, New York Genome Center, New York Genome Center, New York City, New York, 10013, US; Regan, Elizabeth, National Jewish Health, Denver, Colorado, 80206, US; Reiner, Alex, Fred Hutchinson Cancer Research Center, University of Washington, Seattle, Washington, 98109, US; Reupena, Muagututi‘a Sefuiva, Lutia I Puava Ae Mapu I Fagalele, Apia, WS; Rice, Ken, University of Washington, Seattle, Washington, 98195, US; Rich, Stephen, University of Virginia, Charlottesville, Virginia, 22903, US; Robillard, Rebecca, University of Ottawa, Sleep Research Unit, University of Ottawa Institute for Mental Health Research, Ottawa, ON K1Z 7K4, CA; Robine, Nicolas, New York Genome Center, New York City, New York, 10013, US; Roden, Dan, Vanderbilt University, Medicine, Pharmacology, Biomedicla Informatics, Nashville, Tennessee, 37235, US; Roselli, Carolina, Broad Institute, Cambridge, Massachusetts, 02142, US; Rotter, Jerome, Lundquist Institute, Pediatrics, Torrance, California, 90502, US; Ruczinski, Ingo, Johns Hopkins University, Baltimore, Maryland, 21218, US; Runnels, Alexi, New York Genome Center, New York City, New York, 10013, US; Russell, Pamela, University of Colorado at Denver, Denver, Colorado, 80204, US; Ruuska, Sarah, Blood Works Northwest, Seattle, Washington, 98104, US; Ryan, Kathleen, University of Maryland, Baltimore, Maryland, 21201, US; Sabino, Ester Cerdeira, Universidade de Sao Paulo, Faculdade de Medicina, Sao Paulo, 01310000, BR; Saleheen, Danish, Columbia University, New York, New York, 10027, US; Salimi, Shabnam, University of Maryland, Pathology , Seattle, Washington, 98195, US; Salvi, Sejal, Baylor College of Medicine Human Genome Sequencing Center, Houston, Texas, 77030, US; Salzberg, Steven, Johns Hopkins University,

Baltimore, Maryland, 21218, US; Sandow, Kevin, Lundquist Institute, TGPS, Torrance, California, 90502, US; Sankaran, Vijay G., Harvard University, Division of Hematology/Oncology, Boston, Massachusetts, 02115, US; Santibanez, Jireh, Baylor College of Medicine Human Genome Sequencing Center, Houston, Texas, 77030, US; Schwander, Karen, Washington University in St Louis, St Louis, Missouri, 63130, US; Schwartz, David, University of Colorado at Denver, Denver, Colorado, 80204, US; Sciurba, Frank, University of Pittsburgh, Pittsburgh, Pennsylvania, 15260, US; Seidman, Christine , Harvard Medical School, Genetics, Boston, Massachusetts, 02115, US; Seidman, Jonathan, Harvard Medical School, Boston, Massachusetts, 02115, US; Sériès, Frédéric, Université Laval, Quebec City, G1V 0A6, CA; Sheehan, Vivien, Emory University, Pediatrics, Atlanta, Georgia, 30307, US; Sherman, Stephanie L., Emory University, Human Genetics, Atlanta, Georgia, 30322, US; Shetty, Amol, University of Maryland, Baltimore, Maryland, 21201, US; Shetty, Aniket, University of Colorado at Denver, Denver, Colorado, 80204, US; Sheu, Wayne Hui-Heng, Taichung Veterans General Hospital Taiwan, Taichung City, 407, TW; Shoemaker, M. Benjamin, Vanderbilt University, Medicine/Cardiology, Nashville, Tennessee, 37235, US; Silver, Brian, UMass Memorial Medical Center, Worcester, Massachusetts, 01655, US; Silverman, Edwin, Brigham & Women's Hospital, Boston, Massachusetts, 02115, US; Skomro, Robert, University of Saskatchewan, Saskatoon, SK S7N 5C9, CA; Smith, Albert Vernon, University of Michigan, , ; Smith, Jennifer, University of Michigan, Ann Arbor, Michigan, 48109, US; Smith, Josh, University of Washington, Seattle, Washington, 98195, US; Smith, Nicholas, University of Washington, Epidemiology, Seattle, Washington, 98195, US; Smith, Tanja, New York Genome Center, New York, New York, 10013, US; Smoller, Sylvia, Albert Einstein College of Medicine, New York, New York, 10461, US; Snively, Beverly, Wake Forest Baptist Health, Biostatistical Sciences, Winston-Salem, North Carolina, 27157, US; Snyder, Michael, Stanford University, Stanford, California, 94305, US; Sofer, Tamar, Brigham & Women's Hospital, Boston, Massachusetts, 02115, US; Sotoodehnia, Nona, University of Washington, Seattle, Washington, 98195, US; Stilp, Adrienne M., University of Washington, Seattle, Washington, 98195, US; Storm, Garrett , University of Colorado at Denver, Genomic Cardiology, Aurora, Colorado, 80045, US; Streeten, Elizabeth, University of Maryland, Baltimore, Maryland, 21201, US; Su, Jessica Lasky, Brigham & Women's Hospital, Channing Department of Medicine, Boston, Massachusetts, 02115, US; Sung, Yun Ju, Washington University in St Louis, St Louis, Missouri, 63130, US; Sylvia, Jody, Brigham & Women's Hospital, Boston, Massachusetts, 02115, US; Szpiro, Adam, University of Washington, Seattle, Washington, 98195, US; Taliun, Daniel, University of Michigan, Ann Arbor, Michigan, 48109, US; Tang, Hua, Stanford University, Genetics, Stanford, California, 94305, US; Taub, Margaret, Johns Hopkins University, Baltimore, Maryland, 21218, US; Taylor, Kent D., Lundquist Institute, Institute for Translational Genomics and Populations Sciences, Torrance, California, 90502, US; Taylor, Matthew, University of Colorado Anschutz Medical Campus, Aurora, Colorado, 80045, US; Taylor, Simeon, University of Maryland, Baltimore, Maryland, 21201, US; Telen, Marilyn, Duke University, Durham, North Carolina, 27708, US; Thornton, Timothy A., University of Washington, Seattle, Washington, 98195, US; Threlkeld, Machiko, University of Washington, University of Washington, Department of Genome Sciences, Seattle, Washington, 98195, US; Tinker, Lesley, Fred Hutchinson Cancer Research Center, Cancer Prevention Division of Public Health Sciences, Seattle, Washington, 98109, US; Tirschwell, David, University of Washington, Seattle, Washington, 98195, US; Tishkoff, Sarah, University of Pennsylvania, Genetics, Philadelphia, Pennsylvania, 19104, US; Tiwari, Hemant, University of Alabama, Biostatistics, Birmingham, Alabama, 35487, US; Tong, Catherine, University of Washington, Department of Biostatistics, Seattle, Washington, 98195, US; Tracy, Russell, University of Vermont, Pathology & Laboratory Medicine, Burlington, Vermont, 05405, US; Tsai, Michael, University of Minnesota, Minneapolis, Minnesota, 55455, US; Vaidya, Dhananjay, Johns Hopkins University, Baltimore, Maryland, 21218, US; Van Den Berg, David, University of Southern California, USC Methylation Characterization Center, University of Southern California, California, 90033, US; VandeHaar, Peter, University of Michigan, Ann Arbor, Michigan, 48109, US; Vrieze, Scott, University of Minnesota, Minneapolis, Minnesota, 55455, US; Walker, Tarik, University of Colorado at Denver, Denver, Colorado, 80204, US; Wallace, Robert, University of Iowa, Iowa City, Iowa, 52242, US; Walts, Avram, University of Colorado at Denver, Denver, Colorado, 80204, US; Wang, Fei Fei, University of Washington, Seattle, Washington, 98195, US; Wang, Heming, Brigham & Women's Hospital, Mass General Brigham, Boston, Massachusetts, 02115, US; Wang, Jiongming, University of Michigan, , US; Watson, Karol, University of California, Los Angeles, Los Angeles, California, 90095, US; Watt, Jennifer, Baylor College of Medicine Human Genome Sequencing Center, Houston, Texas, 77030, US; Weeks, Daniel E., University of Pittsburgh, Pittsburgh, Pennsylvania, 15260, US; Weinstock, Joshua, University of Michigan, Biostatistics, Ann Arbor, Michigan, 48109, US; Weir, Bruce, University of Washington, Seattle, Washington, 98195, US; Weiss, Scott T, Brigham & Women's Hospital, Channing Division of Network Medicine, Department of Medicine, Boston, Massachusetts, 02115, US; Weng, Lu-Chen, Massachusetts General Hospital, Boston, Massachusetts, 02114, US; Wessel, Jennifer, Indiana University, Epidemiology, Indianapolis, Indiana, 46202, US; Willer, Cristen, University of Michigan, Internal Medicine, Ann Arbor, Michigan, 48109, US; Williams, Kayleen, University of Washington, Biostatistics, Seattle, Washington, 98195, US; Williams, L. Keoki, Henry Ford Health System, Detroit, Michigan, 48202, US; Wilson, Carla, Brigham & Women's Hospital, Boston, Massachusetts, 02115, US; Wilson, James, Beth Israel Deaconess Medical Center, Cardiology, Cambridge, Massachusetts, 02139, US; Winterkorn, Lara, New York Genome Center, New York City, New York, 10013, US; Wong, Quenna, University of Washington, Seattle, Washington, 98195, US; Wu, Joseph, Stanford University, Stanford Cardiovascular Institute, Stanford, California, 94305, US; Xu, Huichun, University of Maryland, Baltimore, Maryland, 21201, US; Yanek, Lisa, Johns Hopkins University, Baltimore, Maryland, 21218, US; Yang, Ivana, University of Colorado at Denver, Denver, Colorado, 80204, US; Yu, Ketian, University of Michigan, Ann Arbor, Michigan, 48109, US; Zekavat, Seyedeh Maryam, Broad Institute, Cambridge, Massachusetts, 02142, US; Zhang, Yingze, University of Pittsburgh, Medicine, Pittsburgh, Pennsylvania, 15260, US; Zhao, Snow Xueyan, National Jewish Health, Denver, Colorado, 80206, US; Zhao, Wei, University of Michigan, Department of Epidemiology, Ann Arbor, Michigan, 48109, US; Zhu, Xiaofeng, Case Western Reserve University, Department of Population and Quantitative Health Sciences , Cleveland, Ohio, 44106, US; Zody, Michael, New York Genome Center, New York, New York, 10013, US; Zoellner, Sebastian, University of Michigan, Ann Arbor, Michigan, 48109, US

### Supplementary Note 5. REFERENCES

**Supplementary Figure 1. Scatter plots of PC1 *vs.* PC2 by population group**

Individuals with reported population memberships in each population group are denoted by filled circles in grey. Unfilled circles in colors represent inferred population memberships (N = 8,015), using Harmonized Ancestry and Race/Ethnicity (HARE) method (see methods for details). A) European, B) Amish, C) Samoan, D) Taiwanese, E) Han Chinese, F) Asian, G) Puerto Rican, H) Cuban, I) Central American, J) South American, K) Mexican, L) Costa Rican, M) Dominican, N) Barbadian, O) African/African American/Black.

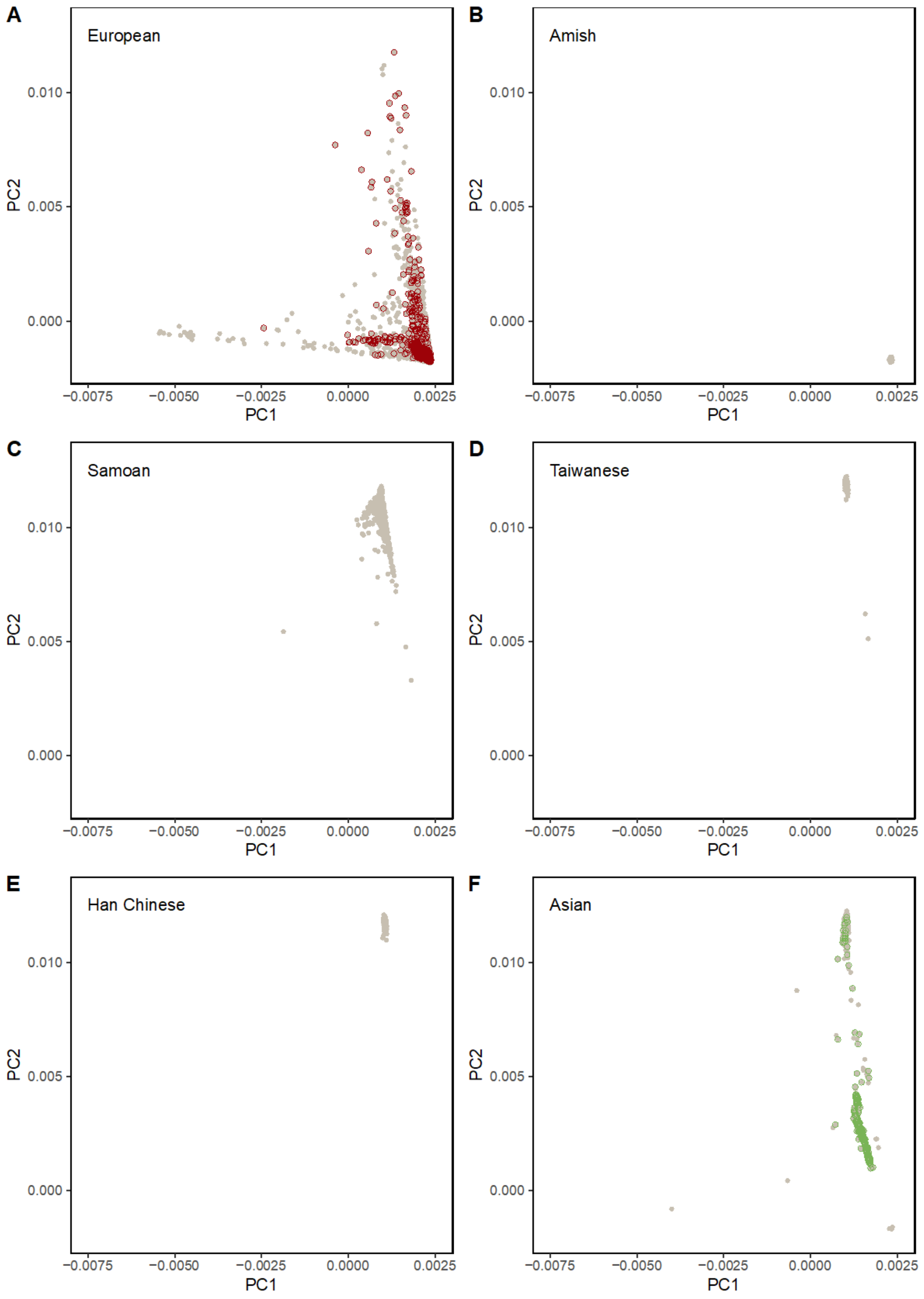

**Supplementary Figure 1. Scatter plots of PC1 *vs.* PC2 by population group**

Individuals with reported population memberships in each population group are denoted by filled circles in grey. Unfilled circles in colors represent inferred population memberships (N = 8,015), using Harmonized Ancestry and Race/Ethnicity (HARE) method (see methods for details). A) European, B) Amish, C) Samoan, D) Taiwanese, E) Han Chinese, F) Asian, G) Puerto Rican, H) Cuban, I) Central American, J) South American, K) Mexican, L) Costa Rican, M) Dominican, N) Barbadian, O) African/African American/Black.

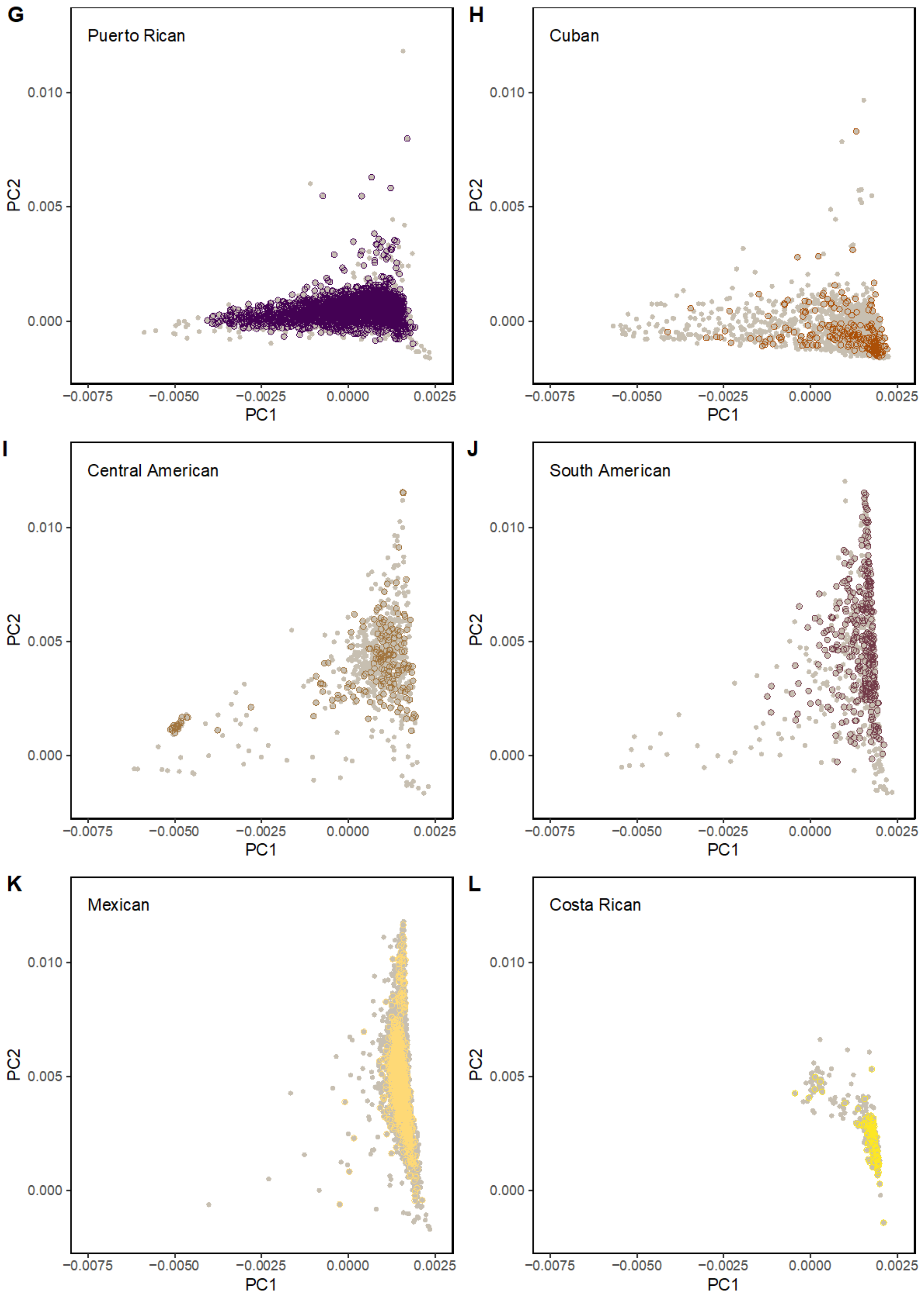

**Supplementary Figure 1. Scatter plots of PC1 *vs.* PC2 by population group**

Individuals with reported population memberships in each population group are denoted by filled circles in grey. Unfilled circles in colors represent inferred population memberships (N = 8,015), using Harmonized Ancestry and Race/Ethnicity (HARE) method (see methods for details). A) European, B) Amish, C) Samoan, D) Taiwanese, E) Han Chinese, F) Asian, G) Puerto Rican, H) Cuban, I) Central American, J) South American, K) Mexican, L) Costa Rican, M) Dominican, N) Barbadian, O) African/African American/Black.

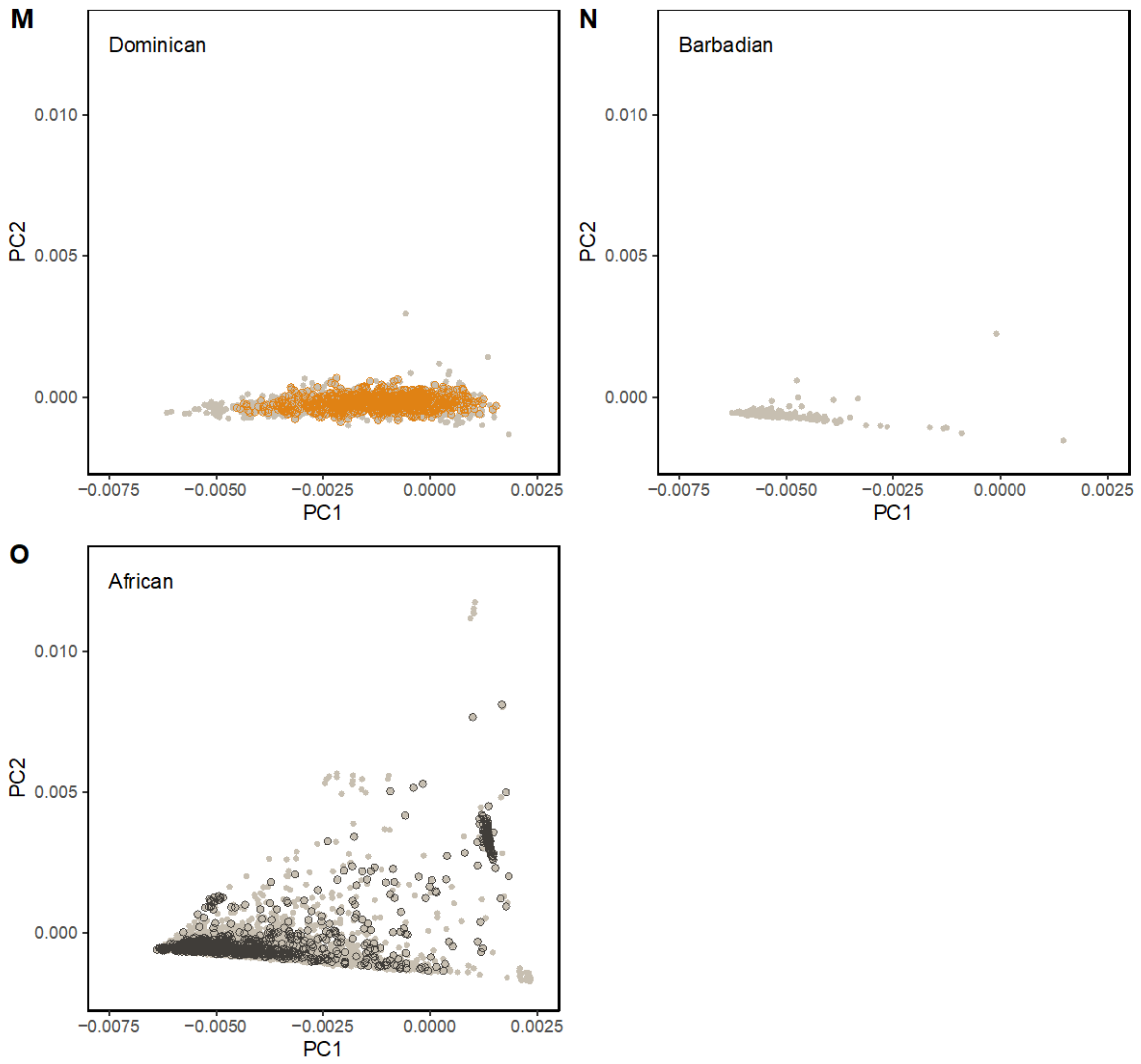

**Supplementary Figure 2. QQ plot of primary BMI GWAS**

Quantile-quantile plot of multi-population, single variant analysis (N = 88,873 individuals, N = 90,142,062 variants).

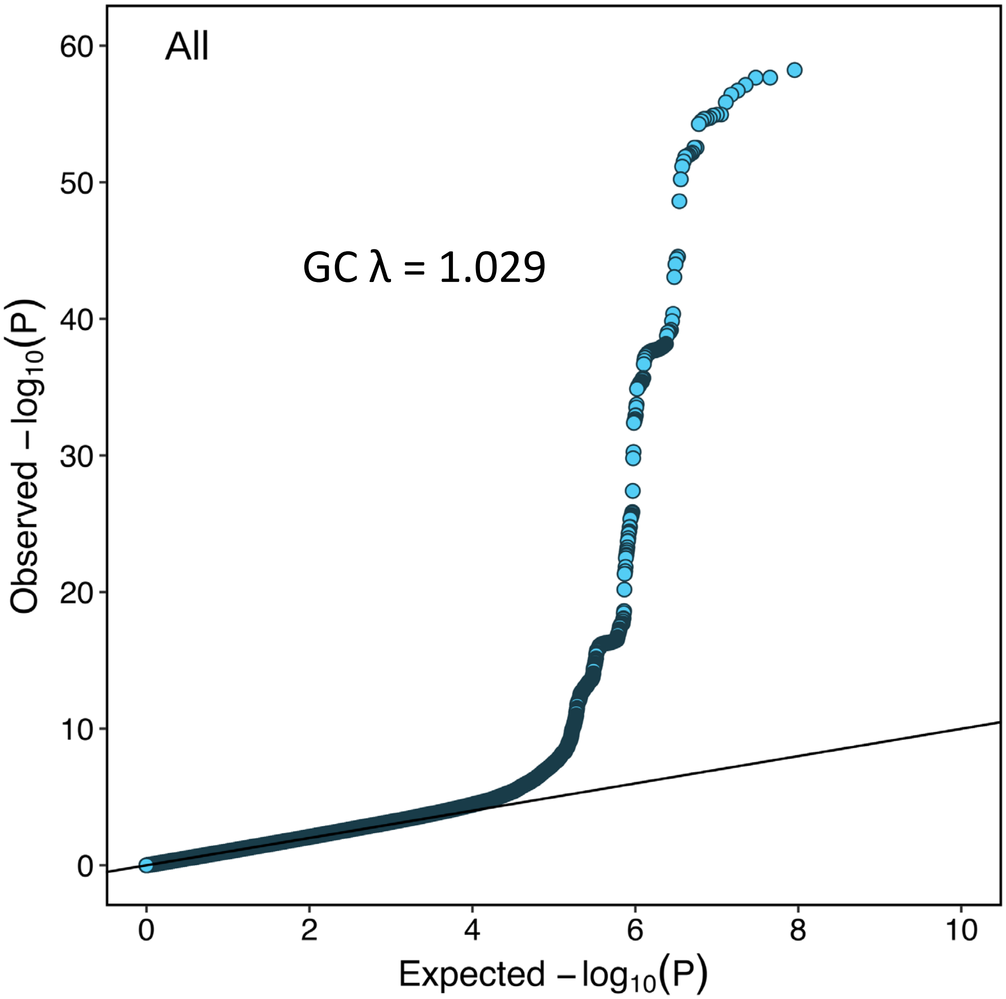

**Supplementary Figure 3. Regional association plots**

Regional association plots for each significant locus in the multi-population analysis, including all variants ± 500 kb from index variant. The plots appear in order of chromosomal location. TOPMed study populations were used to calculate linkage disequilibrium (LD). The red line indicates genome-wide significance threshold $P = 5 \times 10^{-9}$.

A) *SEC16B*, rs543874; B) *TMEM18*, rs939584; C) *ADCY3*, rs10182181; D) *ETV5*, rs869400; E) *GNPDA2*, rs12507026; F) *POC5*, rs2307111; G) *TFAP2B*, rs2206277; H) *HNF4G*, rs830463; I) *BDNF*, rs3838785; J) *BCDIN3D*, rs7138803; K) *OLFM4*, rs9568868; L) *FTO*, rs1421085; M) *MC4R*, rs6567160; N) *ZC3H4*, rs28590228; O) *MTMR3*, rs111490516; P) *DMD*, rs1379871.

### A) ***SEC16B*, rs543874**

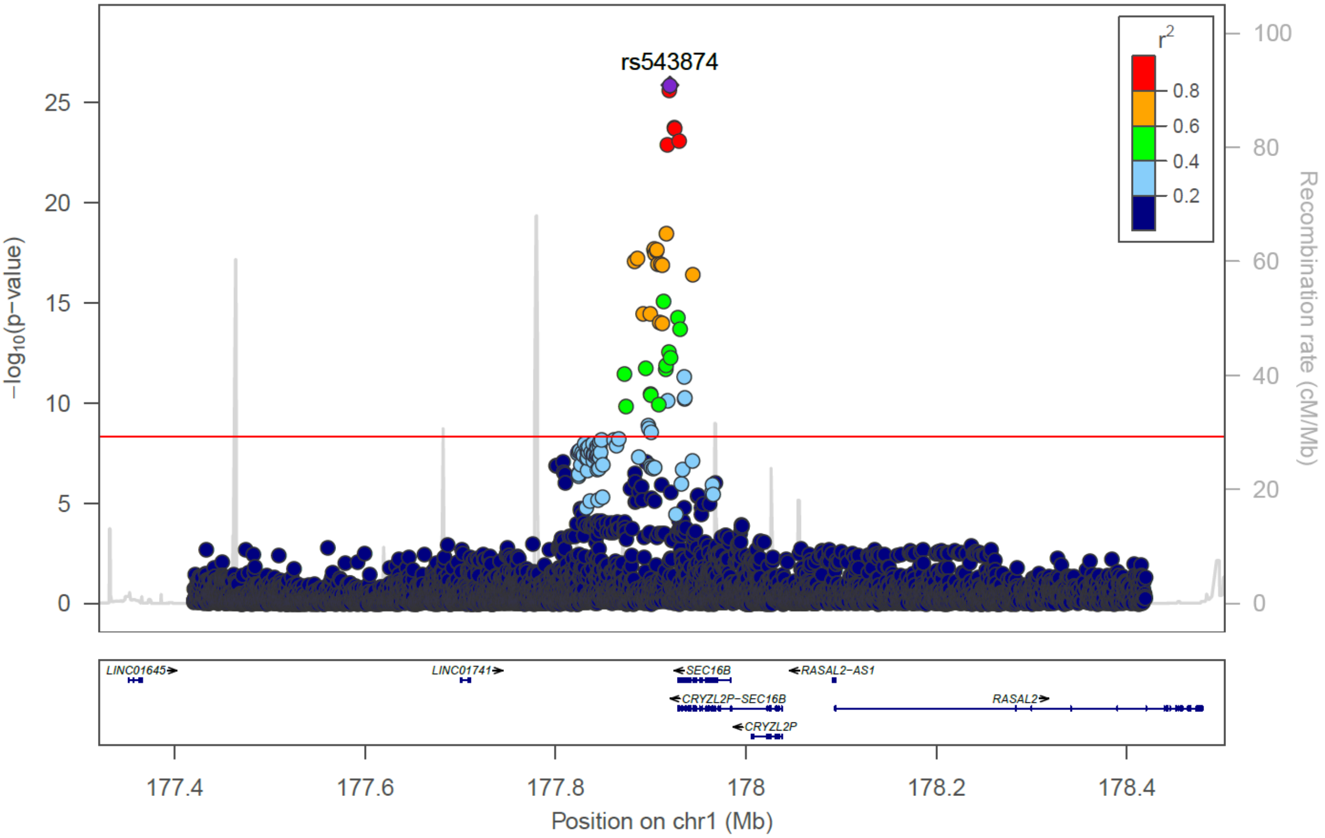

B) *TMEM18*, rs939584

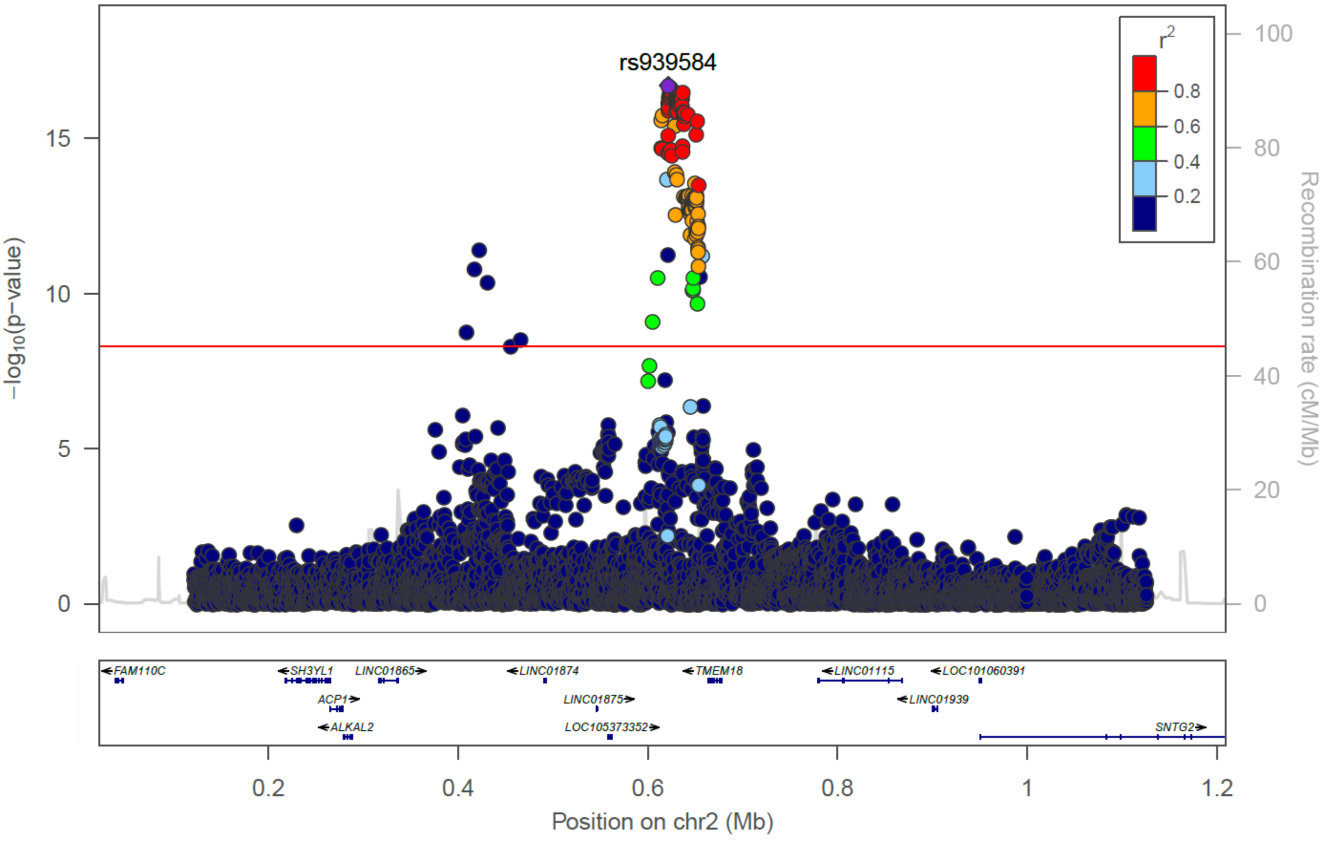


C) *ADCY3*, rs10182181

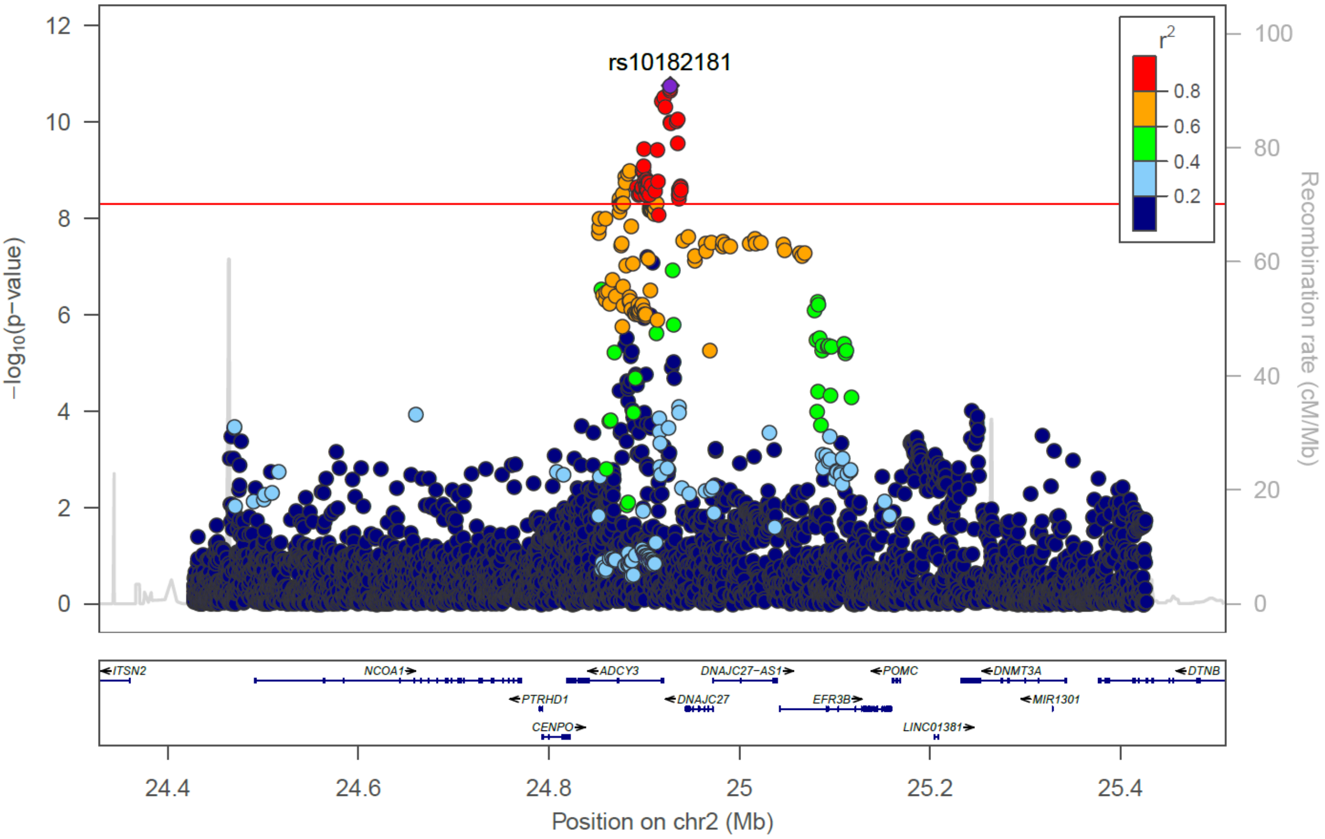

**D) *ETV5*, rs869400**

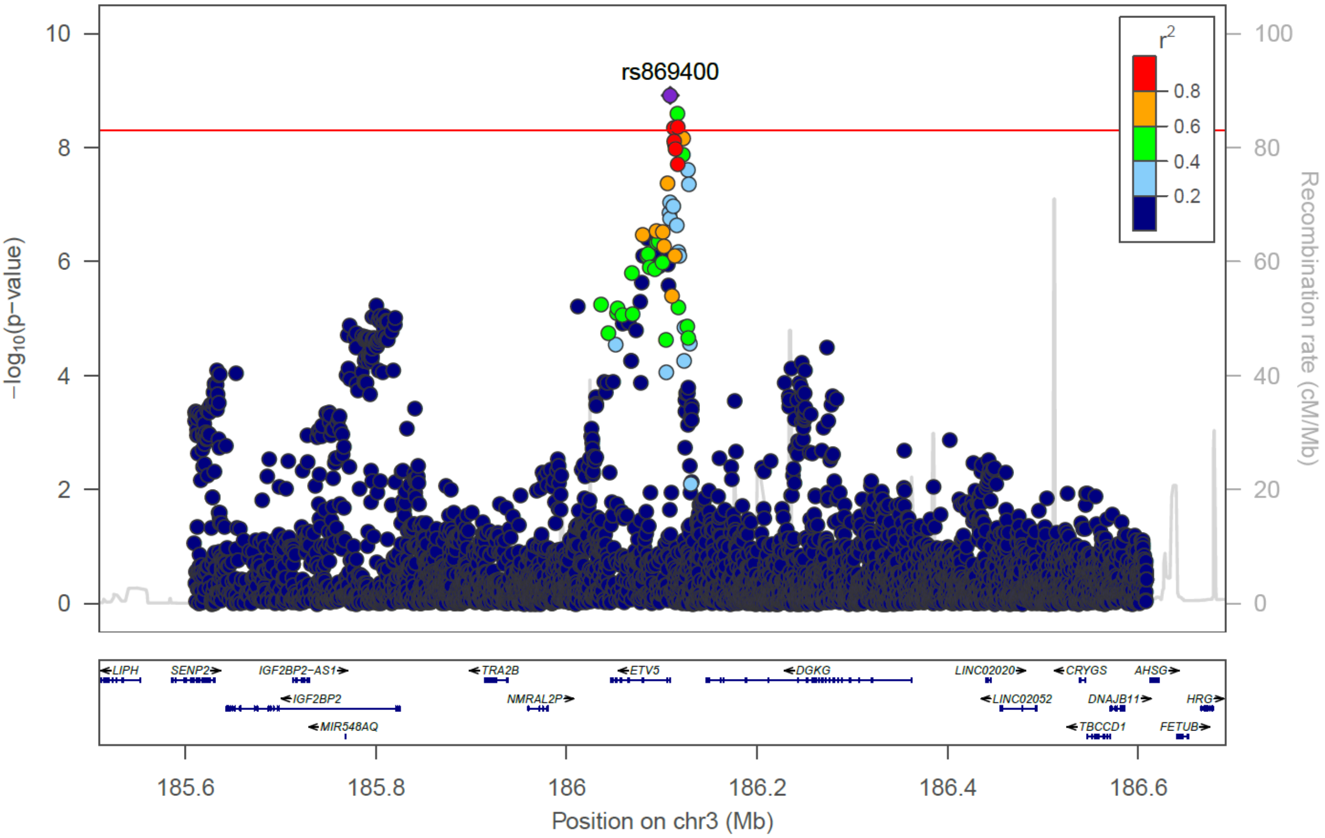


**E) *GNPDA2*, rs12507026**

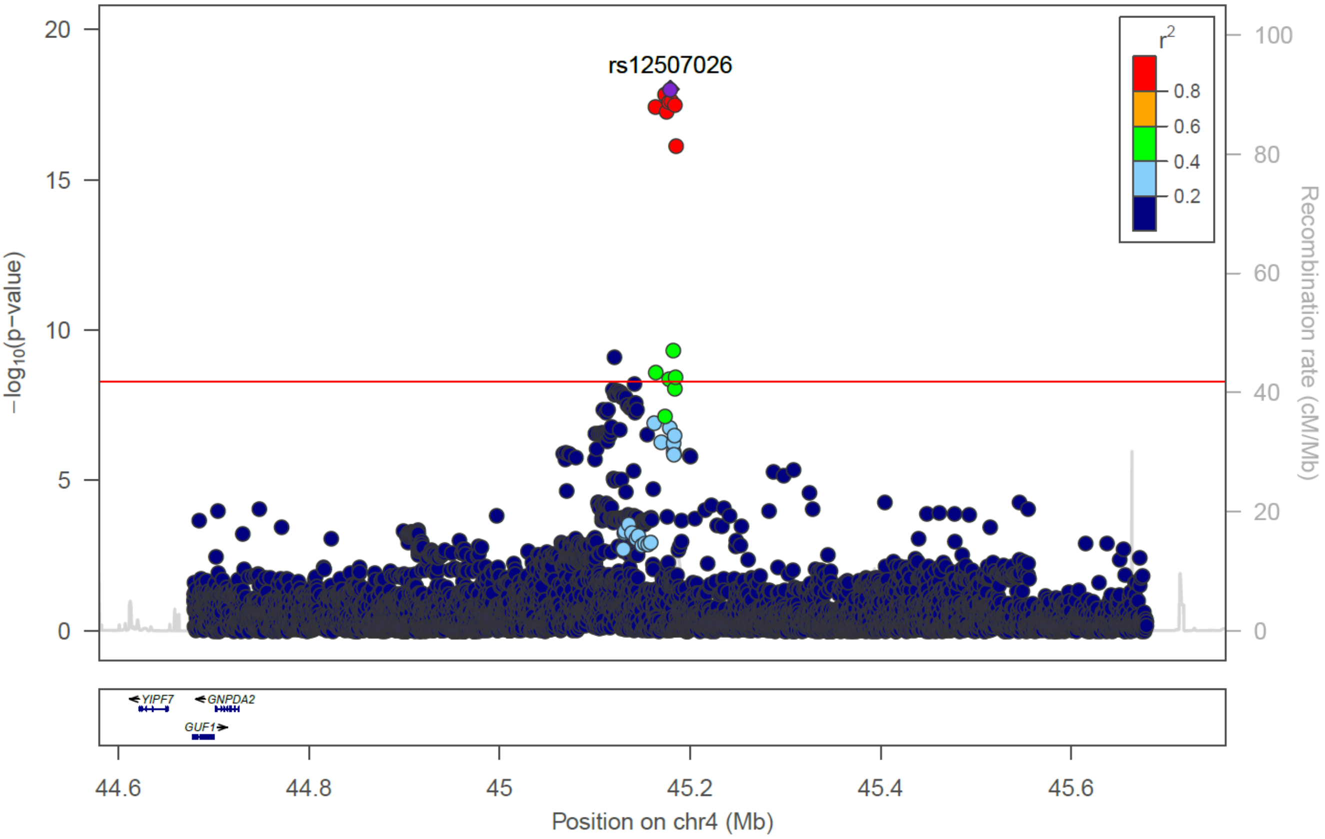

F) *POC5*, rs2307111

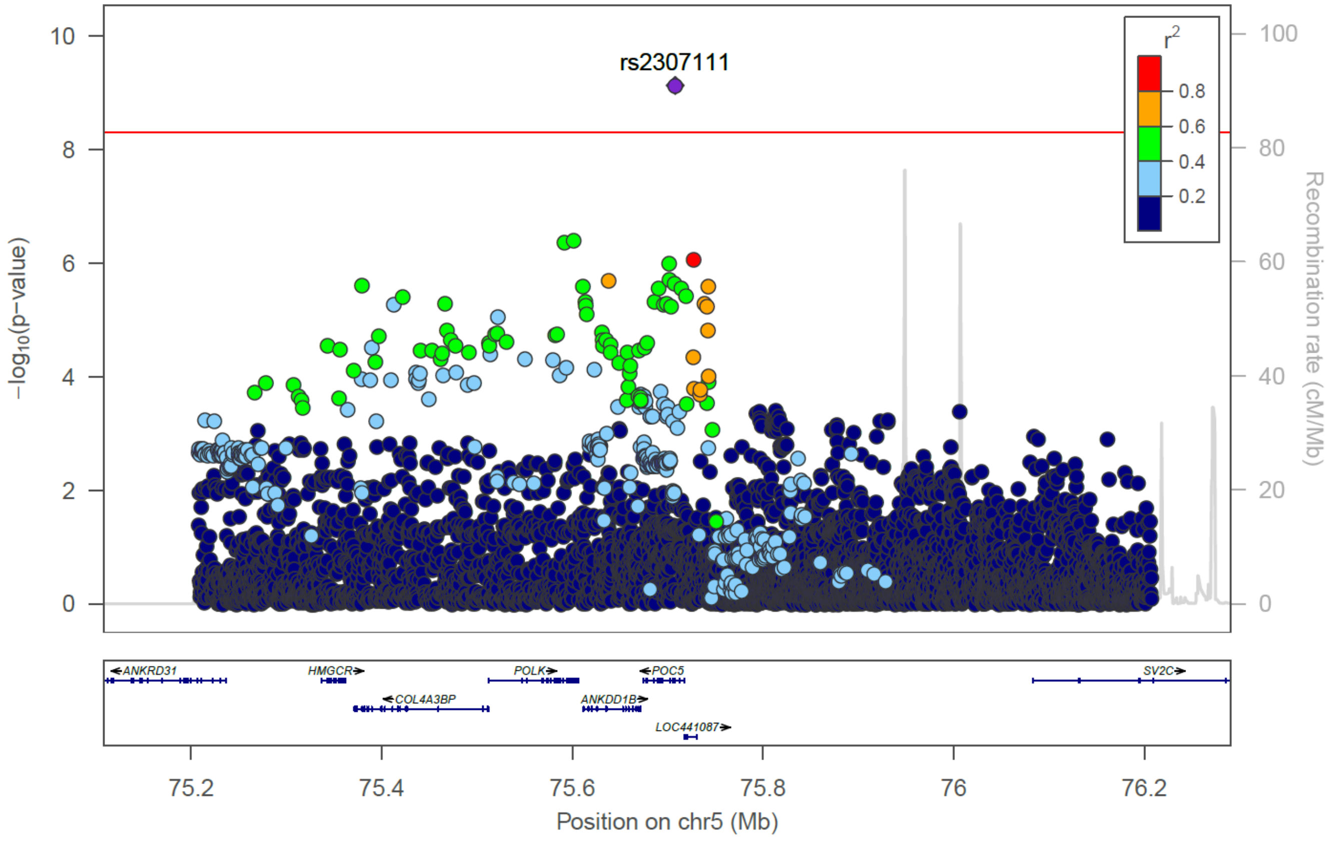


G) *TFAP2B*, rs2206277

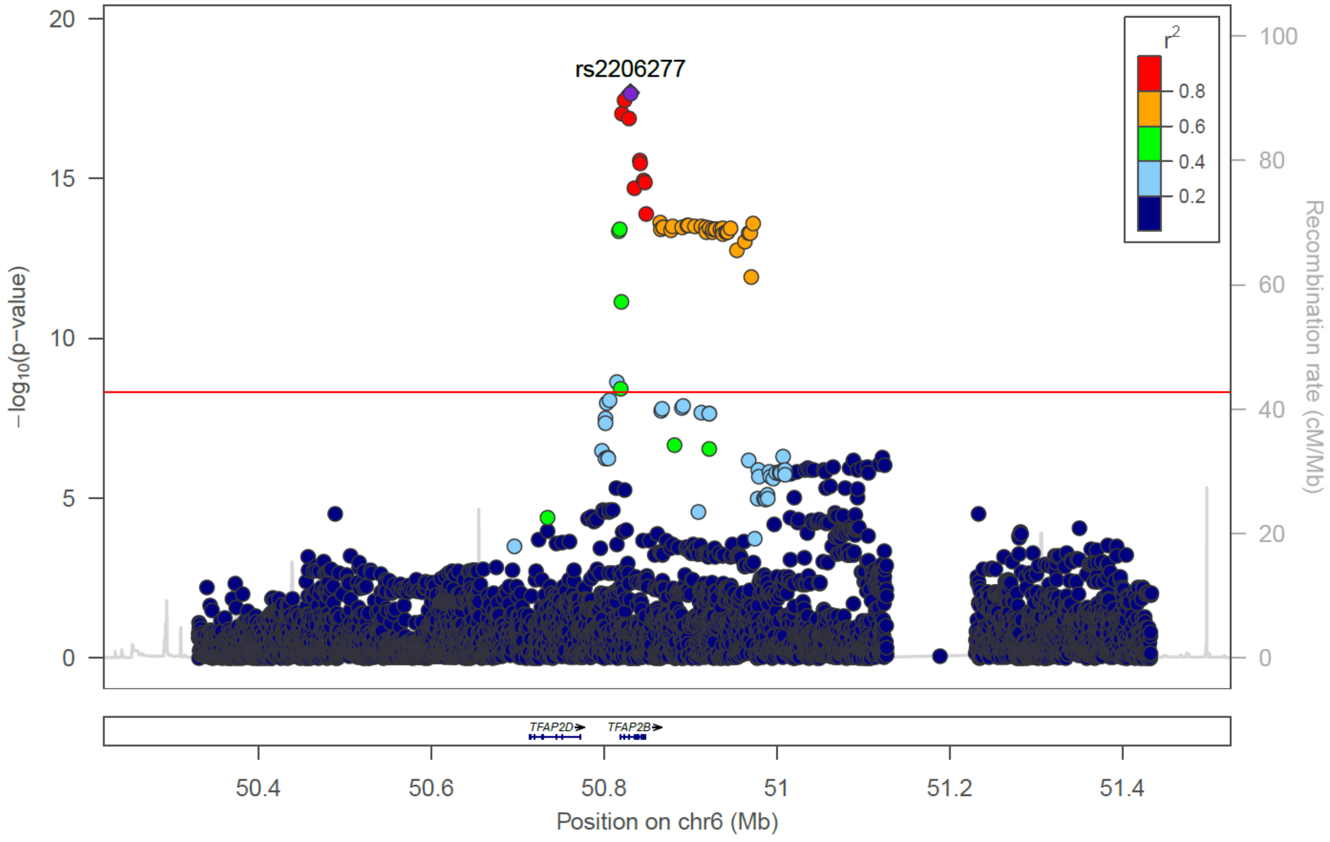

**H) *HNF4G*, rs830463**

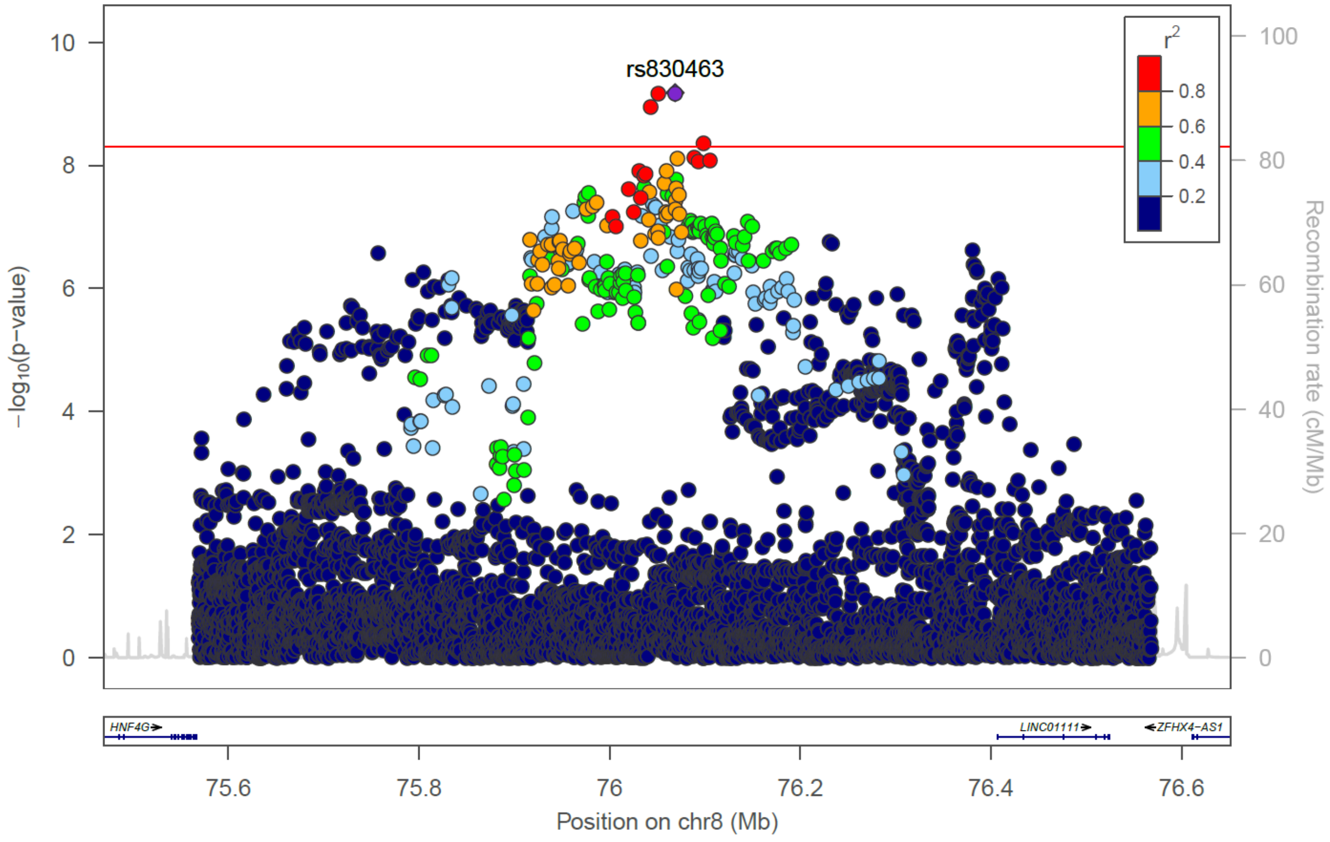


**I) *BDNF*, rs3838785**

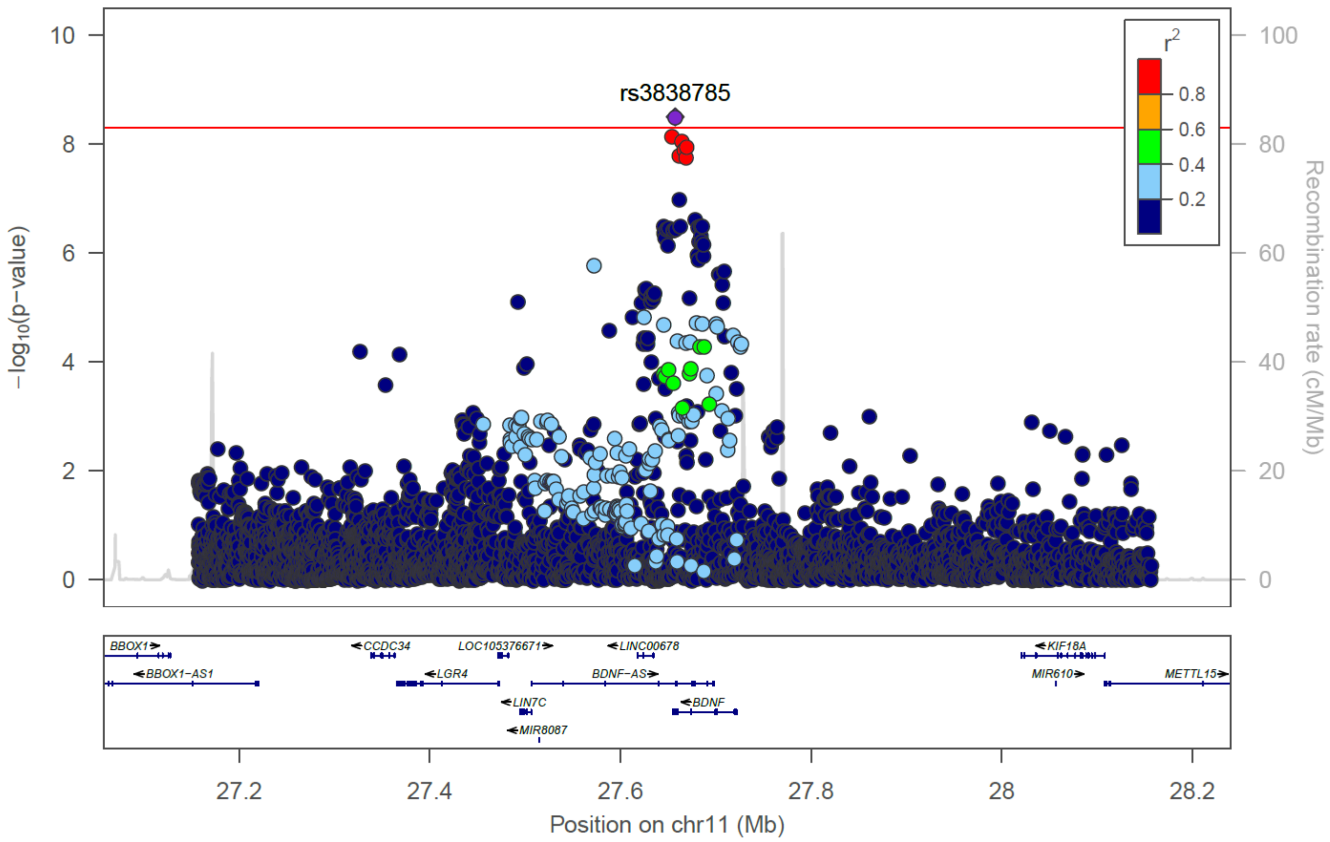

**J) *BCDIN3D*, rs7138803**

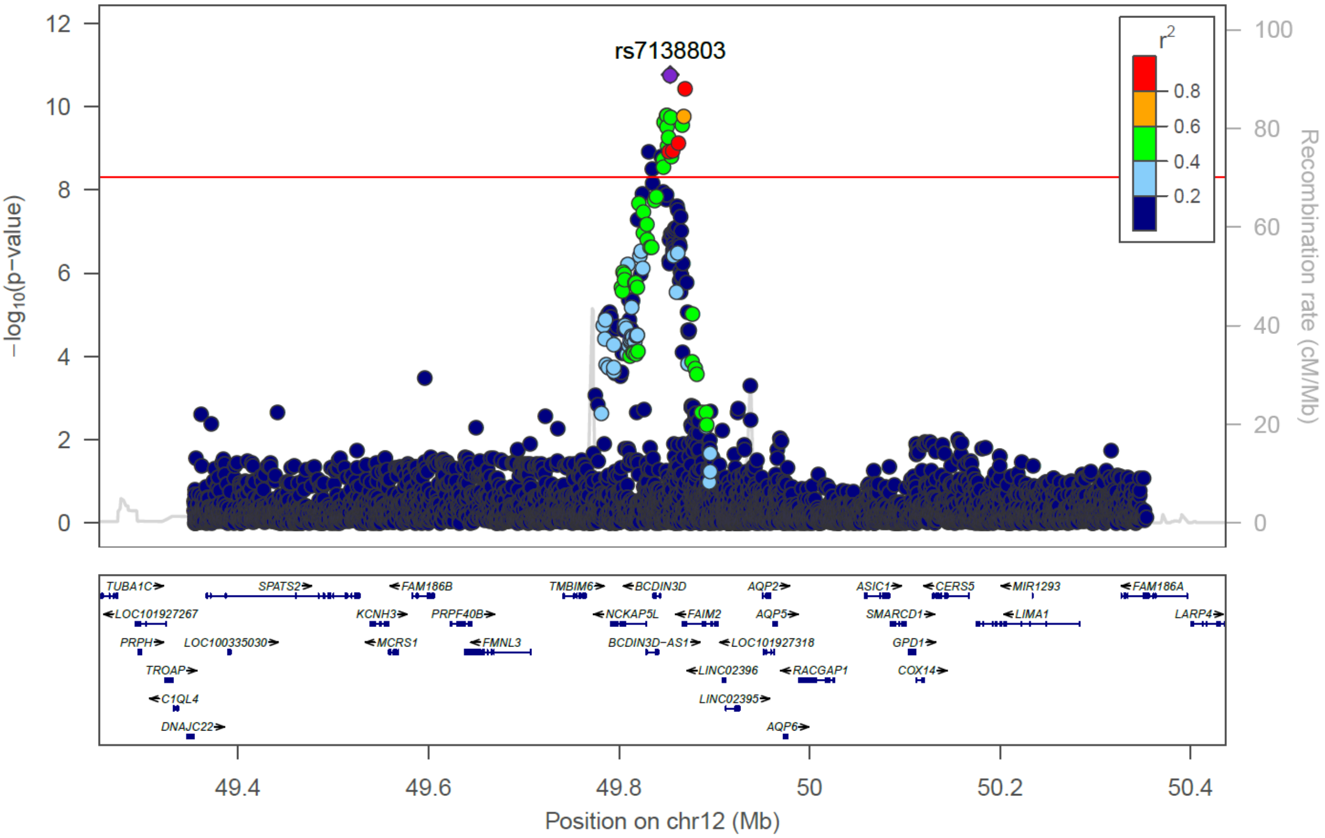


**K) *OLFM4*, rs9568868**

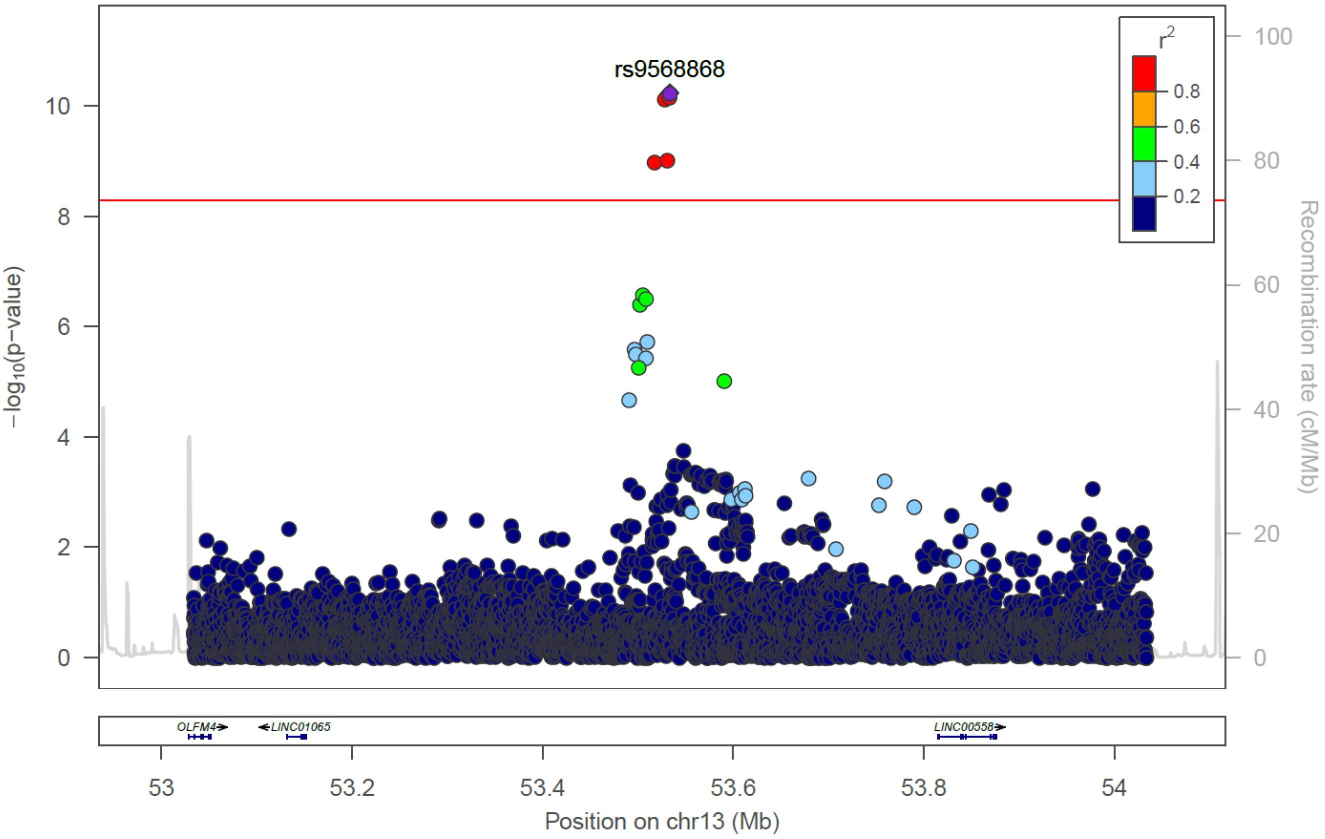

### L) *FTO*, rs1421085

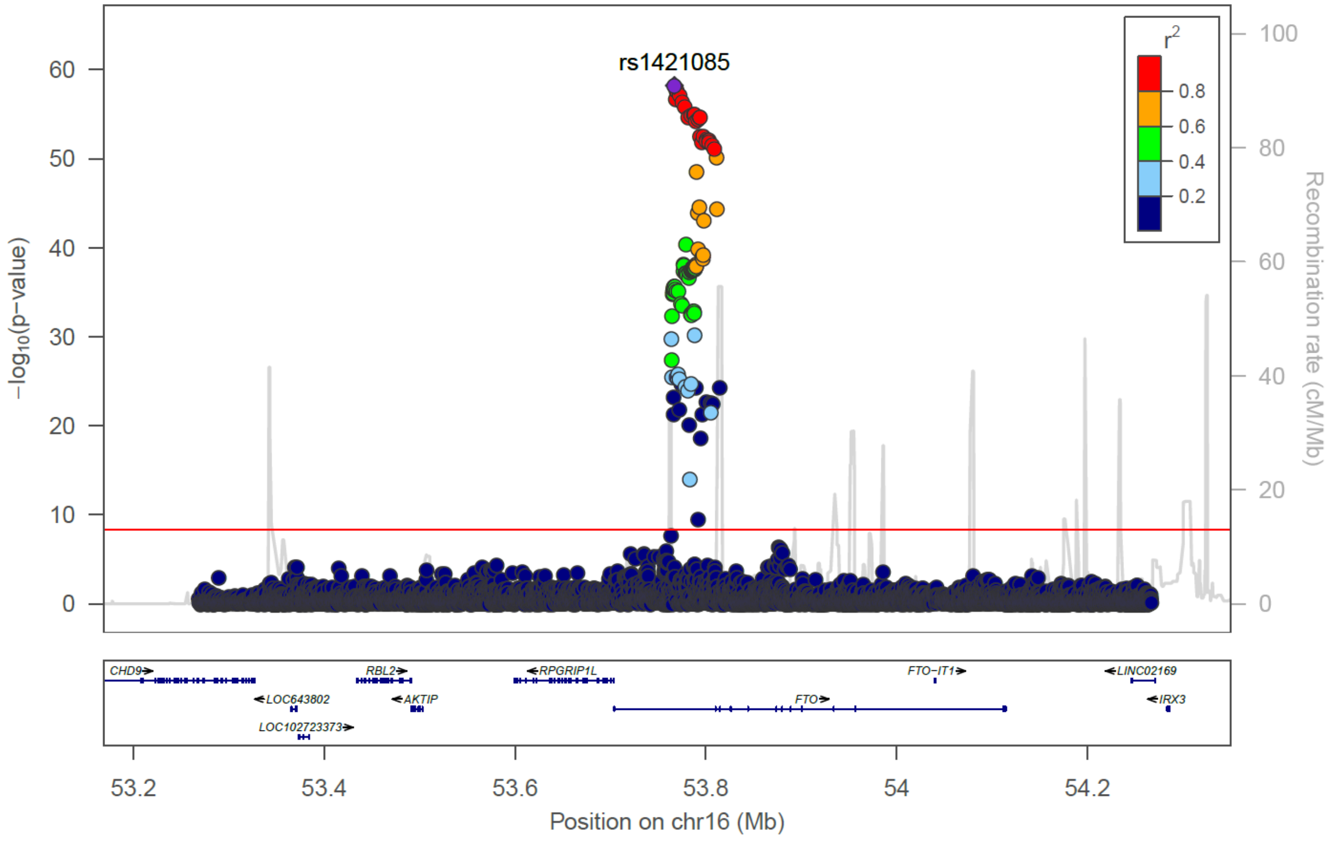


## M) *MC4R*, rs6567160

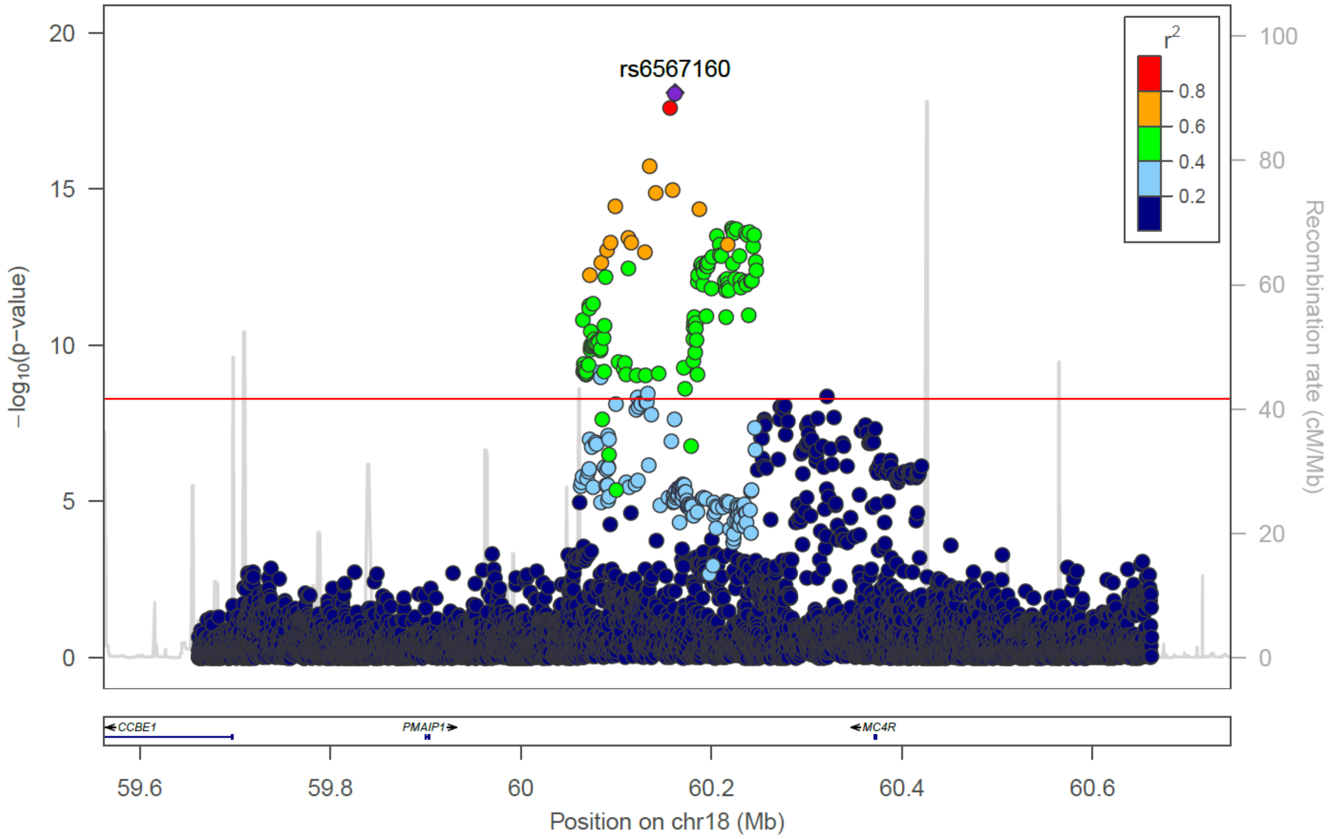

N) *ZC3H4*, rs28590228

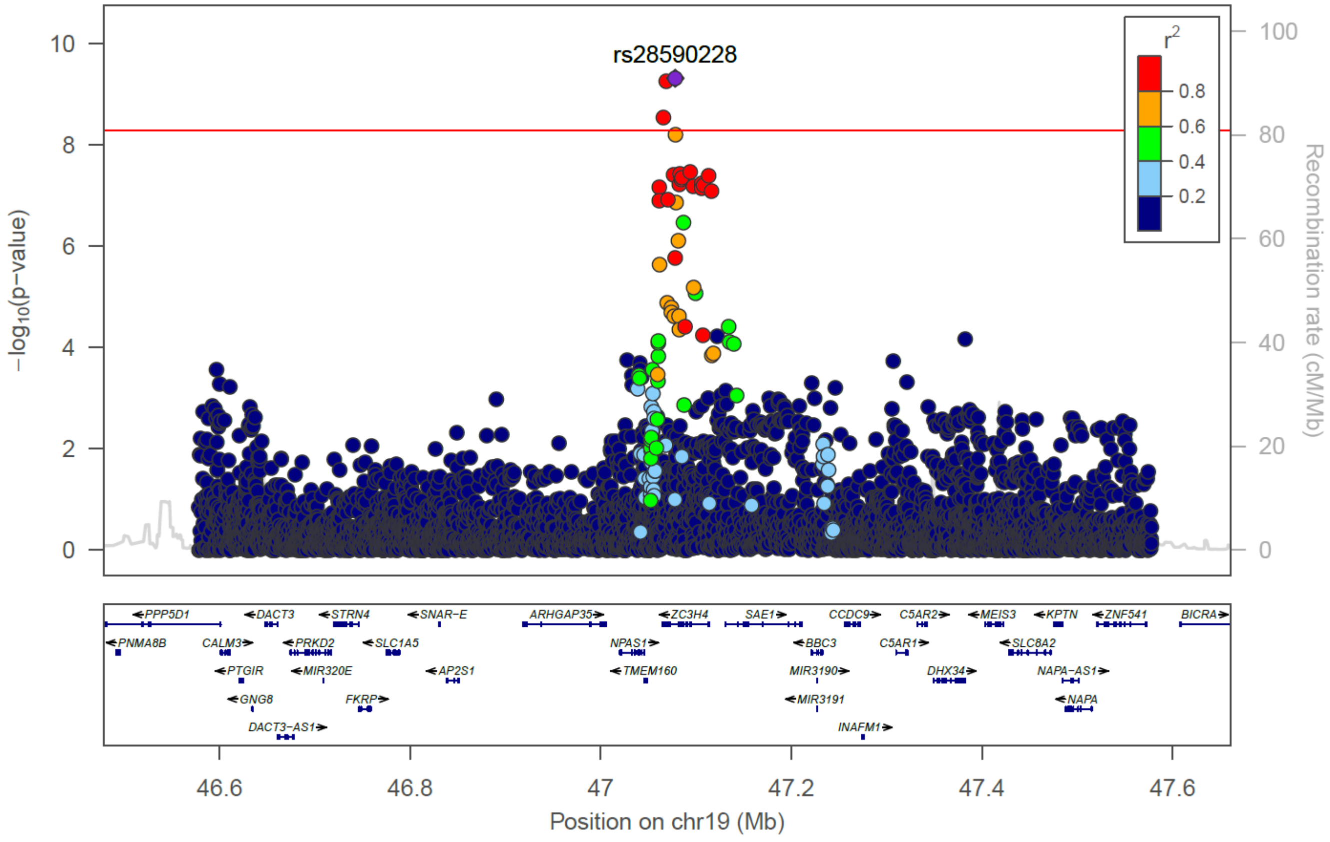


O) *MTMR3*, rs111490516

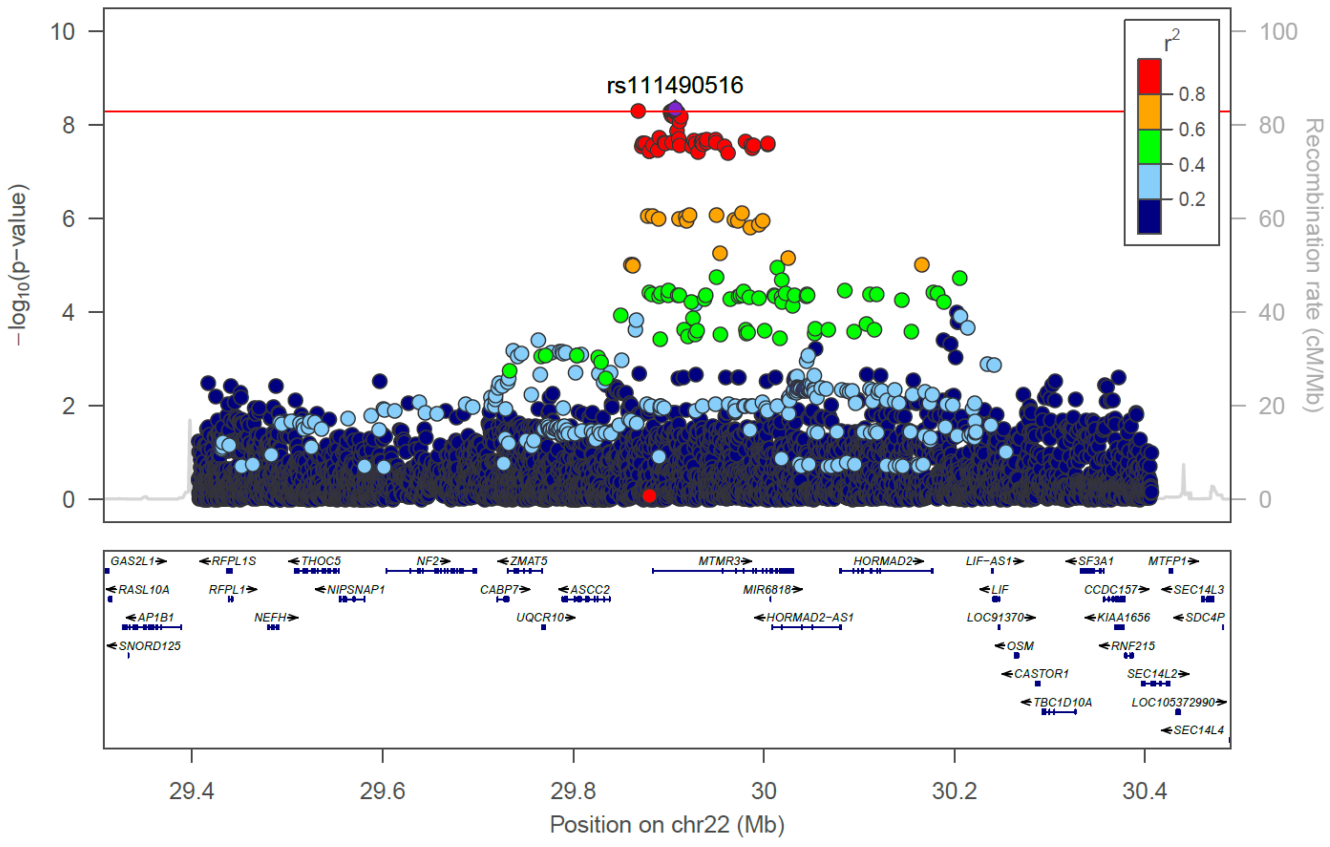

### P) *DMD*, rs1379871

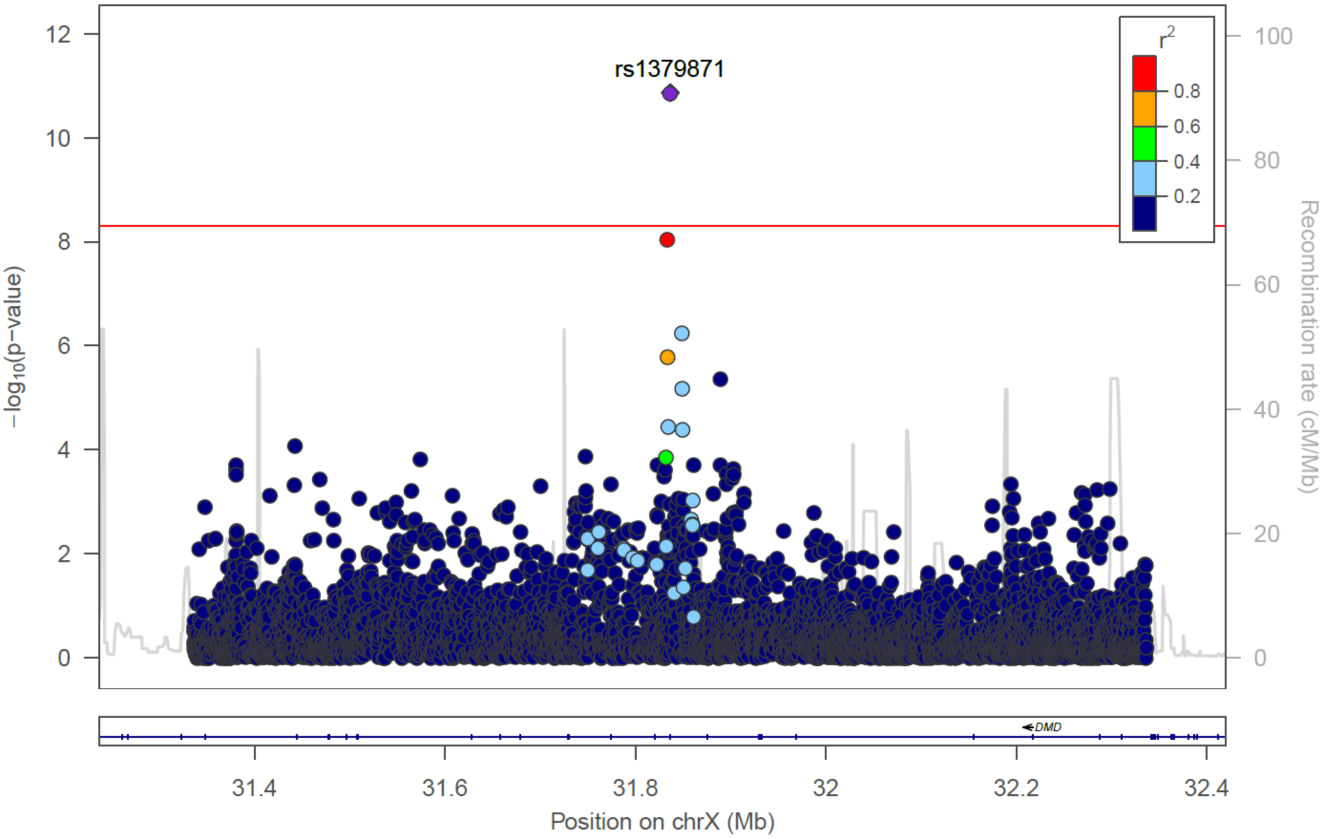

**Supplementary Figure 4. Manhattan plot of African population group BMI GWAS**

Manhattan plot of African population group, single variant analysis (N = 22,488 individuals). The novel locus *(MTMR3)* is highlighted in red. Previously reported BMI loci are in dark beige. The horizontal dashed line indicates genome-wide significant threshold $P = 5 \times 10^{-9}$.

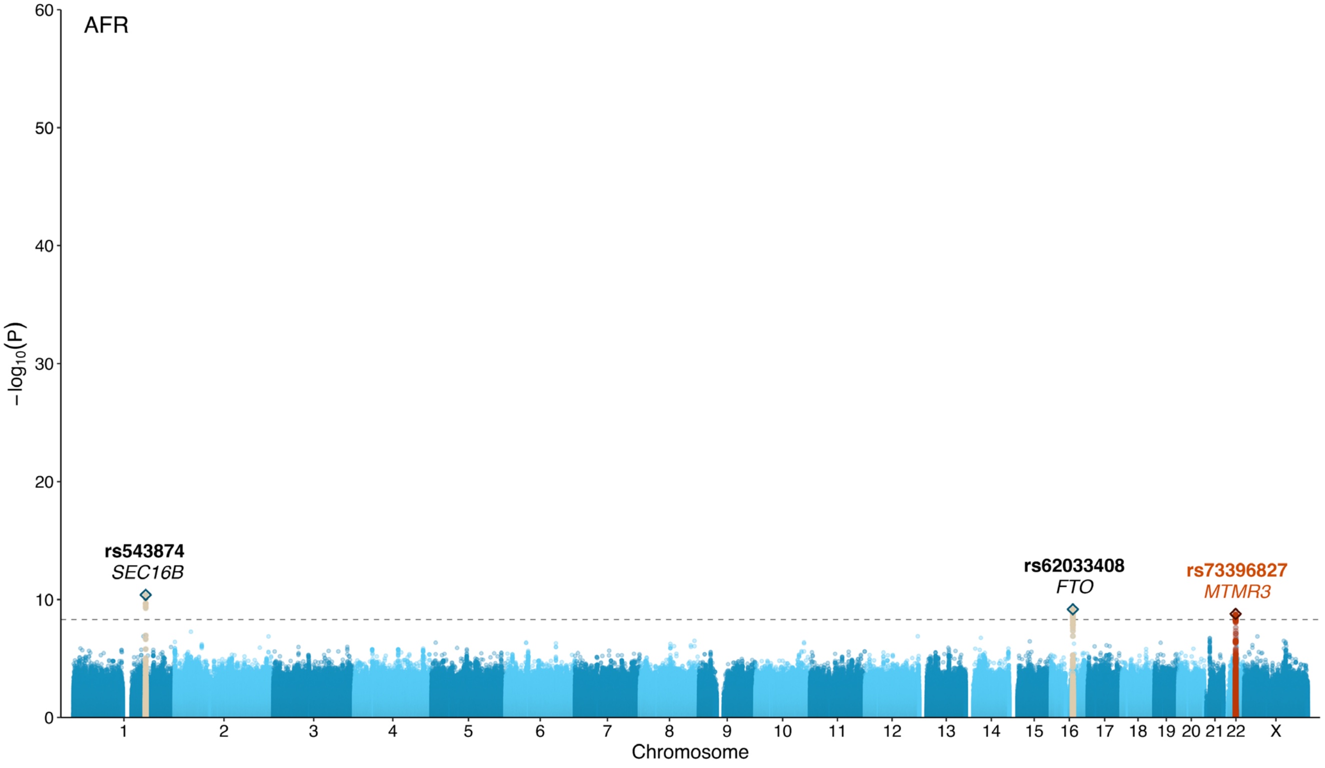

**Supplementary Figure 5. QQ plot of African population group BMI GWAS**

Quantile-quantile plot of African population group, single variant analysis (N = 22,488 individuals).

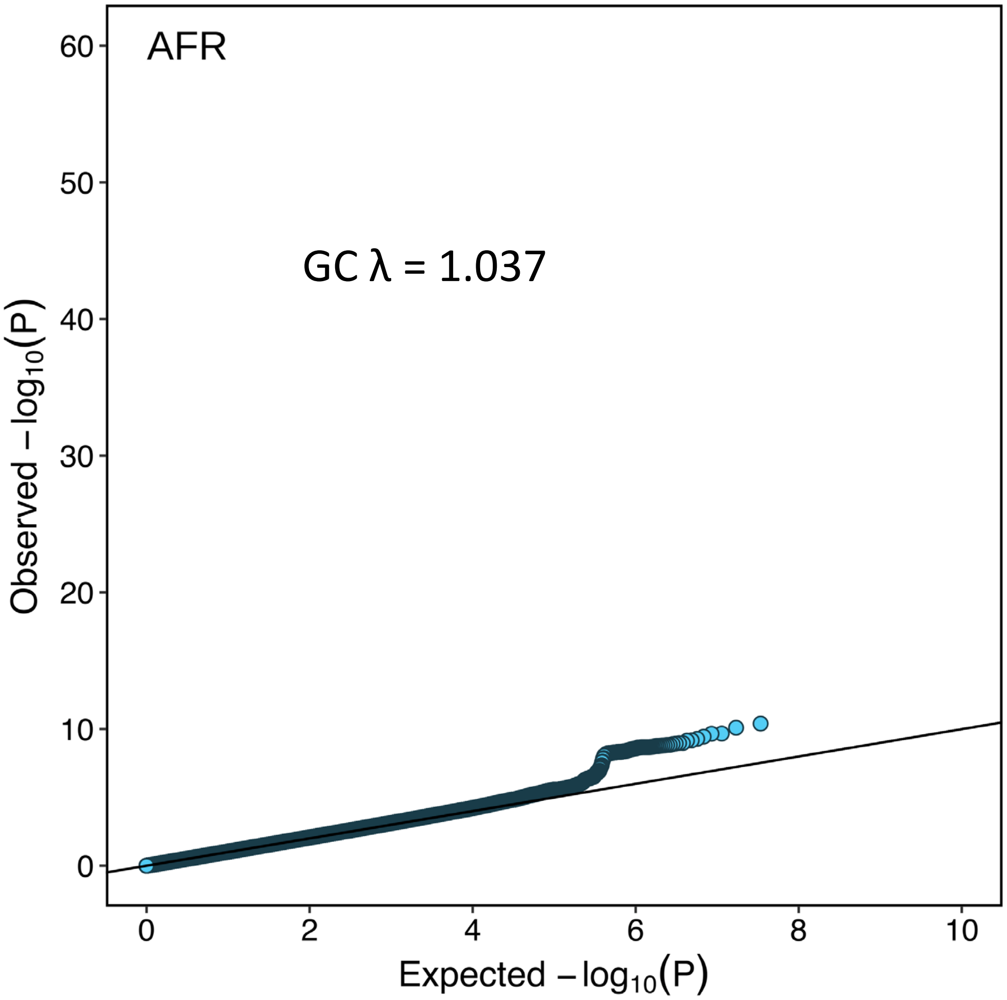

**Supplementary Figure 6. Manhattan plot of European population group BMI GWAS**

Manhattan plot of European population group, single variant analysis (N = 43,434 individuals). Previously reported BMI loci are in dark beige. The horizontal dashed line indicates genome-wide significant threshold $P = 5 \times 10^{-9}$.

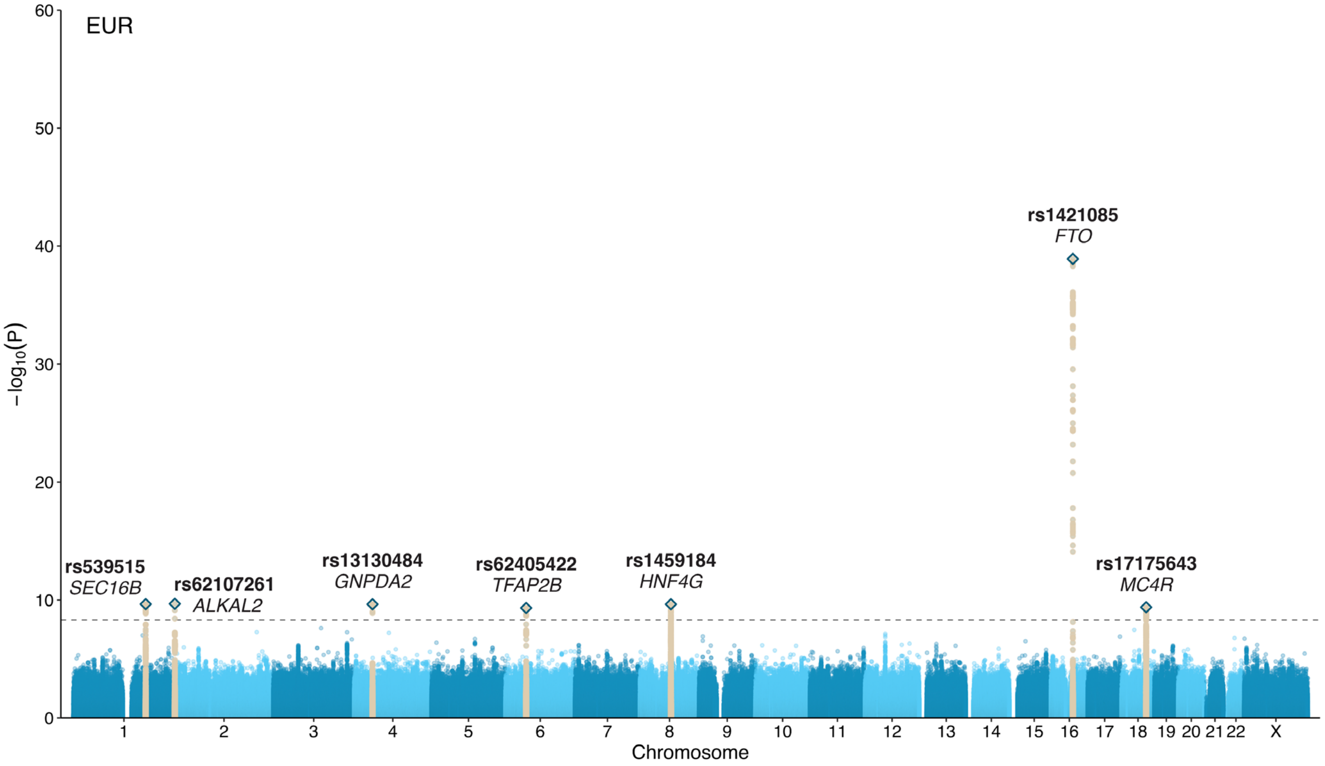

**Supplementary Figure 7. QQ plot of European population group BMI GWAS**

Quantile-quantile plot of European population group, single variant analysis (N = 43,434 individuals).

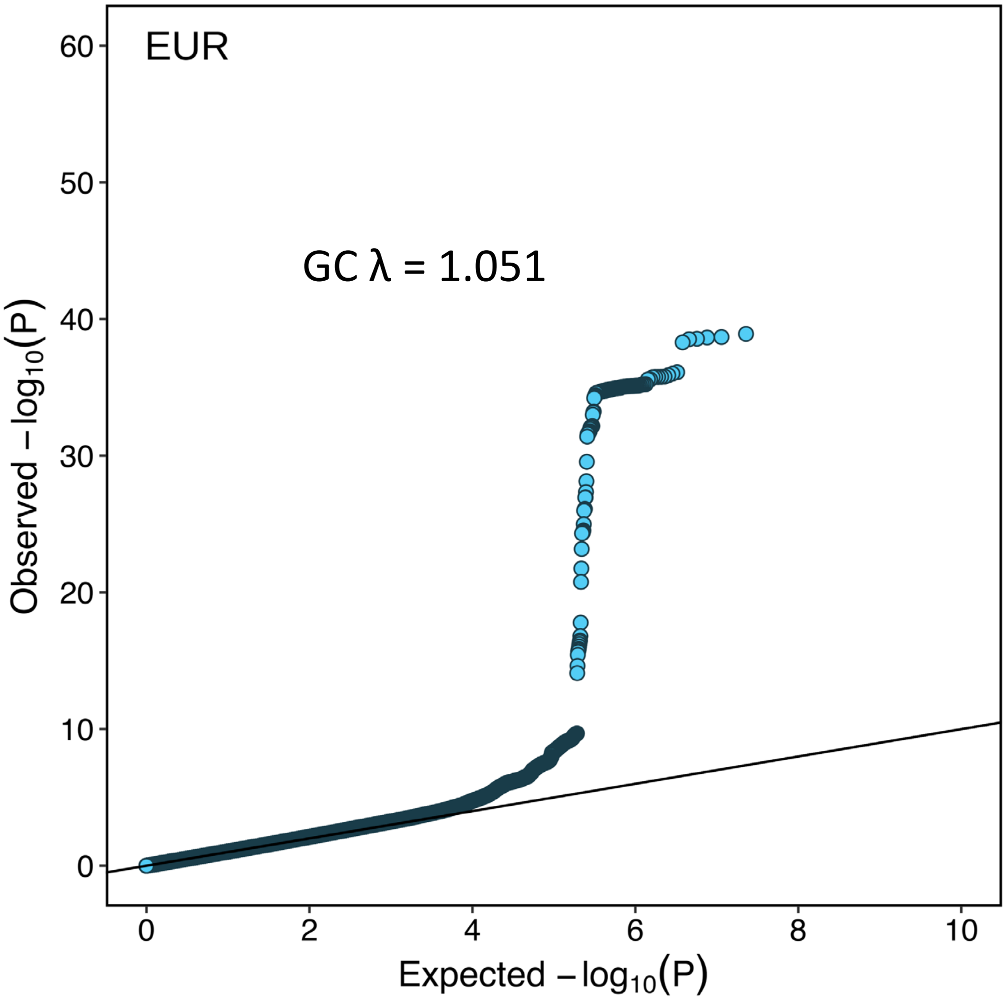

**Supplementary Figure 8. Regional association plots of secondary signals**

Regional association plots for each significant secondary signal in the multi-population analysis following conditional analysis on top variant, including all variants ± 500 kb from index variant. TOPMed study populations were used to calculate LD. The red line indicates $P = 5.67 \times 10^{-7}$. A) *ALKAL2*, rs62107261; B) *MC4R*, rs78769612.

### A) *ALKAL2*, rs62107261

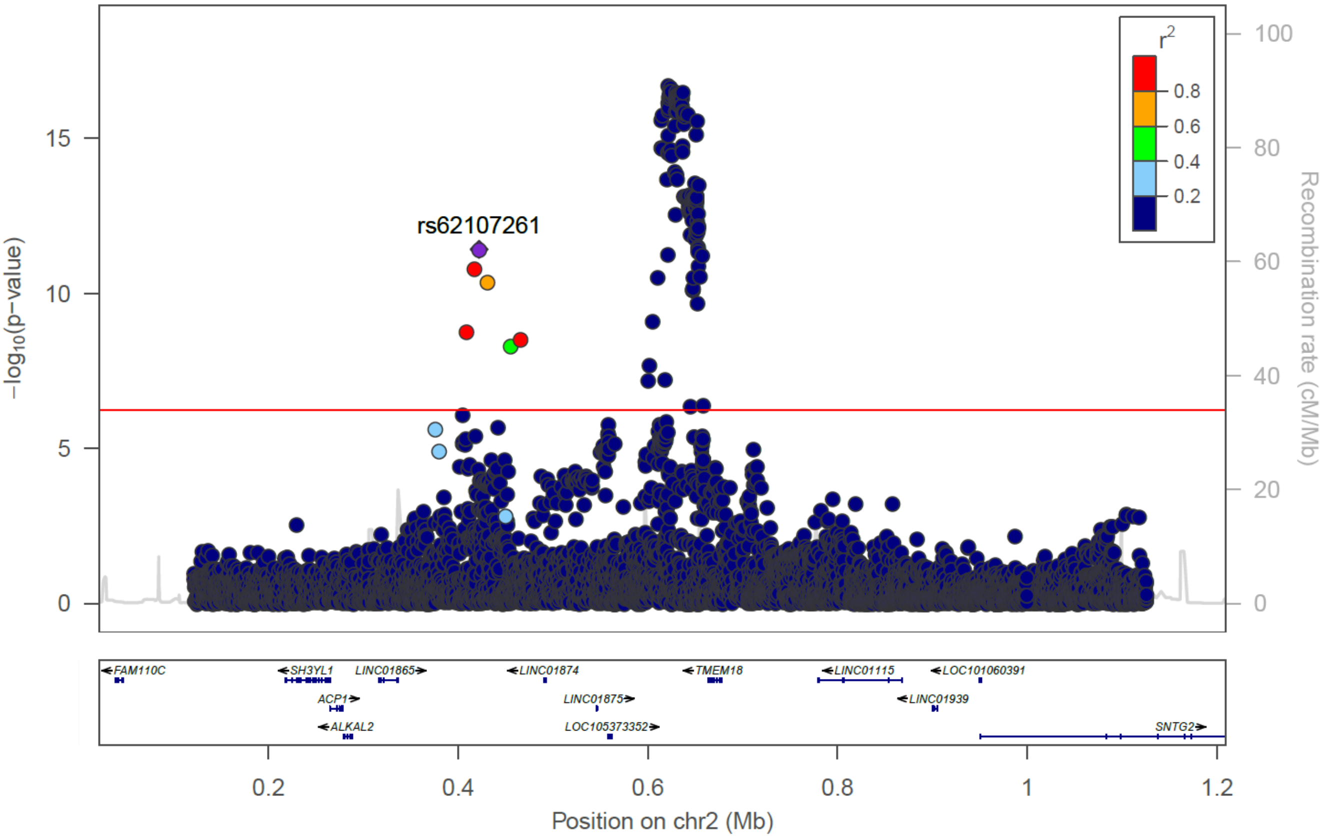


## B) *MC4R*, rs78769612

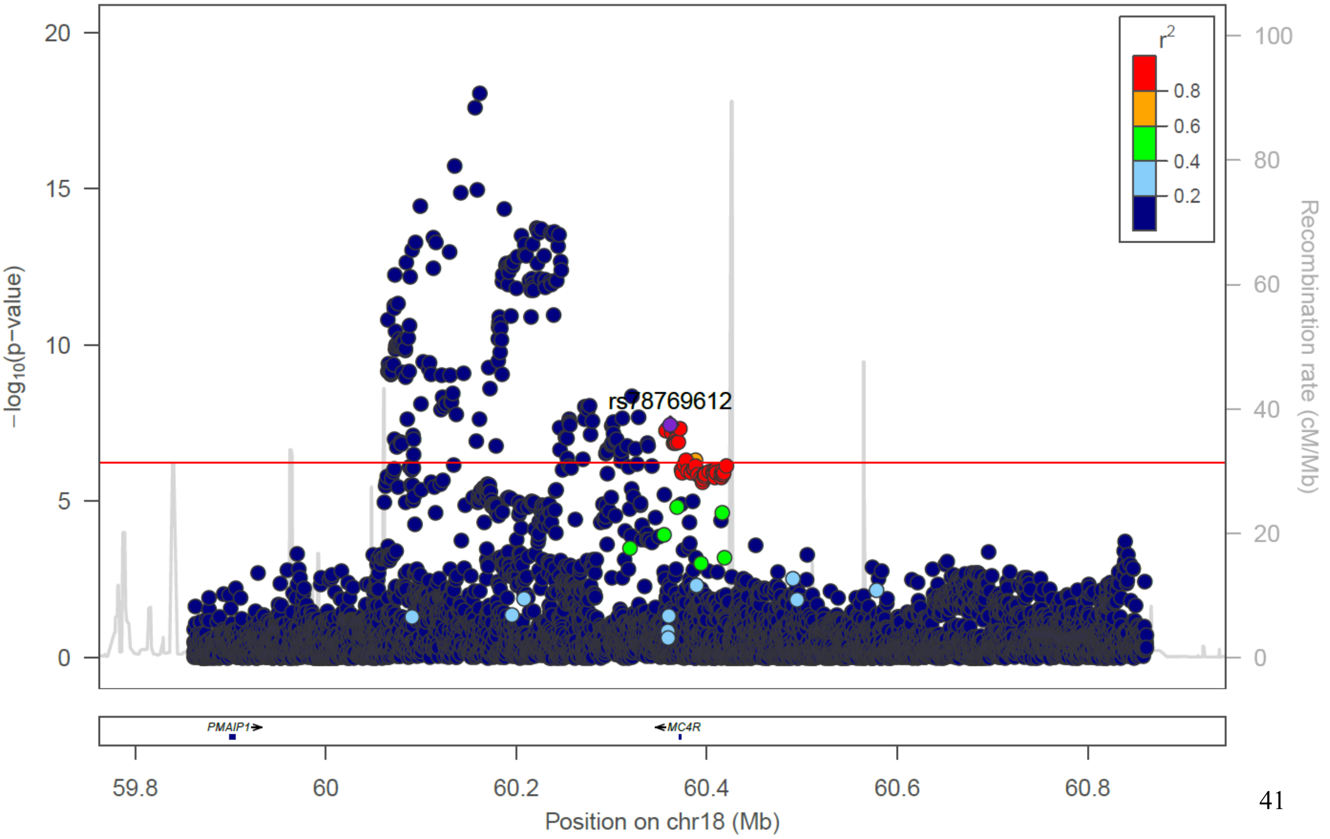

**Supplementary Figure 9. LD matrix heatmap for conditionally independent SNPs in known BMI-risk loci**

Pairwise LD matrix heatmap for lead index SNP in discovery analyses (bold and underlined) and published BMI GWAS SNPs within 500 kb (+/-) of index SNPs. A) rs2206277 index SNP in *TFAP2B* locus; B) rs3838785 in *BDNF* locus.

A.

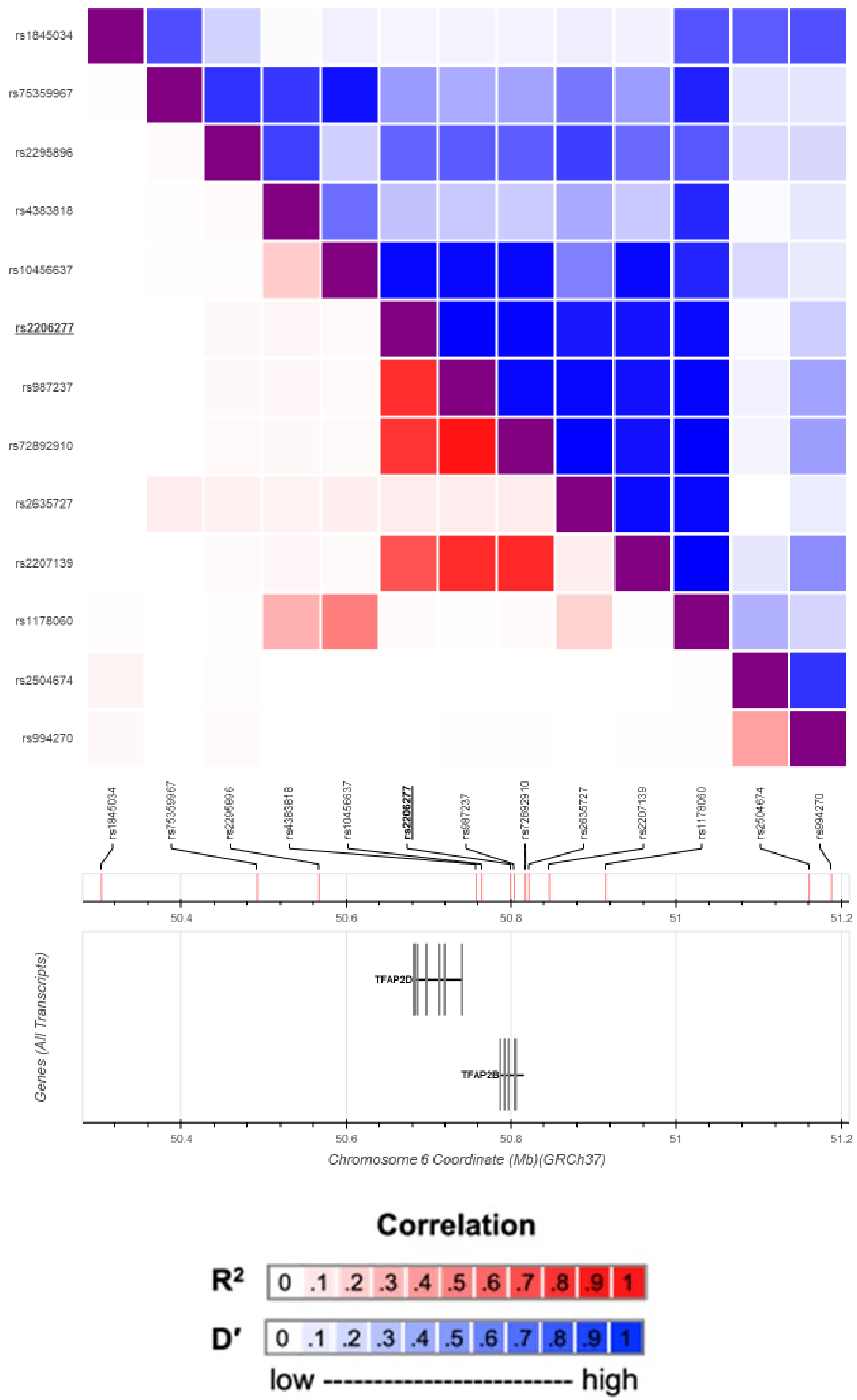

**Supplementary Figure 9. LD matrix heatmap for conditionally independent SNPs in known BMI-risk loci**

Pairwise LD matrix heatmap for lead index SNP in discovery analyses (bold and underlined) and published BMI GWAS SNPs within 500 kb (+/-) of index SNPs. A) rs2206277 index SNP in *TFAP2B* locus; B) rs3838785 in *BDNF* locus.

**B.**

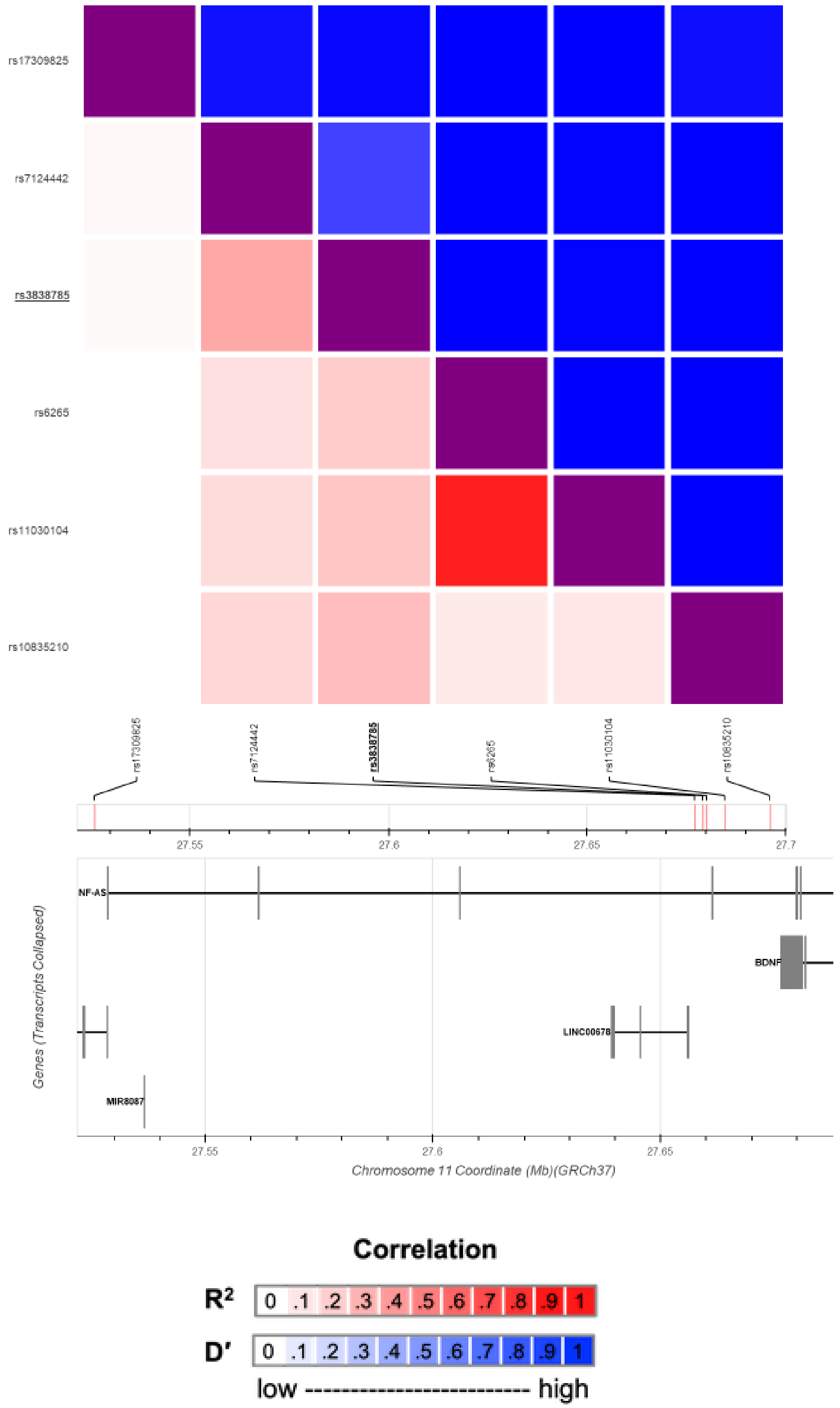

**Supplementary Figure 10**. **Fine-mapping regional plots**

Regional plots of posterior probability (PP) from fine-mapping analysis in PAINTOR, including all variants ± 100 kb from index variant for each locus with any variant exhibiting a moderate PP > 0.5. The plots appear in order of chromosomal location. TOPMed study populations were used to calculate LD. Shape and color indicate potential functional consequence of each variant as reported in Variant Effect Prediction (VEP) tool or GeneHancer (see methods for details).

A) rs543874, *SEC16B;* B) rs869400, *ETV5*; C) rs2307111, *POC5*; D) rs1421085, *FTO*; E) rs6567160, *MC4R*; F) rs55731973, *ZC3H4*; G) rs1379871, *DMD*.

**A) rs543874 (chr1:177920345), *SEC16B***

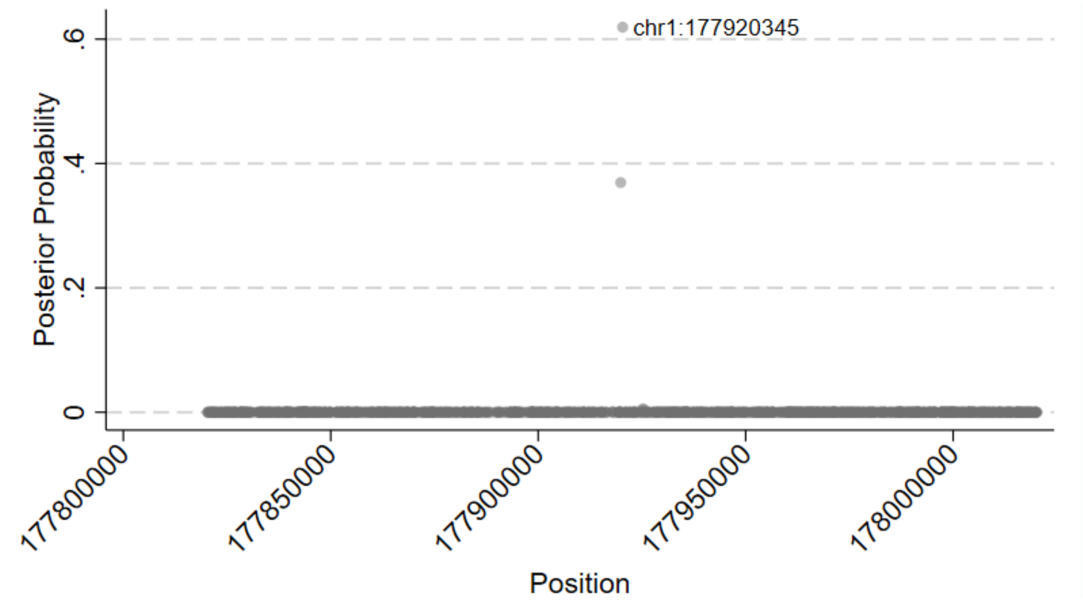


**B) rs869400 (chr3:186108951), *ETV5***

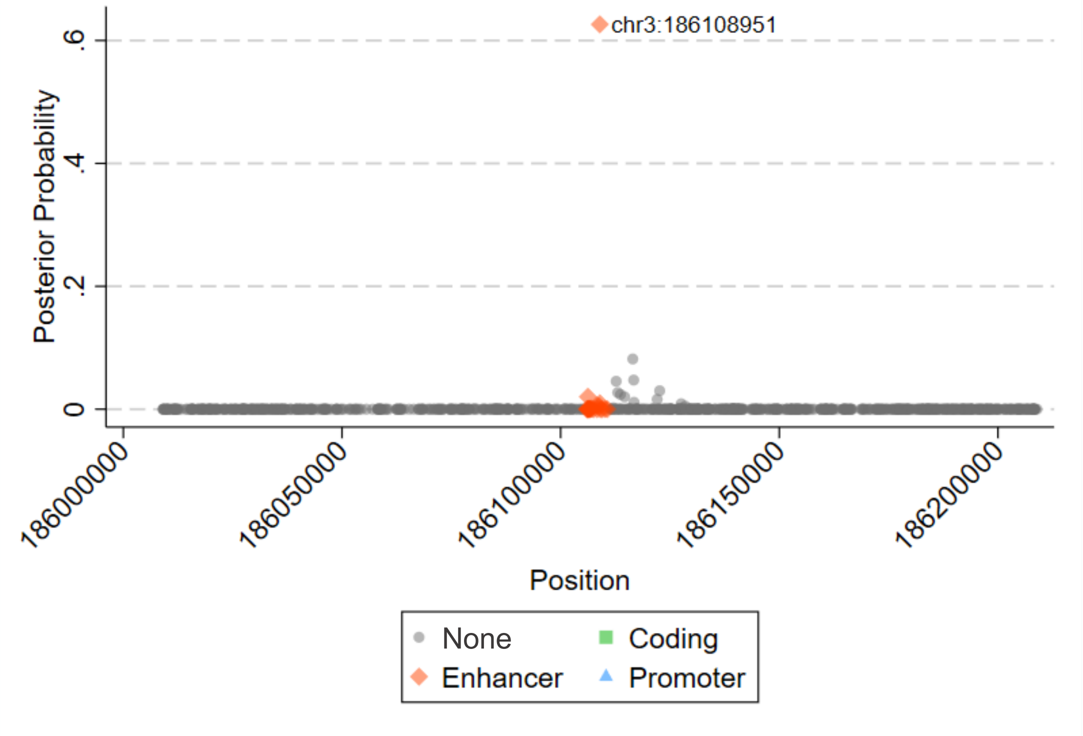

**C) rs2307111 (chr5: 75707853), *POC5***

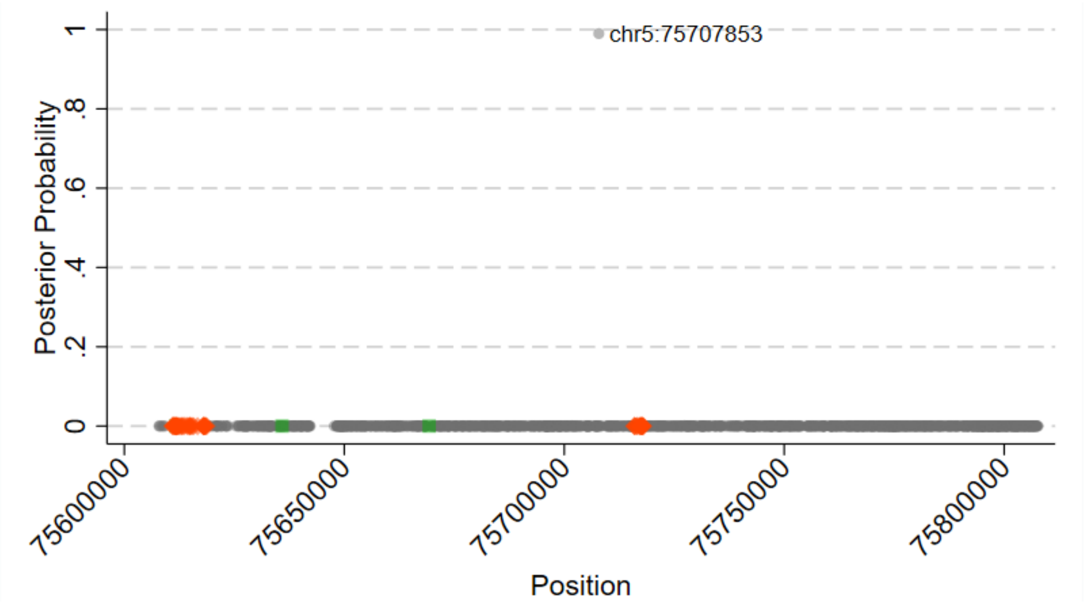


**D) rs1421085 (chr16:53767042), *FTO***

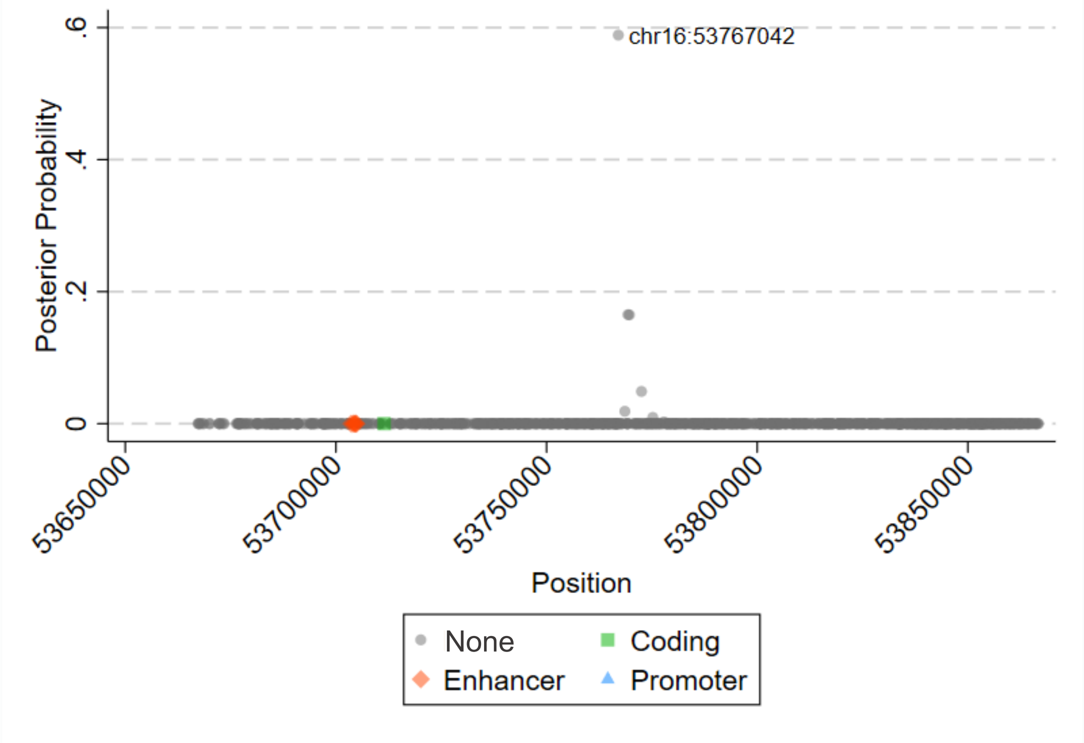

**E) rs6567160 (chr18:60161902), *MC4R***

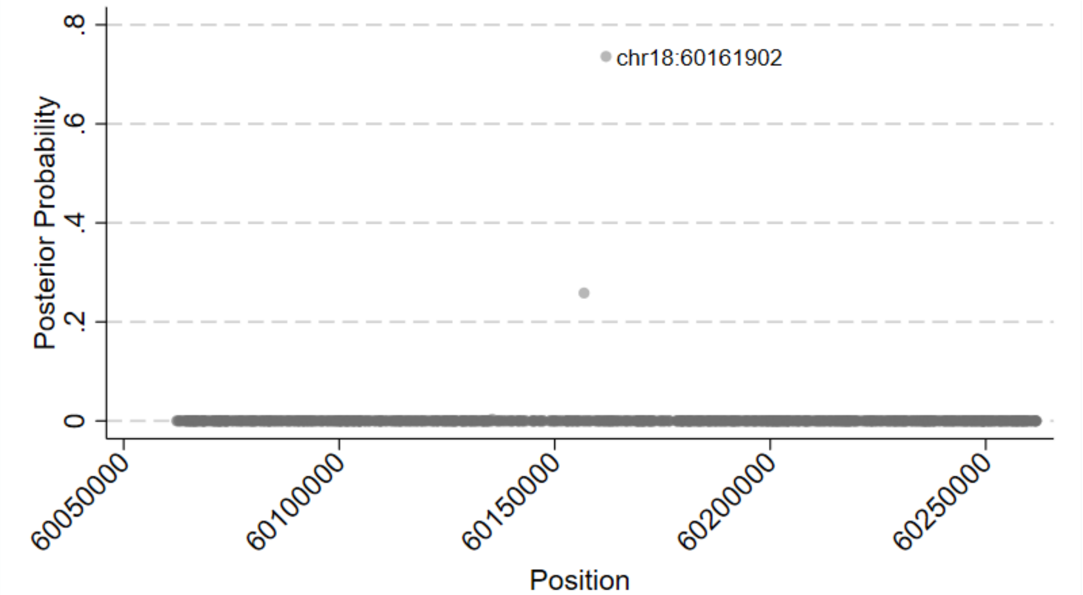


**F) rs55731973 (chr19:47113120), *ZC3H4***

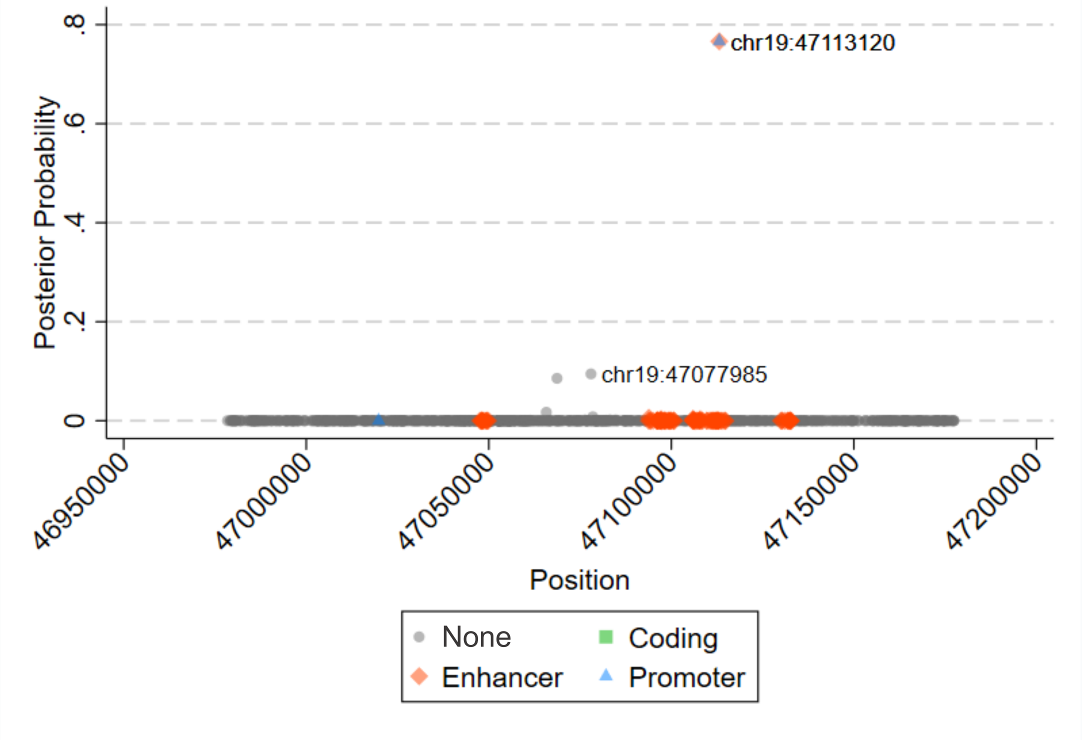

**G) rs1379871 (chrX:31836665), *DMD***

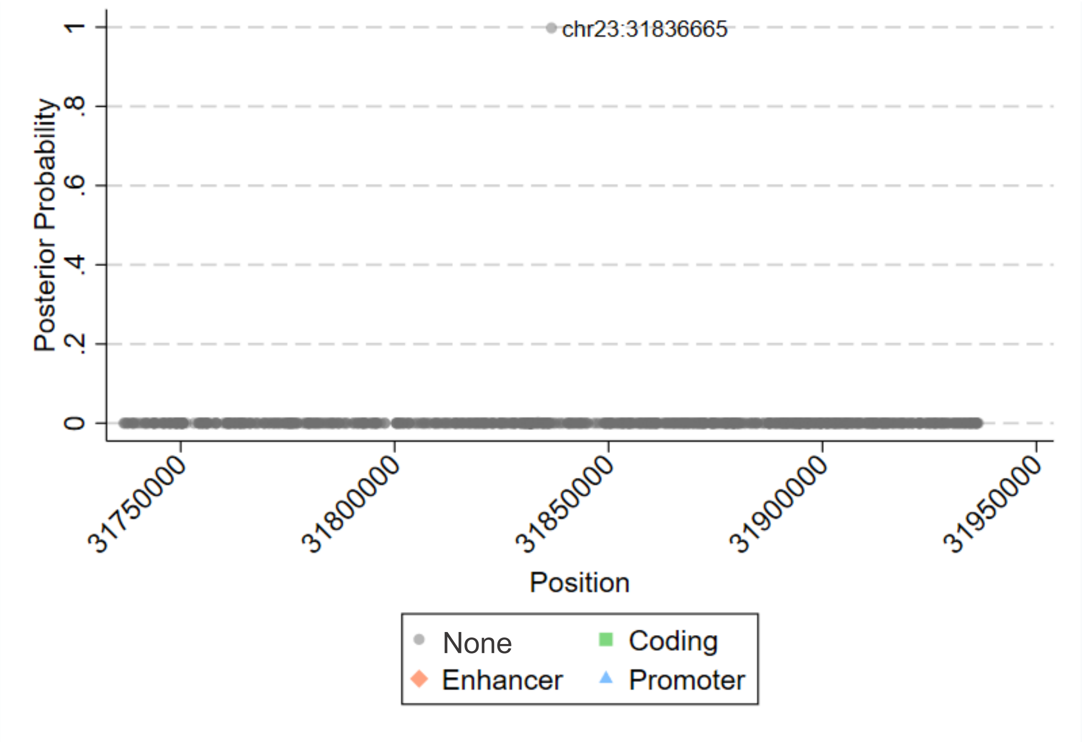

**Supplementary Figure 11. PheWAS meta-analysis Manhattan plot**

Manhattan plot of the PheWAS meta-analysis results. The red line indicates phenome-wide significance threshold ($P < 0.05/538$ PheCodes $= 9.3 \times 10^{-5}$), and the blue line indicates suggestive significance ($P < 0.001$). Only suggestively significant PheCodes are annotate with their phenotype. Arrow indicates direction of effect.

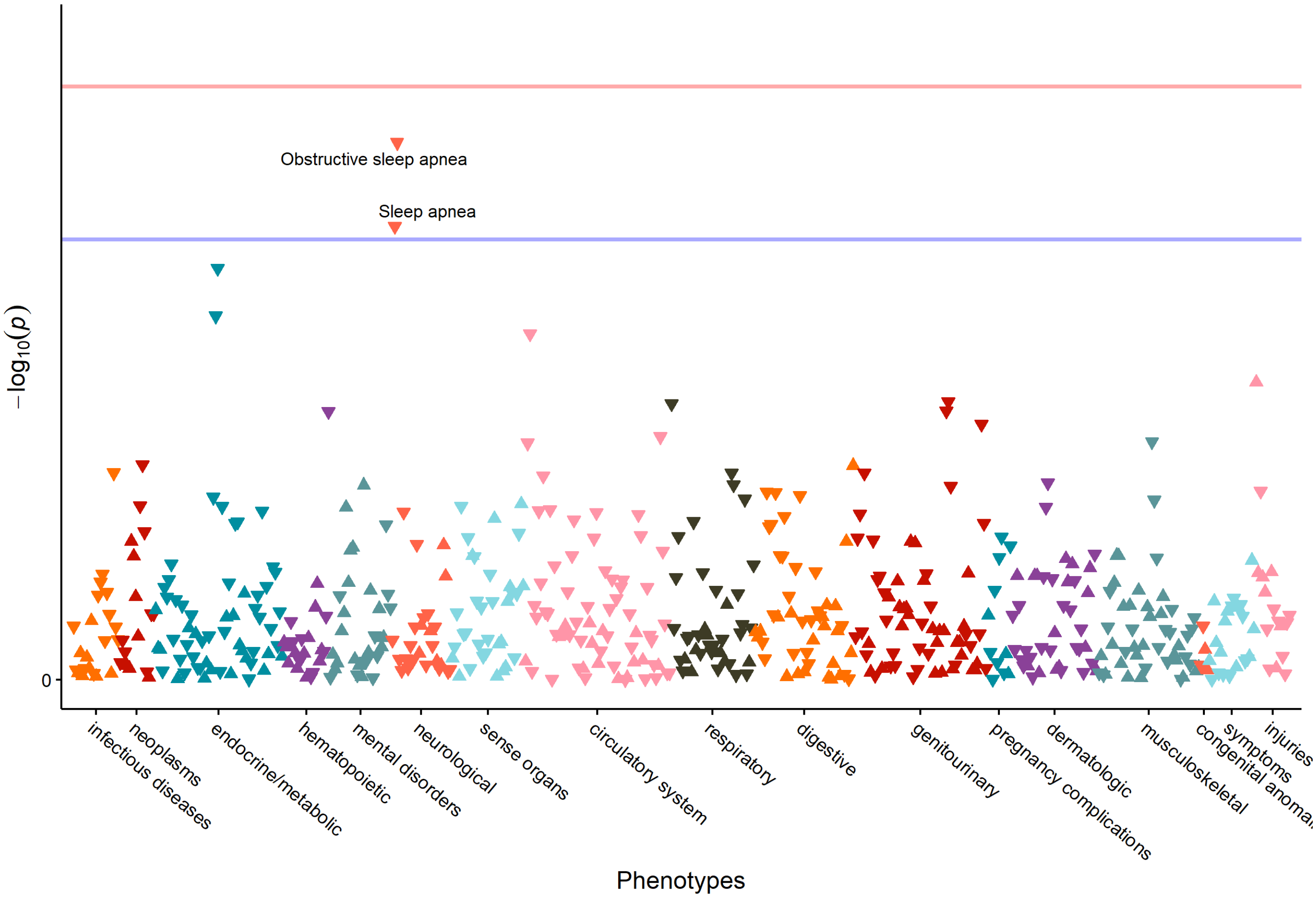

**Supplementary Data 1. Participant counts by study and population group**

| Study | Population groups | | | | | | | | | | | | | | | Total | Individuals with imputed population membership by study, n (%) |
|---|---|---|---|---|---|---|---|---|---|---|---|---|---|---|---|---|---|
| | **African** | **Amish** | **Asian** | **Barbadian** | **Central American** | **Costa Rican** | **Cuban** | **Dominican** | **European** | **Han Chinese** | **Mexican** | **Puerto Rican** | **Samoan** | **South American** | **Taiwanese** | | |
| Amish | 0 | 1106 | 0 | 0 | 0 | 0 | 0 | 0 | 0 | 0 | 0 | 0 | 0 | 0 | 0 | **1106** | 0 (0.0%) |
| ARIC | 1503 | 0 | 0 | 0 | 0 | 0 | 0 | 0 | 6158 | 0 | 0 | 0 | 0 | 0 | 0 | **7661** | 0 (0.0%) |
| BAGS | 0 | 0 | 0 | 248 | 0 | 0 | 0 | 0 | 0 | 0 | 0 | 0 | 0 | 0 | 0 | **248** | 0 (0.0%) |
| BioMe | 3138 | 0 | 408 | 0 | 161 | 0 | 149 | 763 | 3200 | 0 | 101 | 2595 | 0 | 289 | 0 | **10804** | 5248 (48.6%) |
| CARDIA | 1379 | 0 | 0 | 0 | 0 | 0 | 0 | 0 | 1684 | 0 | 0 | 0 | 0 | 0 | 0 | **3063** | 0 (0.0%) |
| CCAF | 0 | 0 | 0 | 0 | 0 | 0 | 1 | 0 | 360 | 0 | 0 | 0 | 0 | 0 | 0 | **361** | 4 (1.1%) |
| CFS | 493 | 0 | 2 | 0 | 0 | 0 | 0 | 0 | 450 | 0 | 1 | 2 | 0 | 0 | 0 | **948** | 14 (1.5%) |
| CHS | 711 | 0 | 0 | 0 | 0 | 0 | 0 | 0 | 2788 | 0 | 12 | 0 | 0 | 0 | 0 | **3511** | 53 (1.5%) |
| COPDGene | 3196 | 0 | 0 | 0 | 0 | 0 | 0 | 0 | 6661 | 0 | 0 | 0 | 0 | 0 | 0 | **9857** | 0 (0.0%) |
| CRA | 0 | 0 | 0 | 0 | 0 | 341 | 0 | 0 | 0 | 0 | 0 | 0 | 0 | 0 | 0 | **341** | 123 (36.1%) |
| DHS (AA CAC) | 384 | 0 | 0 | 0 | 0 | 0 | 0 | 0 | 0 | 0 | 0 | 0 | 0 | 0 | 0 | **384** | 0 (0.0%) |
| FHS | 4 | 0 | 1 | 0 | 0 | 0 | 1 | 0 | 4092 | 0 | 0 | 0 | 0 | 0 | 0 | **4098** | 480 (11.7%) |
| GALAII | 0 | 0 | 0 | 0 | 29 | 0 | 3 | 1 | 2 | 0 | 175 | 130 | 0 | 4 | 0 | **344** | 32 (9.3%) |
| GeneSTAR | 777 | 0 | 0 | 0 | 0 | 0 | 0 | 0 | 971 | 0 | 0 | 0 | 0 | 0 | 0 | **1748** | 0 (0.0%) |
| GENOA | 1193 | 0 | 0 | 0 | 0 | 0 | 0 | 0 | 0 | 0 | 0 | 0 | 0 | 0 | 0 | **1193** | 0 (0.0%) |
| GenSalt | 0 | 0 | 0 | 0 | 0 | 0 | 0 | 0 | 0 | 1787 | 0 | 0 | 0 | 0 | 0 | **1787** | 0 (0.0%) |
| GOLDN | 0 | 0 | 0 | 0 | 0 | 0 | 0 | 0 | 914 | 0 | 0 | 0 | 0 | 0 | 0 | **914** | 0 (0.0%) |
| HCHS/SOL | 23 | 0 | 0 | 0 | 509 | 0 | 1899 | 1124 | 33 | 0 | 1728 | 2073 | 0 | 297 | 0 | **7686** | 293 (3.8%) |
| HVH | 26 | 0 | 8 | 0 | 0 | 0 | 0 | 0 | 650 | 0 | 3 | 2 | 0 | 0 | 0 | **689** | 24 (3.5%) |
| HyperGEN | 1838 | 0 | 1 | 0 | 0 | 0 | 0 | 0 | 0 | 0 | 0 | 0 | 0 | 0 | 0 | **1839** | 0 (0.0%) |
| JHS | 3121 | 0 | 0 | 0 | 0 | 0 | 0 | 0 | 0 | 0 | 0 | 0 | 0 | 0 | 0 | **3121** | 0 (0.0%) |
| LTRC | 83 | 0 | 4 | 0 | 0 | 0 | 0 | 0 | 1276 | 0 | 12 | 8 | 0 | 0 | 0 | **1383** | 51 (3.7%) |
| Mayo_VTE | 3 | 0 | 0 | 0 | 0 | 0 | 1 | 0 | 920 | 0 | 5 | 1 | 0 | 0 | 0 | **930** | 54 (5.8%) |
| MESA | 1846 | 0 | 603 | 0 | 64 | 0 | 35 | 144 | 1861 | 0 | 544 | 140 | 0 | 81 | 0 | **5318** | 14 (0.3%) |
| MGH_AF | 1 | 0 | 0 | 0 | 1 | 0 | 3 | 0 | 974 | 0 | 2 | 0 | 0 | 1 | 0 | **982** | 25 (2.5%) |
| OMG_SCD | 443 | 0 | 0 | 0 | 0 | 0 | 0 | 0 | 0 | 0 | 0 | 1 | 0 | 0 | 0 | **444** | 444 (100.0%) |
| Partners | 4 | 0 | 1 | 0 | 0 | 0 | 0 | 0 | 117 | 0 | 0 | 0 | 0 | 0 | 0 | **122** | 2 (1.6%) |
| SAFS | 5 | 0 | 2 | 0 | 0 | 0 | 0 | 0 | 23 | 0 | 1504 | 0 | 0 | 0 | 0 | **1534** | 914 (59.6%) |
| SAGE | 448 | 0 | 0 | 0 | 0 | 0 | 0 | 0 | 0 | 0 | 0 | 0 | 0 | 0 | 0 | **448** | 1 (0.2%) |
| Samoan | 0 | 0 | 0 | 0 | 0 | 0 | 0 | 0 | 0 | 0 | 0 | 0 | 1274 | 0 | 0 | **1274** | 0 (0.0%) |
| THRV | 0 | 0 | 0 | 0 | 0 | 0 | 0 | 0 | 0 | 0 | 0 | 0 | 0 | 0 | 2053 | **2053** | 0 (0.0%) |
| VAFAR | 0 | 0 | 0 | 0 | 0 | 0 | 0 | 0 | 173 | 0 | 0 | 0 | 0 | 0 | 0 | **173** | 0 (0.0%) |
| VU_AF | 46 | 0 | 5 | 0 | 1 | 0 | 1 | 0 | 1030 | 0 | 2 | 0 | 0 | 1 | 0 | **1086** | 6 (0.6%) |
| walk_PHaSST | 373 | 0 | 0 | 0 | 3 | 0 | 1 | 11 | 1 | 0 | 0 | 0 | 0 | 2 | 0 | **391** | 29 (7.4%) |
| WGHS | 0 | 0 | 0 | 0 | 0 | 0 | 0 | 0 | 111 | 0 | 0 | 0 | 0 | 0 | 0 | **111** | 0 (0.0%) |
| WHI | 1450 | 0 | 206 | 0 | 8 | 0 | 34 | 3 | 8985 | 0 | 176 | 39 | 0 | 20 | 0 | **10921** | 204 (1.9%) |
| **Total** | **22488** | **1106** | **1241** | **248** | **776** | **341** | **2128** | **2046** | **43434** | **1787** | **4265** | **4991** | **1274** | **695** | **2053** | **88873** | **8015 (9.0%)** |
| **Individuals with imputed population membership by population group, n (%)** | 1038 (4.6%) | 0 (0.0%) | 212 (17.1%) | 0 (0.0%) | 193 (24.9%) | 123 (36.1%) | 202 (9.5%) | 834 (40.8%) | 1249 (2.9%) | 0 (0.0%) | 1128 (26.4%) | 2712 (54.3%) | 0 (0.0%) | 324 (46.6%) | 0 (0.0%) | **8015 (9.0%)** | |

**Supplementary Data 2. BMI and percent female by study**

| Study | N | Women, N (%) | BMI, mean (SD) |
|---|---|---|---|
| Amish | 1106 | 552 (49.91) | 26.97 (4.65) |
| ARIC | 7661 | 4230 (55.21) | 27.35 (5.23) |
| BAGS | 248 | 141 (56.85) | 28.00 (6.87) |
| BioMe | 10804 | 6242 (57.77) | 29.12 (7.01) |
| CARDIA | 3063 | 1739 (56.77) | 24.45 (4.96) |
| CCAF | 361 | 69 (19.11) | 30.42 (6.34) |
| CFS | 948 | 521 (54.96) | 32.27 (8.75) |
| CHS | 3511 | 2045 (58.25) | 26.84 (4.73) |
| COPDGene | 9857 | 4608 (46.75) | 28.82 (6.26) |
| CRA | 341 | 197 (57.77) | 26.39 (4.30) |
| DHS (AA CAC) | 384 | 220 (57.29) | 33.92 (7.79) |
| FHS | 4098 | 2224 (54.27) | 25.82 (4.83) |
| GALAII | 344 | 226 (65.70) | 27.47 (6.90) |
| GeneSTAR | 1748 | 1037 (59.32) | 29.62 (7.17) |
| GENOA | 1193 | 835 (69.99) | 31.08 (6.66) |
| GenSalt | 1787 | 843 (47.17) | 23.44 (3.12) |
| GOLDN | 914 | 481 (52.63) | 28.33 (5.75) |
| HCHS/SOL | 7686 | 4478 (58.26) | 30.11 (6.40) |
| HVH | 689 | 241 (34.98) | 31.86 (7.90) |
| HyperGEN | 1839 | 1165 (63.35) | 32.01 (7.75) |
| JHS | 3121 | 1952 (62.54) | 31.89 (7.31) |
| LTRC | 1383 | 664 (48.01) | 28.32 (5.91) |
| Mayo_VTE | 930 | 527 (56.67) | 31.29 (8.02) |
| MESA | 5318 | 2801 (52.67) | 28.51 (5.48) |
| MGH_AF | 982 | 202 (20.57) | 28.65 (5.56) |
| OMG_SCD | 444 | 243 (54.73) | 25.22 (6.84) |
| Partners | 122 | 43 (35.25) | 30.80 (7.42) |
| SAFS | 1534 | 899 (58.60) | 31.19 (7.41) |
| SAGE | 448 | 273 (60.94) | 28.11 (7.51) |
| Samoan | 1274 | 772 (60.60) | 33.75 (6.84) |
| THRV | 2053 | 1043 (50.80) | 24.76 (3.47) |
| VAFAR | 173 | 55 (31.79) | 32.70 (6.78) |
| VU_AF | 1086 | 300 (27.62) | 31.19 (6.85) |
| walk_PHaSST | 391 | 210 (53.71) | 24.83 (4.86) |
| WGHS | 111 | 111 (100.00) | 28.20 (6.11) |
| WHI | 10921 | 10920 (99.99) | 28.73 (5.97) |

**Supplementary Data 3. BMI and percent female by population group**

| Population group | N | Women, N (%) | BMI, mean (SD) |
|---|---|---|---|
| African | 22488 | 13903 (61.82) | 30.22 (7.32) |
| Amish | 1106 | 552 (49.91) | 26.97 (4.65) |
| Asian | 1241 | 711 (57.29) | 24.79 (4.42) |
| Barbadian | 248 | 141 (56.85) | 28.00 (6.87) |
| Central American | 776 | 466 (60.05) | 29.98 (6.26) |
| Costa Rican | 341 | 197 (57.77) | 26.39 (4.30) |
| Cuban | 2128 | 1119 (52.58) | 29.20 (5.84) |
| Dominican | 2046 | 1304 (63.73) | 29.05 (5.63) |
| European | 43434 | 26071 (60.02) | 27.76 (5.83) |
| Han Chinese | 1787 | 843 (47.17) | 23.44 (3.12) |
| Mexican | 4265 | 2555 (59.91) | 30.52 (6.72) |
| Puerto Rican | 4991 | 3027 (60.65) | 30.47 (6.95) |
| Samoan | 1274 | 772 (60.60) | 33.75 (6.84) |
| South American | 695 | 405 (58.27) | 28.73 (5.67) |
| Taiwanese | 2053 | 1043 (50.80) | 24.76 (3.47) |

**Supplementary Data 4. Study-specific descriptive statistics of age and BMI**

| Study | Population group | Trait | Men | | | | | | Women | | | | | |
|---|---|---|---|---|---|---|---|---|---|---|---|---|---|---|
| | | | N | Mean | SD | Median | Min | Max | N | Mean | SD | Median | Min | Max |
| **Discovery studies** | | | | | | | | | | | | | | |
| Amish | Amish | Age (years) | 554 | 49.39 | 17.29 | 49 | 20 | 90 | 552 | 51.63 | 16.29 | 53 | 20 | 90 |
| | | BMI (kg/m²) | 554 | 26.16 | 3.55 | 25.93 | 18.43 | 43.6 | 552 | 27.78 | 5.42 | 27.47 | 16.91 | 46.85 |
| ARIC | African | Age (years) | 608 | 53.95 | 6.07 | 53 | 44 | 66 | 895 | 53.85 | 5.89 | 53 | 45 | 65 |
| | | BMI (kg/m²) | 608 | 27.7 | 4.98 | 27.05 | 15.65 | 52.25 | 895 | 31.01 | 6.96 | 29.6 | 14.17 | 65.77 |
| ARIC | European | Age (years) | 2823 | 54.43 | 5.7 | 54 | 44 | 66 | 3335 | 53.71 | 5.74 | 53 | 44 | 66 |
| | | BMI (kg/m²) | 2823 | 27.31 | 3.91 | 26.81 | 16.07 | 53.75 | 3335 | 26.34 | 5.27 | 25.26 | 14.35 | 54.61 |
| BAGS | Barbadian | Age (years) | 107 | 40.84 | 12.66 | 43 | 18 | 74 | 141 | 38.19 | 10.83 | 39 | 18 | 72 |
| | | BMI (kg/m²) | 107 | 26.34 | 4.95 | 25.48 | 15.86 | 47.55 | 141 | 29.26 | 7.81 | 28.69 | 14.84 | 59.53 |
| BioMe | Asian | Age (years) | 212 | 53.12 | 13.47 | 54 | 23 | 81 | 196 | 46.77 | 14.42 | 44.5 | 21 | 91 |
| | | BMI (kg/m²) | 212 | 26.59 | 5.51 | 25.43 | 14.35 | 49.46 | 196 | 24.19 | 4.61 | 23.27 | 17.01 | 43.41 |
| BioMe | African | Age (years) | 1074 | 52.06 | 12.63 | 51 | 19 | 90 | 2064 | 53.05 | 13.17 | 53 | 19 | 92 |
| | | BMI (kg/m²) | 1074 | 28.63 | 6.36 | 27.79 | 15.29 | 65.61 | 2064 | 31.9 | 8.68 | 30.42 | 13.08 | 78.78 |
| BioMe | Central American | Age (years) | 73 | 52.58 | 14.4 | 54 | 19 | 77 | 88 | 52.45 | 13.99 | 55 | 23 | 77 |
| | | BMI (kg/m²) | 73 | 28.59 | 5.34 | 28.19 | 17.54 | 45.33 | 88 | 30.16 | 5.51 | 29 | 17.95 | 52.48 |
| BioMe | Cuban | Age (years) | 76 | 59.08 | 12.92 | 59 | 27 | 84 | 73 | 57.96 | 14.01 | 58 | 27 | 89 |
| | | BMI (kg/m²) | 76 | 29.56 | 5.44 | 28.32 | 20.8 | 44.94 | 73 | 27.28 | 6.45 | 25.13 | 17.14 | 47.26 |
| BioMe | Dominican | Age (years) | 290 | 61.16 | 13.47 | 63 | 24 | 97 | 473 | 54.34 | 13.31 | 55 | 19 | 89 |
| | | BMI (kg/m²) | 290 | 27.92 | 4.98 | 27.41 | 16.17 | 52.97 | 473 | 29.27 | 5.79 | 28.26 | 18.16 | 58.31 |
| BioMe | Mexican | Age (years) | 45 | 43.87 | 10.64 | 42 | 27 | 77 | 56 | 43.88 | 10.2 | 42 | 31 | 68 |
| | | BMI (kg/m²) | 45 | 28.64 | 4.28 | 29.07 | 21.45 | 37.48 | 56 | 27.89 | 3.97 | 27.92 | 19.31 | 36.33 |
| BioMe | Puerto Rican | Age (years) | 962 | 55.02 | 12.5 | 55 | 25 | 87 | 1633 | 55.22 | 13.63 | 55 | 19 | 97 |
| | | BMI (kg/m²) | 962 | 29.36 | 6.65 | 28.22 | 13.31 | 86.81 | 1633 | 31.05 | 7.11 | 30.17 | 14.26 | 68.02 |
| BioMe | South American | Age (years) | 137 | 57.76 | 13.24 | 58 | 25 | 91 | 152 | 54.38 | 14.54 | 56.5 | 24 | 83 |
| | | BMI (kg/m²) | 137 | 28.85 | 4.99 | 28.35 | 19.85 | 53.11 | 152 | 29.42 | 7 | 28.23 | 19.42 | 70.57 |
| BioMe | European | Age (years) | 1693 | 59.47 | 14.63 | 60 | 19 | 97 | 1507 | 55.31 | 14.86 | 56 | 19 | 90 |
| | | BMI (kg/m²) | 1693 | 27.96 | 5.33 | 27.01 | 15.46 | 63.11 | 1507 | 25.97 | 6.08 | 24.57 | 14.15 | 57.08 |
| CARDIA | African | Age (years) | 550 | 24.15 | 3.65 | 24 | 18 | 30 | 829 | 24.49 | 3.82 | 25 | 18 | 31 |
| | | BMI (kg/m²) | 550 | 24.63 | 4.3 | 23.94 | 15.76 | 44.11 | 829 | 26.04 | 6.56 | 24.31 | 14.51 | 53.42 |
| CARDIA | European | Age (years) | 774 | 25.6 | 3.27 | 26 | 18 | 30 | 910 | 25.59 | 3.32 | 26 | 18 | 30 |
| | | BMI (kg/m²) | 774 | 24.35 | 3.54 | 23.76 | 16.81 | 43.17 | 910 | 22.98 | 4.17 | 21.96 | 16.28 | 46.84 |
| CCAF | Cuban and European* | Age (years) | 292 | 54.34 | 9.02 | 56 | 20 | 76 | 69 | 55.83 | 10.24 | 58 | 22 | 73 |
| | | BMI (kg/m²) | 292 | 30.47 | 5.96 | 29.02 | 20.95 | 70.17 | 69 | 30.24 | 7.79 | 28.65 | 16.48 | 50.2 |
| CFS | European | Age (years) | 216 | 45.81 | 14.9 | 44.5 | 18 | 81 | 234 | 43.49 | 14.3 | 41 | 18 | 84 |
| | | BMI (kg/m²) | 216 | 31.4 | 7.24 | 29.83 | 19.33 | 60.95 | 234 | 30.3 | 8.38 | 29.08 | 17.52 | 66.6 |
| CFS | non-Eurpean[1] | Age (years) | 211 | 43.14 | 14.95 | 42 | 18 | 81 | 287 | 44.23 | 15.13 | 43 | 18 | 86 |
| | | BMI (kg/m²) | 211 | 31.83 | 8.28 | 30.92 | 12.71 | 58.86 | 287 | 34.86 | 9.78 | 32.72 | 16.09 | 84.8 |
| CHS | European | Age (years) | 1196 | 73.06 | 5.55 | 72 | 65 | 94 | 1592 | 72.24 | 5.17 | 71 | 65 | 98 |
| | | BMI (kg/m²) | 1196 | 26.4 | 3.69 | 26.05 | 15.61 | 46.23 | 1592 | 26.31 | 4.8 | 25.75 | 14.65 | 48.05 |
| CHS | non-European[2] | Age (years) | 270 | 72.42 | 5.52 | 71 | 65 | 92 | 453 | 72.89 | 5.62 | 72 | 65 | 93 |
| | | BMI (kg/m²) | 270 | 27.21 | 4.27 | 26.84 | 16.12 | 44.17 | 453 | 29.64 | 6.04 | 29.12 | 16.34 | 58.79 |
| COPDGene | African | Age (years) | 1765 | 54.41 | 6.84 | 53.1 | 39.9 | 80.8 | 1431 | 55.13 | 7.71 | 53.4 | 42.4 | 80.7 |
| | | BMI (kg/m²) | 1765 | 27.71 | 5.71 | 26.76 | 14.78 | 55.27 | 1431 | 30.82 | 7.42 | 30.12 | 12.67 | 64.1 |
| COPDGene | European | Age (years) | 3484 | 62.35 | 8.83 | 62.5 | 45 | 81 | 3177 | 61.64 | 8.87 | 62 | 45 | 85 |
| | | BMI (kg/m²) | 3484 | 28.85 | 5.52 | 28.09 | 13.75 | 58.65 | 3177 | 28.52 | 6.55 | 27.51 | 12.29 | 56.01 |
| CRA | Costa Rican | Age (years) | 144 | 41.15 | 15.02 | 38.91 | 18.11 | 91.62 | 197 | 39.3 | 13.53 | 36.62 | 18.07 | 72.07 |
| | | BMI (kg/m²) | 144 | 26.02 | 4.04 | 25.54 | 17.25 | 38.02 | 197 | 26.66 | 4.48 | 26.43 | 13.63 | 40.37 |
| DHS (AA CAC) | African | Age (years) | 164 | 60.01 | 8.59 | 60.5 | 39 | 79 | 220 | 59.42 | 8.81 | 60 | 36 | 86 |
| | | BMI (kg/m²) | 164 | 31.59 | 6.97 | 30.85 | 17.6 | 63.27 | 220 | 35.66 | 7.93 | 35.04 | 20.91 | 64.82 |
| FHS | African, Cuban, European* | Age (years) | 1874 | 38.97 | 9.95 | 38 | 18 | 83 | 2224 | 38.12 | 9.74 | 37 | 18 | 79 |
| | | BMI (kg/m²) | 1874 | 27.13 | 4.14 | 26.62 | 16.91 | 52.15 | 2224 | 24.72 | 5.08 | 23.44 | 16.34 | 58.08 |
| GALAII | Mexican | Age (years) | 57 | 19.46 | 1.04 | 19 | 18 | 22 | 118 | 19.83 | 1.19 | 19.8 | 18.01 | 21.97 |

| | | | | | | | | | | | | | | |
|---|---|---|---|---|---|---|---|---|---|---|---|---|---|---|
| GALAII | Mexican | BMI (kg/m²) | 57 | 28.09 | 7.23 | 26.5 | 17.2 | 52.2 | 118 | 27.77 | 6.93 | 26.5 | 17.8 | 48.1 |
| GALAII | Puerto Rican | Age (years) | 48 | 19.64 | 0.9 | 19.79 | 18 | 21.21 | 82 | 19.34 | 0.94 | 19.14 | 18.02 | 21.86 |
| | | BMI (kg/m²) | 48 | 27.2 | 6.22 | 25.9 | 17.6 | 44.4 | 82 | 26.75 | 7.3 | 25 | 17.5 | 67.5 |
| GALAII | Other[3] | Age (years) | 13 | 19.67 | 1.06 | 19.75 | 18.21 | 21.46 | 26 | 19.9 | 1.17 | 19.75 | 18.05 | 21.99 |
| | | BMI (kg/m²) | 13 | 27.73 | 5.41 | 29.1 | 19.5 | 38.51 | 26 | 27.44 | 6.99 | 27.15 | 18.7 | 50.18 |
| GeneSTAR | African | Age (years) | 286 | 40.73 | 10.78 | 41 | 21 | 66 | 491 | 41.06 | 10.82 | 41 | 21 | 75 |
| | | BMI (kg/m²) | 286 | 29.06 | 6.44 | 27.87 | 16.89 | 53.48 | 491 | 32.44 | 8.4 | 31.18 | 16.41 | 81.19 |
| GeneSTAR | European | Age (years) | 425 | 40.37 | 10.87 | 41 | 21 | 75 | 546 | 43.29 | 12.32 | 43 | 21 | 79 |
| | | BMI (kg/m²) | 425 | 28.72 | 5.1 | 27.81 | 16.5 | 51.13 | 546 | 28.08 | 7.01 | 26.35 | 16.99 | 61.83 |
| GENOA | African | Age (years) | 358 | 57.37 | 10.2 | 56.7 | 29.4 | 86.1 | 835 | 56.46 | 10.76 | 56.7 | 21 | 90.3 |
| | | BMI (kg/m²) | 358 | 28.4 | 4.76 | 28.24 | 15.24 | 50.32 | 835 | 32.23 | 7.02 | 31.52 | 17.58 | 61.37 |
| GenSalt | Han Chinese | Age (years) | 944 | 39.75 | 8.99 | 39 | 18 | 62 | 843 | 38.71 | 8.84 | 39 | 18 | 59 |
| | | BMI (kg/m²) | 944 | 23.25 | 3.08 | 22.77 | 16.38 | 33.82 | 843 | 23.66 | 3.16 | 23.38 | 15.87 | 37.77 |
| GOLDN | European | Age (years) | 433 | 48.42 | 16.3 | 48 | 18 | 88 | 481 | 47.86 | 16.13 | 47 | 18 | 87 |
| | | BMI (kg/m²) | 433 | 28.46 | 4.9 | 27.89 | 17.13 | 50.73 | 481 | 28.2 | 6.42 | 27.38 | 16.6 | 52.66 |
| HCHS/SOL | Central American | Age (years) | 201 | 43.27 | 13.95 | 44 | 18 | 74 | 308 | 45.69 | 13.4 | 47 | 18 | 74 |
| | | BMI (kg/m²) | 201 | 29.61 | 5.74 | 28.89 | 17.34 | 47.3 | 308 | 30.87 | 7.06 | 29.58 | 16.64 | 62.37 |
| HCHS/SOL | Cuban | Age (years) | 911 | 49.33 | 13.19 | 51 | 18 | 74 | 988 | 48.91 | 12.97 | 50 | 18 | 75 |
| | | BMI (kg/m²) | 911 | 28.75 | 5.18 | 28.33 | 14.9 | 50.91 | 988 | 29.81 | 6.37 | 29.03 | 14.28 | 63.78 |
| HCHS/SOL | Dominican | Age (years) | 386 | 45.01 | 14.78 | 47 | 18 | 75 | 738 | 45.74 | 13.98 | 47 | 18 | 74 |
| | | BMI (kg/m²) | 386 | 28.61 | 4.89 | 28.61 | 17.3 | 55.3 | 738 | 29.91 | 6.15 | 29.13 | 15.23 | 60.57 |
| HCHS/SOL | Mexican | Age (years) | 680 | 42.76 | 14.26 | 44 | 18 | 74 | 1048 | 44.91 | 13.72 | 46 | 18 | 76 |
| | | BMI (kg/m²) | 680 | 29.7 | 5.5 | 28.85 | 16.33 | 52.94 | 1048 | 31.28 | 7.14 | 29.86 | 17.73 | 70.35 |
| HCHS/SOL | Puerto Rican | Age (years) | 884 | 46.25 | 14.43 | 48 | 18 | 74 | 1189 | 48.4 | 14.07 | 50 | 18 | 75 |
| | | BMI (kg/m²) | 884 | 29.53 | 6.36 | 28.56 | 17.67 | 55.87 | 1189 | 31.8 | 7.18 | 30.82 | 15.62 | 67.7 |
| HCHS/SOL | South American | Age (years) | 116 | 44.93 | 12.99 | 46.5 | 18 | 70 | 181 | 47.49 | 13.09 | 48 | 18 | 76 |
| | | BMI (kg/m²) | 116 | 28.24 | 4.76 | 28 | 17.12 | 42.65 | 181 | 29.33 | 6.06 | 28.52 | 19.5 | 49.93 |
| HCHS/SOL | Other[4] | Age (years) | 30 | 47.93 | 14.36 | 48.5 | 21 | 74 | 26 | 45.12 | 17.59 | 42 | 20 | 74 |
| | | BMI (kg/m²) | 30 | 28.75 | 6.28 | 27.27 | 17.65 | 48.49 | 26 | 31.94 | 8.48 | 31.39 | 19.04 | 61.32 |
| HVH | European | Age (years) | 423 | 61.36 | 11.1 | 60 | 31 | 89 | 227 | 63.86 | 13.67 | 64 | 21 | 90 |
| | | BMI (kg/m²) | 423 | 30.89 | 6.69 | 29.53 | 17.07 | 74.34 | 227 | 33.34 | 8.99 | 31.64 | 18.71 | 67.49 |
| HVH | non-European[5] | Age (years) | 25 | 60 | 13.69 | 59 | 35 | 88 | 14 | 51.21 | 12.32 | 49.5 | 23 | 69 |
| | | BMI (kg/m²) | 25 | 31.28 | 10.45 | 29.6 | 20.68 | 76.05 | 14 | 38.36 | 11.28 | 35.26 | 23.78 | 64.73 |
| HyperGEN | African and Asian* | Age (years) | 674 | 46.42 | 12.43 | 46 | 18 | 85 | 1165 | 47.01 | 13 | 47 | 18 | 84 |
| | | BMI (kg/m²) | 674 | 29.6 | 6.31 | 28.55 | 16.51 | 56.66 | 1165 | 33.41 | 8.15 | 32.13 | 16.18 | 73.68 |
| JHS | African | Age (years) | 1169 | 53.58 | 12.91 | 54 | 21 | 89 | 1952 | 54.69 | 12.78 | 55 | 20 | 91 |
| | | BMI (kg/m²) | 1169 | 30.06 | 6.31 | 29.03 | 16.35 | 66.09 | 1952 | 32.99 | 7.65 | 31.92 | 16.02 | 91.8 |
| LTRC | European | Age (years) | 665 | 64.09 | 10.44 | 65 | 25 | 87 | 611 | 62.65 | 10.55 | 64 | 21 | 88 |
| | | BMI (kg/m²) | 665 | 28.74 | 4.97 | 28.39 | 16.37 | 58.63 | 611 | 27.74 | 6.61 | 27.17 | 13.28 | 52.28 |
| LTRC | non-European[6] | Age (years) | 54 | 61.3 | 9.66 | 61 | 40 | 83 | 53 | 56.77 | 12.25 | 57 | 24 | 80 |
| | | BMI (kg/m²) | 54 | 27.17 | 5.85 | 26.57 | 17.37 | 40 | 53 | 30.77 | 7.16 | 29.62 | 19.18 | 51.01 |
| Mayo_VTE | African, Cuban, European, Mexican, Puerto Rican* | Age (years) | 403 | 58.47 | 15.33 | 60 | 18 | 91 | 527 | 52.42 | 17.05 | 53 | 19 | 95 |
| | | BMI (kg/m²) | 403 | 31.13 | 6.6 | 29.94 | 18.21 | 70.53 | 527 | 31.42 | 8.95 | 29.71 | 17.14 | 69.63 |
| MESA | Asian | Age (years) | 302 | 61.44 | 10.2 | 61 | 44 | 84 | 301 | 60.95 | 10.11 | 60 | 44 | 83 |
| | | BMI (kg/m²) | 302 | 24.19 | 3.09 | 23.86 | 16.09 | 33.48 | 301 | 23.96 | 3.31 | 23.82 | 16.63 | 35.35 |
| MESA | African | Age (years) | 815 | 59.43 | 9.61 | 58 | 39 | 84 | 1031 | 60.13 | 8.98 | 59 | 44 | 91 |
| | | BMI (kg/m²) | 815 | 29 | 4.84 | 28.61 | 17.4 | 52.46 | 1031 | 31.07 | 6.25 | 30.12 | 15.68 | 53.45 |
| MESA | Central American | Age (years) | 25 | 56.2 | 6.95 | 56 | 46 | 68 | 39 | 58.46 | 8.87 | 59 | 46 | 77 |
| | | BMI (kg/m²) | 25 | 28.73 | 4.73 | 27.96 | 23.01 | 45.64 | 39 | 30.33 | 5.38 | 30.41 | 20.44 | 47.46 |
| MESA | Cuban | Age (years) | 17 | 65.76 | 11.62 | 68 | 45 | 81 | 18 | 70.06 | 8.13 | 70 | 51 | 82 |
| | | BMI (kg/m²) | 17 | 27.87 | 4.26 | 26.41 | 22.02 | 36.24 | 18 | 28.71 | 6.21 | 27.36 | 21.39 | 46.82 |
| MESA | Dominican | Age (years) | 63 | 57.4 | 9.65 | 55 | 45 | 80 | 81 | 59.33 | 9.64 | 59 | 45 | 79 |
| | | BMI (kg/m²) | 63 | 27.4 | 3.85 | 26.89 | 18.91 | 39.34 | 81 | 28.17 | 4.47 | 27.48 | 19.95 | 38.5 |
| MESA | Mexican | Age (years) | 283 | 60.7 | 10.16 | 60 | 44 | 84 | 261 | 60.28 | 9.64 | 60 | 44 | 82 |
| | | BMI (kg/m²) | 283 | 29.33 | 4.46 | 28.83 | 17.57 | 46.28 | 261 | 30.98 | 5.99 | 30.35 | 18.84 | 52.48 |

| | | | | | | | | | | | | | | |
|---|---|---|---|---|---|---|---|---|---|---|---|---|---|---|
| MESA | Puerto Rican | Age (years) | 63 | 57.9 | 8.67 | 56 | 45 | 79 | 77 | 59.45 | 9.37 | 57 | 45 | 81 |
| | | BMI (kg/m²) | 63 | 28.8 | 4.21 | 28.75 | 21.89 | 43.9 | 77 | 30.73 | 6.51 | 28.67 | 20.13 | 51.56 |
| MESA | South American | Age (years) | 34 | 60.68 | 9.85 | 60 | 45 | 80 | 47 | 61.83 | 11.1 | 61 | 45 | 84 |
| | | BMI (kg/m²) | 34 | 27.2 | 2.87 | 27.05 | 22.92 | 33.95 | 47 | 27.93 | 4.67 | 27.77 | 18.3 | 39.61 |
| MESA | European | Age (years) | 915 | 61.63 | 9.67 | 62 | 45 | 83 | 946 | 61.4 | 9.95 | 61 | 44 | 84 |
| | | BMI (kg/m²) | 915 | 27.98 | 3.98 | 27.61 | 19.01 | 42.41 | 946 | 27.59 | 5.81 | 26.52 | 16.87 | 48.99 |
| MGH_AF | African, Central American, Cuban, European, Mexican, South American* | Age (years) | 780 | 53.81 | 10.69 | 55 | 19 | 82 | 202 | 56.77 | 10.27 | 58 | 18 | 80 |
| | | BMI (kg/m²) | 780 | 28.8 | 5.11 | 27.88 | 15.2 | 57.5 | 202 | 28.07 | 7.03 | 26.56 | 17.14 | 54.73 |
| OMG_SCD | African and Puerto Rican* | Age (years) | 201 | 31.68 | 11.95 | 29 | 18 | 84 | 243 | 34.56 | 12.36 | 33 | 18 | 70 |
| | | BMI (kg/m²) | 201 | 24.22 | 5.88 | 23.2 | 13.8 | 59 | 243 | 26.04 | 7.45 | 24.3 | 16 | 58 |
| Partners | African, Asian, European* | Age (years) | 79 | 48.46 | 9.47 | 51 | 23 | 60 | 43 | 50.16 | 8.97 | 53 | 21 | 63 |
| | | BMI (kg/m²) | 79 | 30.78 | 7.16 | 29.29 | 21.46 | 59.37 | 43 | 30.84 | 7.96 | 29.65 | 19.27 | 53.59 |
| SAFS | Mexican | Age (years) | 620 | 42.14 | 17.28 | 40 | 18 | 90 | 884 | 44.74 | 16.74 | 44 | 18 | 97 |
| | | BMI (kg/m²) | 620 | 30.32 | 7.01 | 29.59 | 14.36 | 65.62 | 884 | 31.8 | 7.63 | 31.07 | 15.55 | 70.2 |
| SAFS | non-Mexican[7] | Age (years) | 15 | 38.27 | 17.51 | 31 | 18 | 75 | 15 | 37.8 | 13.8 | 32 | 20 | 66 |
| | | BMI (kg/m²) | 15 | 28.99 | 6.36 | 30.02 | 19.5 | 39.67 | 15 | 33.72 | 7.68 | 33 | 20.81 | 49.13 |
| SAGE | African | Age (years) | 175 | 20.87 | 4.45 | 19.7 | 18 | 40.9 | 273 | 22.2 | 5.91 | 20 | 18 | 40.7 |
| | | BMI (kg/m²) | 175 | 26.55 | 6.22 | 24.5 | 17.5 | 49.2 | 273 | 29.11 | 8.09 | 27 | 16.26 | 56.9 |
| Samoan | Samoan | Age (years) | 502 | 45.04 | 11.54 | 46.13 | 24.62 | 64.96 | 772 | 44.2 | 11.16 | 43.78 | 24.53 | 65 |
| | | BMI (kg/m²) | 502 | 31.37 | 5.83 | 30.75 | 18.56 | 54.06 | 772 | 35.3 | 7 | 35.04 | 18.05 | 61.07 |
| THRV | Taiwanese | Age (years) | 1010 | 52.25 | 9.95 | 51 | 29 | 86 | 1043 | 51.71 | 9.63 | 51 | 18 | 86 |
| | | BMI (kg/m²) | 1010 | 25.35 | 3.35 | 25.24 | 15.87 | 42.45 | 1043 | 24.19 | 3.49 | 23.64 | 15.37 | 35.82 |
| VAFAR | European | Age (years) | 118 | 57.07 | 8.63 | 58.5 | 32.53 | 78.21 | 55 | 60.49 | 8.22 | 61.54 | 25.68 | 74.78 |
| | | BMI (kg/m²) | 118 | 32.27 | 6.15 | 31.41 | 22.2 | 56.92 | 55 | 33.62 | 7.94 | 32.74 | 21.11 | 53.26 |
| VU_AF | European | Age (years) | 750 | 53.17 | 10.69 | 55 | 19 | 80 | 280 | 53.72 | 11.76 | 56.5 | 18 | 81 |
| | | BMI (kg/m²) | 750 | 31.1 | 6.42 | 30.12 | 16.81 | 69.64 | 280 | 31.27 | 7.63 | 30.1 | 18.79 | 55.76 |
| VU_AF | non-European[8] | Age (years) | 36 | 45.47 | 11.12 | 48 | 21 | 65 | 20 | 48.6 | 10.06 | 48 | 29 | 63 |
| | | BMI (kg/m²) | 36 | 31.36 | 8.36 | 29.79 | 20.36 | 66.29 | 20 | 32.75 | 8.16 | 30.57 | 22.08 | 52.87 |
| walk_PHaSST | African, Central American, Cuban, Domonican, European, South American* | Age (years) | 181 | 35.59 | 12.23 | 33.2 | 18.3 | 72 | 210 | 39.91 | 13.13 | 39.5 | 18.3 | 84 |
| | | BMI (kg/m²) | 181 | 23.77 | 4.28 | 22.84 | 17.09 | 38.67 | 210 | 25.75 | 5.15 | 24.97 | 15.24 | 45.23 |
| WGHS | European | Age (years) | n/a | n/a | n/a | n/a | n/a | n/a | 111 | 49.38 | 3.46 | 48 | 45 | 59 |
| | | BMI (kg/m²) | n/a | n/a | n/a | n/a | n/a | n/a | 111 | 28.2 | 6.11 | 27.37 | 18.88 | 57.61 |
| WHI | Asian | Age (years) | n/a | n/a | n/a | n/a | n/a | n/a | 206 | 66.49 | 7.27 | 67.24 | 50 | 79.04 |
| | | BMI (kg/m²) | n/a | n/a | n/a | n/a | n/a | n/a | 206 | 25.18 | 4.5 | 24.5 | 16.83 | 42.33 |
| WHI | African | Age (years) | n/a | n/a | n/a | n/a | n/a | n/a | 1450 | 63.58 | 7.04 | 63.21 | 50 | 80.84 |
| | | BMI (kg/m²) | n/a | n/a | n/a | n/a | n/a | n/a | 1450 | 31.55 | 6.19 | 30.63 | 17.73 | 60.91 |
| WHI | Cuban | Age (years) | n/a | n/a | n/a | n/a | n/a | n/a | 34 | 64 | 8.28 | 64.04 | 51.05 | 78.55 |
| | | BMI (kg/m²) | n/a | n/a | n/a | n/a | n/a | n/a | 34 | 28.78 | 4.98 | 28.35 | 19.35 | 44.11 |
| WHI | Mexican | Age (years) | n/a | n/a | n/a | n/a | n/a | n/a | 176 | 62.62 | 6.81 | 62.28 | 50.02 | 78.1 |
| | | BMI (kg/m²) | n/a | n/a | n/a | n/a | n/a | n/a | 176 | 29.07 | 5.37 | 28.16 | 18.63 | 45.55 |
| WHI | Puerto Rican | Age (years) | n/a | n/a | n/a | n/a | n/a | n/a | 39 | 62.74 | 6.39 | 63.05 | 51.09 | 76.01 |
| | | BMI (kg/m²) | n/a | n/a | n/a | n/a | n/a | n/a | 39 | 28.78 | 6.03 | 27.88 | 20.53 | 48.68 |
| WHI | South American | Age (years) | n/a | n/a | n/a | n/a | n/a | n/a | 20 | 63.73 | 7.84 | 64.53 | 50.18 | 75.2 |
| | | BMI (kg/m²) | n/a | n/a | n/a | n/a | n/a | n/a | 20 | 26.98 | 3.93 | 26.36 | 21.15 | 38.09 |
| WHI | European[9] | Age (years) | n/a | n/a | n/a | n/a | n/a | n/a | 8985 | 67.37 | 6.54 | 68.07 | 49.92 | 80.07 |
| | | BMI (kg/m²) | n/a | n/a | n/a | n/a | n/a | n/a | 8985 | 28.36 | 5.83 | 27.34 | 15.59 | 60.91 |
| WHI | Other[10] | Age (years) | n/a | n/a | n/a | n/a | n/a | n/a | 11 | 63.29 | 7.99 | 61.08 | 52.35 | 78.09 |
| | | BMI (kg/m²) | n/a | n/a | n/a | n/a | n/a | n/a | 11 | 27.02 | 7.62 | 24.98 | 20.84 | 47.59 |
| **TOTAL** | | **Age (years)** | **35764** | **52.16** | **14.98** | **53** | **18** | **97** | **53109** | **54.47** | **15.16** | **56** | **18** | **98** |
| | | **BMI (kg/m²)** | **35764** | **28.12** | **5.54** | **27.31** | **12.71** | **86.81** | **53109** | **28.98** | **7.02** | **27.73** | **12.29** | **91.8** |
| **Replication studies** | | | | | | | | | | | | | | |
| **Study** | **Population group** | **Trait** | **Men** | | | | | | **Women** | | | | | |
| | | | **N** | **Mean** | **SD** | **Median** | **Min** | **Max** | **N** | **Mean** | **SD** | **Median** | **Min** | **Max** |
| MEC | | Age (years) | 3281 | 59.72 | 7.4 | 61 | 44 | 70 | 3825 | 57.65 | 7.61 | 57 | 44 | 70 |
| | | BMI (kg/m²) | 3281 | 27.64 | 4.31 | 27.18 | 10.87 | 54.16 | 3825 | 29.5 | 6.25 | 28.38 | 13.42 | 68.73 |

| | | | | | | | | | | | | | | | |
|---|---|---|---|---|---|---|---|---|---|---|---|---|---|---|---|
| MVP | African, African American, Black | Age (years) | 69346 | 58.92 | 11.65 | 60 | 20 | 103 | | 10543 | 48.59 | 11.37 | 50 | 20 | 94 |
| | | BMI (kg/m²) | 69346 | 30.06 | 6.07 | 29.42 | 12.21 | 85.46 | | 10543 | 31.42 | 6.32 | 30.83 | 14.18 | 66.8 |
| BioMe | | Age (years) | 2492 | 48.63 | 13.92 | 49.56 | 18.01 | 83.24 | | 4146 | 48.95 | 15.1 | 49.32 | 18.02 | 84.04 |
| | | BMI (kg/m²) | 2492 | 28.47 | 6.55 | 27.46 | 14.13 | 69.09 | | 4146 | 31.89 | 8.66 | 30.61 | 13.08 | 89.14 |
| UKBB | | Age (years) | 3718 | 51.65 | 8.23 | 50 | 39 | 70 | | 5148 | 52.05 | 7.95 | 51 | 40 | 70 |
| | | BMI (kg/m²) | 3718 | 28.33 | 4.29 | 27.93 | 16.66 | 57.48 | | 5148 | 30.34 | 5.98 | 29.59 | 16.02 | 68.13 |
| REGARDS | | Age (years) | 3409 | 64.03 | 9.06 | 63 | 45 | 94 | | 5312 | 63.37 | 9.27 | 62 | 45 | 96 |
| | | BMI (kg/m²) | 3409 | 28.97 | 5.49 | 28.43 | 12.31 | 51.73 | | 5312 | 32.02 | 7.14 | 31.09 | 12.16 | 89.44 |

*BMI, body mass index; SD, standard deviation; n/a, not available*

*Combined because when stratified by sex, the number of participant is ≤ 5 in at least one of the population group.

1. CFS non-European: Collapsed African, Asian, Mexican, and Puerto Rican.
2. CHS non-European: Collapsed African and Mexican.
3. GALA II Other: Collapsed Central American, Cuban, Dominican, European, and South American.
4. HCHS_SOL Other: Collapsed African and European.
5. HVH non-European: Collapsed African, Asian, Mexican, and Puerto Rican.
6. LTRC non-European: Collapsed African, Asian, Mexican, and Puerto Rican.
7. SAFS non-Mexican: Collapsed African, Asian, and European.
8. VU_AF non-European: Collapsed African, Asian, South American, Central American, Cuban, and Mexican.
9. WHI European includes 1 participant who is biologically male.
10. WHI Other: Collapsed Central American and Dominican.

**Supplementary Data 5. Genome-wide significant variants by locus and ALT frequency by population group**

| | | | | | | | | | | | | | | ALT frequency by population group | | | | | | | | | | | | | | |
|---|---|---|---|---|---|---|---|---|---|---|---|---|---|---|---|---|---|---|---|---|---|---|---|---|---|---|---|---|
| **CHR** | **POS (hg38)** | **REF** | **ALT** | **ALT Freq** | **rsID** | **Beta** | **SE** | ***P*-value** | **PVE (%)** | **Known index variant[a]** | **OASIS annotaion** | **Nearest gene** | **Novel locus[b]** | **African** | **Amish** | **Asian** | **Barbadian** | **Central American** | **Costa Rican** | **Cuban** | **Dominican** | **European** | **Han Chinese** | **Mexican** | **Puerto Rican** | **Samoan** | **South American** | **Taiwanese** |
| 1 | 177920345 | A | G | 20% | rs543874 | 0.0639 | 0.0060 | 1.38E-26 | 0.128 | Yes | intergenic | *SEC16B* | No | 25% | 15% | 15% | 29% | 26% | 23% | 19% | 21% | 19% | 22% | 18% | 22% | 5% | 21% | 16% |
| 2 | 621558 | C | T | 85% | rs939584 | 0.0576 | 0.0068 | 1.99E-17 | 0.081 | No | intergenic | *TMEM18* | No | 12% | 18% | 10% | 12% | 12% | 8% | 15% | 14% | 18% | 11% | 11% | 16% | 1% | 12% | 8% |
| 2 | 24927427 | A | G | 56% | rs10182181 | 0.0350 | 0.0052 | 1.76E-11 | 0.051 | Yes | intergenic | *ADCY3* | No | 16% | 65% | 54% | 11% | 63% | 63% | 52% | 36% | 53% | 57% | 67% | 47% | 47% | 65% | 56% |
| 3 | 186108951 | T | G | 82% | rs869400 | 0.0383 | 0.0063 | 1.21E-09 | 0.042 | No | UTR5 | *ETV5* | No | 23% | 11% | 7% | 25% | 11% | 9% | 18% | 20% | 18% | 2% | 10% | 19% | 5% | 12% | 7% |
| 4 | 45179317 | A | T | 36% | rs12507026 | 0.0449 | 0.0051 | 9.55E-19 | 0.088 | Yes | intergenic | *GNPDA2* | No | 24% | 32% | 28% | 27% | 37% | 48% | 40% | 34% | 43% | 32% | 37% | 37% | 21% | 41% | 28% |
| 5 | 75707853 | T | C | 55% | rs2307111 | -0.0324 | 0.0053 | 7.43E-10 | 0.043 | Yes | exonic, missense | *POC5* | No | 15% | 55% | 43% | 10% | 53% | 52% | 52% | 38% | 59% | 42% | 61% | 46% | 55% | 54% | 44% |
| 6 | 50830813 | C | T | 19% | rs2206277 | 0.0543 | 0.0062 | 2.05E-18 | 0.086 | Novel | intronic | *TFAP2B* | No | 14% | 12% | 25% | 11% | 30% | 28% | 18% | 18% | 18% | 24% | 39% | 24% | 10% | 34% | 21% |
| 8 | 76068626 | A | G | 47% | rs830463 | 0.0305 | 0.0049 | 6.58E-10 | 0.043 | No | intergenic | *HNF4G* | No | 33% | 60% | 37% | 30% | 46% | 50% | 49% | 43% | 56% | 35% | 51% | 48% | 24% | 48% | 36% |
| 11 | 27657463 | GT | G | 58% | rs3838785 | -0.0303 | 0.0051 | 3.14E-09 | 0.040 | Novel | ncRNA_intronic, deletion | *BDNF* | No | 63% | 21% | 27% | 63% | 57% | 55% | 38% | 52% | 31% | 26% | 52% | 48% | 32% | 53% | 24% |
| 12 | 49853685 | G | A | 30% | rs7138803 | 0.0363 | 0.0054 | 1.69E-11 | 0.051 | Yes | intergenic | *BCDIN3D* | No | 18% | 40% | 29% | 17% | 22% | 15% | 31% | 23% | 38% | 29% | 24% | 26% | 10% | 23% | 27% |
| 13 | 53533448 | G | T | 14% | rs9568868 | 0.0472 | 0.0072 | 5.73E-11 | 0.048 | No | intergenic | *OLFM4* | No | 5% | 28% | 24% | 2% | 31% | 33% | 13% | 11% | 13% | 25% | 34% | 19% | 43% | 33% | 25% |
| 16 | 53767042 | T | C | 29% | rs1421085 | 0.0901 | 0.0056 | 6.11E-59 | 0.295 | Yes | intronic | *FTO* | No | 11% | 50% | 19% | 9% | 21% | 30% | 35% | 25% | 42% | 10% | 22% | 27% | 20% | 23% | 14% |
| 18 | 60161902 | T | C | 21% | rs6567160 | 0.0525 | 0.0059 | 8.22E-19 | 0.088 | Yes | intergenic | *MC4R* | No | 19% | 31% | 20% | 25% | 12% | 14% | 19% | 20% | 23% | 24% | 12% | 19% | 17% | 12% | 20% |
| 19 | 47077985 | C | T | 50% | rs28590228 | 0.0332 | 0.0053 | 4.75E-10 | 0.044 | No | intronic | *ZC3H4* | No | 20% | 66% | 33% | 18% | 48% | 59% | 58% | 40% | 68% | 29% | 57% | 46% | 68% | 51% | 26% |
| 22 | 29906934 | C | T | 4% | rs111490516 | 0.0783 | 0.0133 | 4.52E-09 | 0.039 | Novel | intronic | *MTMR3* | Yes | 13% | 0% | 0% | 13% | 2% | 0% | 2% | 5% | 0% | 0% | 1% | 3% | 0% | 1% | 0% |
| X | 31836665 | G | C | 41% | rs1379871 | 0.0287 | 0.0042 | 1.35E-11 | 0.052 | Yes | intronic | *DMD* | No | 45% | 18% | 63% | 52% | 62% | 53% | 40% | 42% | 33% | 70% | 62% | 47% | 48% | 64% | 67% |

*CHR, chromosome; POS, position; REF, reference allele; ALT, alternative allele; ALT freq, aternative allele frequency; SE, standard error; PVE, percent variance explained; OASIS, Omics Analysis, Search & Information System*

[a] Known index variant 'Yes' indicates previously published index variant from NHGRI-EBI GWAS Catalog; 'No' indicates index variant within 500 kb ± of the published lead variant, not independent of known signal in conditional analysis; 'Novel' indicates new lead variant either not published or conditionally independent.

[b] Novel locus 'Yes' was defined if there is no known index variant within 500 kb ± of the lead variant in current analysis.

**Supplementary Data 6. Genome-wide significant variants by locus from African and European population group-specific analyses**

| Population group | rsID | CHR | POS (hg38) | REF/ALT | ALT Freq | Beta | SE | *P*- value | PVE (%) | Nearest Gene | Known index variant[a] | Novel locus[b] | Index SNP in discovery analysis | Distance from index SNP | $R^2$ |
|---|---|---|---|---|---|---|---|---|---|---|---|---|---|---|---|
| African | rs543874 | 1 | 177920345 | A/G | 25.0% | 0.0731 | 0.0111 | 4.00E-11 | 0.194 | *SEC16B* | Yes | No | rs543874 (self) | 0 | - |
| | rs62033408 | 16 | 53794050 | A/G | 10.6% | 0.0982 | 0.0159 | 6.72E-10 | 0.170 | *FTO* | No | No | rs1421085 | 27008 | 0.951 |
| | rs73396827 | 22 | 29906123 | C/T | 12.6% | 0.0873 | 0.0145 | 1.66E-09 | 0.162 | *MTMR3* | No | Yes | rs111490516 | 811 | 0.997 |
| European | rs539515 | 1 | 177919890 | A/C | 18.9% | 0.0556 | 0.0088 | 2.22E-10 | 0.093 | *SEC16B* | No | No | rs543874 | 455 | 0.997 |
| | rs62107261 | 2 | 422144 | T/C | 4.8% | -0.1015 | 0.0160 | 2.08E-10 | 0.093 | *ALKAL2* | Yes | No | rs939584 | 199414 | 0.000[c] |
| | rs13130484 | 4 | 45173674 | C/T | 43.1% | 0.0439 | 0.0069 | 2.28E-10 | 0.093 | *GNPDA2* | No | No | rs12507026 | 5643 | 0.986 |
| | rs62405422 | 6 | 50829192 | T/C | 18.5% | 0.0552 | 0.0089 | 4.74E-10 | 0.089 | *TFAP2B* | No | No | rs2206277 | 1621 | 0.997 |
| | rs1459184 | 8 | 76032892 | A/T | 54.2% | 0.0437 | 0.0069 | 2.28E-10 | 0.093 | *HNF4G* | No | No | rs830463 | 35734 | 0.912 |
| | rs1421085 | 16 | 53767042 | T/C | 41.9% | 0.0917 | 0.0070 | 1.22E-39 | 0.400 | *FTO* | Yes | No | rs1421085 (self) | 0 | - |
| | rs17175643 | 18 | 60229509 | C/T | 26.3% | 0.0486 | 0.0078 | 4.11E-10 | 0.090 | *MC4R* | No | No | rs6567160 | 67607 | 0.751 |

*CHR, chromosome; POS, position; REF, reference allele; ALT, alternative allele; ALT freq, alternative allele frequency; SE, standard error; PVE, percent variance explained; SNP, single nucleotide polymorphism*

[a] Known index variant 'Yes' indicates previously published index variant from NHGRI-EBI GWAS Catalog; 'No' indicates index variant within 500 kb ± of the published lead variant, not independent of known signal in conditional analysis; 'Novel' indicates new lead variant either not published or conditionally independent.

[b] Novel locus 'Yes' was defined if there is no known index variant within 500 kb ± of the lead variant in current analysis.

[c] Linkage disequilibrium was calculated using our own data in the total population. All other $R^2$ values were calculated using TOPLD (http://topld.genetics.unc.edu/).

**Supplementary Data 7. Replication of rs111490516**

| Study | N | Imputation reference panel | Imputation $R^2$ | Freq | Beta | SE | *P*- value |
|---|---|---|---|---|---|---|---|
| MEC | 7,907 | 1000 Genomes | 0.9910 | 0.113 | 0.0354 | 0.0236 | 0.1325 |
| MVP | 79,889 | 1000 Genomes Project phase 3, version 5 | 0.7083 | 0.109 | 0.0181 | 0.0090 | 0.0442 |
| BioMe | 4,413 | TOPMed | 0.9775 | 0.125 | 0.0439 | 0.0311 | 0.1939 |
| UKBB | 8,863 | UK10K + HRC | 0.9827 | 0.129 | 0.0260 | 0.0222 | 0.2414 |
| REGARDS | 8,676 | TOPMed | 0.9981 | 0.125 | 0.0521 | 0.0226 | 0.0213 |
| **Meta-analysis** | **109,748** | | | **0.114** | **0.0253** | **0.0072** | **4.76E-04** |
| Discovery | 88,873 | | | 0.037 | 0.0783 | 0.0133 | 4.52E-09 |
| **Meta-analysis of Discovery + Replication** | **198,621** | | | **0.079** | **0.0372** | **0.0063** | **4.19E-09** |

*Freq, frequency; SE, standard error*

*MEC, Multiethnic Cohort Study; MVP, Million Veteran Program; BioMe, BioMe BioBank; UKBB, United Kingdom BioBank; REGARDS, Reasons for Geographic And Racial Differences in Stroke Study*

**Supplementary Data 8. Variant annotation from Variant Effect Predictor (VEP) for all SNPs in high LD ($R^2 > 0.8$) with top SNP in novel *MTMR3* locus.**

| Uploaded_variation | Location | Allele | Consequence | IMPACT | Entrez Gene SYMBOL | Feature_type | Feature | BIOTYPE | INTRON | HGVSc | Existing_variation | DISTANCE | STRAND | AF | SOMATIC | PHENO | CADD_PHRED | CADD_RAW | SpliceAI_pred_DS_AG | SpliceAI_pred_DS_AL | SpliceAI_pred_DS_DG | SpliceAI_pred_DS_DL | SpliceAI_pred_SYMBOL |
|---|---|---|---|---|---|---|---|---|---|---|---|---|---|---|---|---|---|---|---|---|---|---|---|
| rs111490516 | 22:29906934-29906934 | T | intron_variant | MODIFIER | *MTMR3* | Transcript | ENST00000323630.9 | protein_coding | 1/18 | - | rs111490516 | - | 1 | 0.0397 | - | - | 0.443 | -0.229 | 0.00 | 0.00 | 0.00 | 0.00 | *MTMR3* |
| | | T | intron_variant | | *MTMR3* | Transcript | ENST00000333027.7 | protein_coding | 1/19 | - | | - | 1 | 0.0397 | - | - | 0.443 | -0.229 | 0.00 | 0.00 | 0.00 | 0.00 | *MTMR3* |
| | | T | intron_variant | | *MTMR3* | Transcript | ENST00000351488.7 | protein_coding | 1/18 | - | | - | 1 | 0.0397 | - | - | 0.443 | -0.229 | 0.00 | 0.00 | 0.00 | 0.00 | *MTMR3* |
| | | T | intron_variant | | *MTMR3* | Transcript | ENST00000401950.7 | protein_coding | 1/19 | - | | - | 1 | 0.0397 | - | - | 0.443 | -0.229 | 0.00 | 0.00 | 0.00 | 0.00 | *MTMR3* |
| | | T | intron_variant,non_coding_transcript_variant | | *MTMR3* | Transcript | ENST00000415511.1 | protein_coding_CDS_not_defined | 1/5 | - | | - | 1 | 0.0397 | - | - | 0.443 | -0.229 | 0.00 | 0.00 | 0.00 | 0.00 | *MTMR3* |
| | | T | intron_variant | | *MTMR3* | Transcript | ENST00000445401.5 | protein_coding | 1/5 | - | | - | 1 | 0.0397 | - | - | 0.443 | -0.229 | 0.00 | 0.00 | 0.00 | 0.00 | *MTMR3* |
| | | T | intron_variant,non_coding_transcript_variant | | *MTMR3* | Transcript | ENST00000495098.5 | protein_coding_CDS_not_defined | 1/4 | - | | - | 1 | 0.0397 | - | - | 0.443 | -0.229 | 0.00 | 0.00 | 0.00 | 0.00 | *MTMR3* |
| rs6006286 | 22:29868401-29868401 | C | regulatory_region_variant | MODIFIER | - | RegulatoryFeature | ENSR00001059241 | enhancer | - | - | rs6006286 | - | - | 0.0421 | - | - | 1.44 | 0.0161 | - | - | - | - | - |
| | | G | regulatory_region_variant | | - | RegulatoryFeature | ENSR00001059241 | enhancer | - | - | | - | - | - | - | - | 1.648 | 0.0458 | - | - | - | - | - |
| | | C | intergenic_variant | | - | - | - | - | - | - | | - | - | 0.0421 | - | - | 1.44 | 0.0161 | - | - | - | - | - |
| | | G | intergenic_variant | | - | - | - | - | - | - | | - | - | - | - | - | 1.648 | 0.0458 | - | - | - | - | - |
| rs73394881 | 22:29871862-29871862 | A | regulatory_region_variant | MODIFIER | - | RegulatoryFeature | ENSR00001059243 | enhancer | - | - | rs73394881 | - | - | 0.0351 | - | - | **18.25** | 1.8692 | - | - | - | - | - |
| | | A | intergenic_variant | | - | - | - | - | - | - | | - | - | 0.0351 | - | - | **18.25** | 1.8692 | - | - | - | - | - |
| rs73394884 | 22:29872883-29872883 | G | regulatory_region_variant | MODIFIER | - | RegulatoryFeature | ENSR00001059243 | enhancer | - | - | rs73394884 | - | - | 0.0353 | - | - | 5.087 | 0.3697 | - | - | - | - | - |
| | | T | regulatory_region_variant | | - | RegulatoryFeature | ENSR00001059243 | enhancer | - | - | | - | - | - | - | - | 5.593 | 0.4151 | - | - | - | - | - |
| | | G | intergenic_variant | | - | - | - | - | - | - | | - | - | 0.0353 | - | - | 5.087 | 0.3697 | - | - | - | - | - |
| | | T | intergenic_variant | | - | - | - | - | - | - | | - | - | - | - | - | 5.593 | 0.4151 | - | - | - | - | - |
| rs74832232 | 22:29873682-29873682 | G | regulatory_region_variant | MODIFIER | - | RegulatoryFeature | ENSR00001059243 | enhancer | - | - | rs74832232 | - | - | 0.0353 | - | - | **17.07** | 1.7057 | - | - | - | - | - |
| | | G | intergenic_variant | | - | - | - | - | - | - | | - | - | 0.0353 | - | - | **17.07** | 1.7057 | - | - | - | - | - |
| rs73394885 | 22:29875581-29875581 | A | intergenic_variant | MODIFIER | - | - | - | - | - | - | rs73394885 | - | - | 0.0353 | - | - | 4.218 | 0.2939 | - | - | - | - | - |
| rs73394892 | 22:29880062-29880062 | A | upstream_gene_variant | MODIFIER | *MTMR3* | Transcript | NM_021090.4 | protein_coding | - | - | rs73394892 | 3112 | 1 | 0.0012 | - | - | 0.255 | -0.341 | - | - | - | - | - |
| | | C | upstream_gene_variant | | *MTMR3* | Transcript | NM_021090.4 | protein_coding | - | - | | 3112 | 1 | - | - | - | 0.214 | -0.378 | - | - | - | - | - |
| | | T | upstream_gene_variant | | *MTMR3* | Transcript | NM_021090.4 | protein_coding | - | - | | 3112 | 1 | 0.0357 | - | - | 0.202 | -0.39 | - | - | - | - | - |
| | | A | upstream_gene_variant | | *MTMR3* | Transcript | NM_153050.3 | protein_coding | - | - | | 3112 | 1 | 0.0012 | - | - | 0.255 | -0.341 | - | - | - | - | - |
| | | C | upstream_gene_variant | | *MTMR3* | Transcript | NM_153050.3 | protein_coding | - | - | | 3112 | 1 | - | - | - | 0.214 | -0.378 | - | - | - | - | - |
| | | T | upstream_gene_variant | | *MTMR3* | Transcript | NM_153050.3 | protein_coding | - | - | | 3112 | 1 | 0.0357 | - | - | 0.202 | -0.39 | - | - | - | - | - |
| | | A | upstream_gene_variant | | *MTMR3* | Transcript | NM_153051.3 | protein_coding | - | - | | 3112 | 1 | 0.0012 | - | - | 0.255 | -0.341 | - | - | - | - | - |
| | | C | upstream_gene_variant | | *MTMR3* | Transcript | NM_153051.3 | protein_coding | - | - | | 3112 | 1 | - | - | - | 0.214 | -0.378 | - | - | - | - | - |
| | | T | upstream_gene_variant | | *MTMR3* | Transcript | NM_153051.3 | protein_coding | - | - | | 3112 | 1 | 0.0357 | - | - | 0.202 | -0.39 | - | - | - | - | - |
| rs60573683 | 22:29883670-29883670 | T | intron_variant | MODIFIER | *MTMR3* | Transcript | NM_021090.4 | protein_coding | 1/19 | NM_021090.4:c.-138+311G>T | rs60573683 | - | 1 | 0.0353 | - | - | 3.147 | 0.2021 | 0.00 | 0 | 0 | 0.00 | *MTMR3* |
| | | T | intron_variant | | *MTMR3* | Transcript | NM_153050.3 | protein_coding | 1/19 | NM_153050.3:c.-138+311G>T | | - | 1 | 0.0353 | - | - | 3.147 | 0.2021 | 0.00 | 0.00 | 0.00 | 0.00 | *MTMR3* |
| | | T | intron_variant | | *MTMR3* | Transcript | NM_153051.3 | protein_coding | 1/18 | NM_153051.3:c.-138+311G>T | | - | 1 | 0.0353 | - | - | 3.147 | 0.2021 | 0.00 | 0.00 | 0.00 | 0.00 | *MTMR3* |
| | | T | regulatory_region_variant | | - | RegulatoryFeature | ENSR00000145133 | promoter | - | - | | - | - | 0.0353 | - | - | 3.147 | 0.2021 | - | - | - | - | - |
| rs73394897 | 22:29888561-29888561 | C | intron_variant | MODIFIER | *MTMR3* | Transcript | NM_021090.4 | protein_coding | 1/19 | NM_021090.4:c.-138+5202T>C | rs73394897, COSV60304704 | - | 1 | 0.0353 | 0,1 | 0,1 | **11.25** | 0.9703 | 0.00 | 0.00 | 0.00 | 0.00 | *MTMR3* |
| | | C | intron_variant | | *MTMR3* | Transcript | NM_153050.3 | protein_coding | 1/19 | NM_153050.3:c.-138+5202T>C | | - | 1 | 0.0353 | 0,1 | 0,1 | **11.25** | 0.9703 | 0.00 | 0.00 | 0.00 | 0.00 | *MTMR3* |
| | | C | intron_variant | | *MTMR3* | Transcript | NM_153051.3 | protein_coding | 1/18 | NM_153051.3:c.-138+5202T>C | | - | 1 | 0.0353 | 0,1 | 0,1 | **11.25** | 0.9703 | 0.00 | 0.00 | 0.00 | 0.00 | *MTMR3* |
| rs112565384 | 22:29890308-29890308 | A | intron_variant | MODIFIER | *MTMR3* | Transcript | NM_021090.4 | protein_coding | 1/19 | NM_021090.4:c.-138+6949G>A | rs112565384 | - | 1 | 0.0353 | - | - | 1.965 | 0.0856 | 0.00 | 0.00 | 0.00 | 0.00 | *MTMR3* |
| | | A | intron_variant | | *MTMR3* | Transcript | NM_153050.3 | protein_coding | 1/19 | NM_153050.3:c.-138+6949G>A | | - | 1 | 0.0353 | - | - | 1.965 | 0.0856 | 0.00 | 0.00 | 0.00 | 0.00 | *MTMR3* |
| | | A | intron_variant | | *MTMR3* | Transcript | NM_153051.3 | protein_coding | 1/18 | NM_153051.3:c.-138+6949G>A | | - | 1 | 0.0353 | - | - | 1.965 | 0.0856 | 0.00 | 0.00 | 0.00 | 0.00 | *MTMR3* |
| rs73396810 | 22:29895377-29895377 | T | intron_variant | MODIFIER | *MTMR3* | Transcript | NM_021090.4 | protein_coding | 1/19 | NM_021090.4:c.-138+12018G>T | rs73396810 | - | 1 | 0.0353 | - | - | 1.479 | 0.022 | 0.00 | 0.00 | 0.00 | 0.00 | *MTMR3* |
| | | T | intron_variant | | *MTMR3* | Transcript | NM_153050.3 | protein_coding | 1/19 | NM_153050.3:c.-138+12018G>T | | - | 1 | 0.0353 | - | - | 1.479 | 0.022 | 0.00 | 0.00 | 0.00 | 0.00 | *MTMR3* |
| | | T | intron_variant | | *MTMR3* | Transcript | NM_153051.3 | protein_coding | 1/18 | NM_153051.3:c.-138+12018G>T | | - | 1 | 0.0353 | - | - | 1.479 | 0.022 | 0.00 | 0.00 | 0.00 | 0.00 | *MTMR3* |
| rs73396811 | 22:29896358-29896358 | G | intron_variant | MODIFIER | *MTMR3* | Transcript | NM_021090.4 | protein_coding | 1/19 | NM_021090.4:c.-138+12999A>G | rs73396811 | - | 1 | 0.0353 | - | - | 5.789 | 0.4329 | 0.00 | 0.00 | 0.00 | 0.00 | *MTMR3* |
| | | G | intron_variant | | *MTMR3* | Transcript | NM_153050.3 | protein_coding | 1/19 | NM_153050.3:c.-138+12999A>G | | - | 1 | 0.0353 | - | - | 5.789 | 0.4329 | 0.00 | 0.00 | 0.00 | 0.00 | *MTMR3* |
| | | G | intron_variant | | *MTMR3* | Transcript | NM_153051.3 | protein_coding | 1/18 | NM_153051.3:c.-138+12999A>G | | - | 1 | 0.0353 | - | - | 5.789 | 0.4329 | 0.00 | 0.00 | 0.00 | 0.00 | *MTMR3* |
| rs73396818 | 22:29902155-29902155 | T | intron_variant | MODIFIER | *MTMR3* | Transcript | NM_021090.4 | protein_coding | 1/19 | NM_021090.4:c.-138+18796A>T | rs73396818 | - | 1 | 0.0397 | - | - | 0.366 | -0.267 | 0.00 | 0.00 | 0.00 | 0.00 | *MTMR3* |
| | | T | intron_variant | | *MTMR3* | Transcript | NM_153050.3 | protein_coding | 1/19 | NM_153050.3:c.-138+18796A>T | | - | 1 | 0.0397 | - | - | 0.366 | -0.267 | 0.00 | 0.00 | 0.00 | 0.00 | *MTMR3* |
| | | T | intron_variant | | *MTMR3* | Transcript | NM_153051.3 | protein_coding | 1/18 | NM_153051.3:c.-138+18796A>T | | - | 1 | 0.0397 | - | - | 0.366 | -0.267 | 0.00 | 0.00 | 0.00 | 0.00 | *MTMR3* |
| rs113463187 | 22:29902988-29902988 | T | intron_variant | MODIFIER | *MTMR3* | Transcript | NM_021090.4 | protein_coding | 1/19 | NM_021090.4:c.-138+19629C>T | rs113463187 | - | 1 | 0.0399 | - | - | 2.775 | 0.1688 | 0.00 | 0.00 | 0.00 | 0.00 | *MTMR3* |
| | | T | intron_variant | | *MTMR3* | Transcript | NM_153050.3 | protein_coding | 1/19 | NM_153050.3:c.-138+19629C>T | | - | 1 | 0.0399 | - | - | 2.775 | 0.1688 | 0.00 | 0.00 | 0.00 | 0.00 | *MTMR3* |
| | | T | intron_variant | | *MTMR3* | Transcript | NM_153051.3 | protein_coding | 1/18 | NM_153051.3:c.-138+19629C>T | | - | 1 | 0.0399 | - | - | 2.775 | 0.1688 | 0.00 | 0.00 | 0.00 | 0.00 | *MTMR3* |
| rs57349783 | 22:29903544-29903544 | C | intron_variant | MODIFIER | *MTMR3* | Transcript | NM_021090.4 | protein_coding | 1/19 | NM_021090.4:c.-138+20185T>C | rs57349783 | - | 1 | 0.0395 | - | - | 1.297 | -0.006 | 0.00 | 0.00 | 0.00 | 0.00 | *MTMR3* |
| | | C | intron_variant | | *MTMR3* | Transcript | NM_153050.3 | protein_coding | 1/19 | NM_153050.3:c.-138+20185T>C | | - | 1 | 0.0395 | - | - | 1.297 | -0.006 | 0.00 | 0.00 | 0.00 | 0.00 | *MTMR3* |
| | | C | intron_variant | | *MTMR3* | Transcript | NM_153051.3 | protein_coding | 1/18 | NM_153051.3:c.-138+20185T>C | | - | 1 | 0.0395 | - | - | 1.297 | -0.006 | 0.00 | 0.00 | 0.00 | 0.00 | *MTMR3* |
| rs2158535 | 22:29903616-29903616 | C | intron_variant | MODIFIER | *MTMR3* | Transcript | NM_021090.4 | protein_coding | 1/19 | NM_021090.4:c.-138+20257T>C | rs2158535 | - | 1 | 0.0395 | - | - | 1.888 | 0.0764 | 0.00 | 0.00 | 0.00 | 0.00 | *MTMR3* |
| | | C | intron_variant | | *MTMR3* | Transcript | NM_153050.3 | protein_coding | 1/19 | NM_153050.3:c.-138+20257T>C | | - | 1 | 0.0395 | - | - | 1.888 | 0.0764 | 0.00 | 0.00 | 0.00 | 0.00 | *MTMR3* |
| | | C | intron_variant | | *MTMR3* | Transcript | NM_153051.3 | protein_coding | 1/18 | NM_153051.3:c.-138+20257T>C | | - | 1 | 0.0395 | - | - | 1.888 | 0.0764 | 0.00 | 0.00 | 0.00 | 0.00 | *MTMR3* |
| rs2107672 | 22:29903764-29903764 | A | intron_variant | MODIFIER | *MTMR3* | Transcript | NM_021090.4 | protein_coding | 1/19 | NM_021090.4:c.-138+20405G>A | rs2107672 | - | 1 | 0.0353 | - | - | 8.09 | 0.6607 | 0.00 | 0.00 | 0.00 | 0.00 | *MTMR3* |
| | | A | intron_variant | | *MTMR3* | Transcript | NM_153050.3 | protein_coding | 1/19 | NM_153050.3:c.-138+20405G>A | | - | 1 | 0.0353 | - | - | 8.09 | 0.6607 | 0.00 | 0.00 | 0.00 | 0.00 | *MTMR3* |
| | | A | intron_variant | | *MTMR3* | Transcript | NM_153051.3 | protein_coding | 1/18 | NM_153051.3:c.-138+20405G>A | | - | 1 | 0.0353 | - | - | 8.09 | 0.6607 | 0.00 | 0.00 | 0.00 | 0.00 | *MTMR3* |
| rs2107673 | 22:29903808-29903808 | A | intron_variant | MODIFIER | *MTMR3* | Transcript | NM_021090.4 | protein_coding | 1/19 | NM_021090.4:c.-138+20449T>A | rs2107673 | - | 1 | - | - | - | 0.204 | -0.387 | 0.00 | 0.00 | 0.00 | 0.00 | *MTMR3* |
| | | C | intron_variant | | *MTMR3* | Transcript | NM_021090.4 | protein_coding | 1/19 | NM_021090.4:c.-138+20449T>C | | - | 1 | 0.0395 | - | - | 0.258 | -0.339 | 0.00 | 0.00 | 0.00 | 0.00 | *MTMR3* |
| | | A | intron_variant | | *MTMR3* | Transcript | NM_153050.3 | protein_coding | 1/19 | NM_153050.3:c.-138+20449T>A | | - | 1 | - | - | - | 0.204 | -0.387 | 0.00 | 0.00 | 0.00 | 0.00 | *MTMR3* |
| | | C | intron_variant | | *MTMR3* | Transcript | NM_153050.3 | protein_coding | 1/19 | NM_153050.3:c.-138+20449T>C | | - | 1 | 0.0395 | - | - | 0.258 | -0.339 | 0.00 | 0.00 | 0.00 | 0.00 | *MTMR3* |
| | | A | intron_variant | | *MTMR3* | Transcript | NM_153051.3 | protein_coding | 1/18 | NM_153051.3:c.-138+20449T>A | | - | 1 | - | - | - | 0.204 | -0.387 | 0.00 | 0.00 | 0.00 | 0.00 | *MTMR3* |
| | | C | intron_variant | | *MTMR3* | Transcript | NM_153051.3 | protein_coding | 1/18 | NM_153051.3:c.-138+20449T>C | | - | 1 | 0.0395 | - | - | 0.258 | -0.339 | 0.00 | 0.00 | 0.00 | 0.00 | *MTMR3* |
| rs73396822 | 22:29904208-29904208 | A | intron_variant | MODIFIER | *MTMR3* | Transcript | NM_021090.4 | protein_coding | 1/19 | NM_021090.4:c.-138+20849C>A | rs73396822 | - | 1 | - | - | - | 5.187 | 0.3785 | 0.00 | 0.00 | 0.00 | 0.00 | *MTMR3* |
| | | G | intron_variant | | *MTMR3* | Transcript | NM_021090.4 | protein_coding | 1/19 | NM_021090.4:c.-138+20849C>G | | - | 1 | 0.0395 | - | - | 5.379 | 0.3957 | 0.00 | 0.00 | 0.00 | 0.00 | *MTMR3* |
| | | A | intron_variant | | *MTMR3* | Transcript | NM_153050.3 | protein_coding | 1/19 | NM_153050.3:c.-138+20849C>A | | - | 1 | - | - | - | 5.187 | 0.3785 | 0.00 | 0.00 | 0.00 | 0.00 | *MTMR3* |
| | | G | intron_variant | | *MTMR3* | Transcript | NM_153050.3 | protein_coding | 1/19 | NM_153050.3:c.-138+20849C>G | | - | 1 | 0.0395 | - | - | 5.379 | 0.3957 | 0.00 | 0.00 | 0.00 | 0.00 | *MTMR3* |
| | | A | intron_variant | | *MTMR3* | Transcript | NM_153051.3 | protein_coding | 1/18 | NM_153051.3:c.-138+20849C>A | | - | 1 | - | - | - | 5.187 | 0.3785 | 0.00 | 0.00 | 0.00 | 0.00 | *MTMR3* |
| | | G | intron_variant | | *MTMR3* | Transcript | NM_153051.3 | protein_coding | 1/18 | NM_153051.3:c.-138+20849C>G | | - | 1 | 0.0395 | - | - | 5.379 | 0.3957 | 0.00 | 0.00 | 0.00 | 0.00 | *MTMR3* |
| rs73396823 | 22:29904279-29904279 | A | intron_variant | MODIFIER | *MTMR3* | Transcript | NM_021090.4 | protein_coding | 1/19 | NM_021090.4:c.-138+20920G>A | rs73396823 | - | 1 | 0.0395 | - | - | 0.683 | -0.14 | 0.00 | 0.00 | 0.00 | 0.00 | *MTMR3* |
| | | C | intron_variant | | *MTMR3* | Transcript | NM_021090.4 | protein_coding | 1/19 | NM_021090.4:c.-138+20920G>C | | - | 1 | - | - | - | 0.572 | -0.177 | 0.00 | 0.00 | 0.00 | 0.00 | *MTMR3* |
| | | A | intron_variant | | *MTMR3* | Transcript | NM_153050.3 | protein_coding | 1/19 | NM_153050.3:c.-138+20920G>A | | - | 1 | 0.0395 | - | - | 0.683 | -0.14 | 0.00 | 0.00 | 0.00 | 0.00 | *MTMR3* |
| | | C | intron_variant | | *MTMR3* | Transcript | NM_153050.3 | protein_coding | 1/19 | NM_153050.3:c.-138+20920G>C | | - | 1 | - | - | - | 0.572 | -0.177 | 0.00 | 0.00 | 0.00 | 0.00 | *MTMR3* |
| | | A | intron_variant | | *MTMR3* | Transcript | NM_153051.3 | protein_coding | 1/18 | NM_153051.3:c.-138+20920G>A | | - | 1 | 0.0395 | - | - | 0.683 | -0.14 | 0.00 | 0.00 | 0.00 | 0.00 | *MTMR3* |
| | | C | intron_variant | | *MTMR3* | Transcript | NM_153051.3 | protein_coding | 1/18 | NM_153051.3:c.-138+20920G>C | | - | 1 | - | - | - | 0.572 | -0.177 | 0.00 | 0.00 | 0.00 | 0.00 | *MTMR3* |
| rs2051858 | 22:29904422-29904422 | T | intron_variant | MODIFIER | *MTMR3* | Transcript | NM_021090.4 | protein_coding | 1/19 | NM_021090.4:c.-138+21063C>T | rs2051858, COSV60307792 | - | 1 | 0.0395 | 0,1 | 0,1 | 1.027 | -0.055 | 0.00 | 0.00 | 0.00 | 0.00 | *MTMR3* |
| | | T | intron_variant | | *MTMR3* | Transcript | NM_153050.3 | protein_coding | 1/19 | NM_153050.3:c.-138+21063C>T | | - | 1 | 0.0395 | 0,1 | 0,1 | 1.027 | -0.055 | 0.00 | 0.00 | 0.00 | 0.00 | *MTMR3* |
| | | T | intron_variant | | *MTMR3* | Transcript | NM_153051.3 | protein_coding | 1/18 | NM_153051.3:c.-138+21063C>T | | - | 1 | 0.0395 | 0,1 | 0,1 | 1.027 | -0.055 | 0.00 | 0.00 | 0.00 | 0.00 | *MTMR3* |
| rs2051859 | 22:29904532-29904532 | A | intron_variant | MODIFIER | *MTMR3* | Transcript | NM_021090.4 | protein_coding | 1/19 | NM_021090.4:c.-138+21173G>A | rs2051859 | - | 1 | 0.0395 | - | - | 4.256 | 0.2972 | 0.00 | 0.00 | 0.00 | 0.00 | *MTMR3* |
| | | A | intron_variant | | *MTMR3* | Transcript | NM_153050.3 | protein_coding | 1/19 | NM_153050.3:c.-138+21173G>A | | - | 1 | 0.0395 | - | - | 4.256 | 0.2972 | 0.00 | 0.00 | 0.00 | 0.00 | *MTMR3* |
| | | A | intron_variant | | *MTMR3* | Transcript | NM_153051.3 | protein_coding | 1/18 | NM_153051.3:c.-138+21173G>A | | - | 1 | 0.0395 | - | - | 4.256 | 0.2972 | 0.00 | 0.00 | 0.00 | 0.00 | *MTMR3* |

| | | | | | | | | | | | | | | | | | | | | | | | |
|---|---|---|---|---|---|---|---|---|---|---|---|---|---|---|---|---|---|---|---|---|---|---|---|
| rs73396826 | 22:29906042-29906042 | A | intron_variant | MODIFIER | *MTMR3* | Transcript | NM_021090.4 | protein_coding | 1/19 | NM_021090.4:c.-138+22683G>A | rs73396826 | - | 1 | 0.0395 | - | - | 0.281 | -0.321 | 0.00 | 0.00 | 0.00 | 0.00 | *MTMR3* |
| | | A | intron_variant | | *MTMR3* | Transcript | NM_153050.3 | protein_coding | 1/19 | NM_153050.3:c.-138+22683G>A | | - | 1 | 0.0395 | - | - | 0.281 | -0.321 | 0.00 | 0.00 | 0.00 | 0.00 | *MTMR3* |
| | | A | intron_variant | | *MTMR3* | Transcript | NM_153051.3 | protein_coding | 1/18 | NM_153051.3:c.-138+22683G>A | | - | 1 | 0.0395 | - | - | 0.281 | -0.321 | 0.00 | 0.00 | 0.00 | 0.00 | *MTMR3* |
| rs73396827 | 22:29906123-29906123 | T | intron_variant | MODIFIER | *MTMR3* | Transcript | NM_021090.4 | protein_coding | 1/19 | NM_021090.4:c.-138+22764C>T | rs73396827 | - | 1 | 0.0395 | - | - | 0.952 | -0.071 | 0.00 | 0.00 | 0.04 | 0.00 | *MTMR3* |
| | | T | intron_variant | | *MTMR3* | Transcript | NM_153050.3 | protein_coding | 1/19 | NM_153050.3:c.-138+22764C>T | | - | 1 | 0.0395 | - | - | 0.952 | -0.071 | 0.00 | 0.00 | 0.04 | 0.00 | *MTMR3* |
| | | T | intron_variant | | *MTMR3* | Transcript | NM_153051.3 | protein_coding | 1/18 | NM_153051.3:c.-138+22764C>T | | - | 1 | 0.0395 | - | - | 0.952 | -0.071 | 0.00 | 0.00 | 0.04 | 0.00 | *MTMR3* |
| rs73396828 | 22:29906206-29906206 | C | intron_variant | MODIFIER | *MTMR3* | Transcript | NM_021090.4 | protein_coding | 1/19 | NM_021090.4:c.-138+22847T>C | rs73396828 | - | 1 | 0.0389 | - | - | 1.86 | 0.073 | 0.00 | 0.00 | 0.00 | 0.00 | *MTMR3* |
| | | C | intron_variant | | *MTMR3* | Transcript | NM_153050.3 | protein_coding | 1/19 | NM_153050.3:c.-138+22847T>C | | - | 1 | 0.0389 | - | - | 1.86 | 0.073 | 0.00 | 0.00 | 0.00 | 0.00 | *MTMR3* |
| | | C | intron_variant | | *MTMR3* | Transcript | NM_153051.3 | protein_coding | 1/18 | NM_153051.3:c.-138+22847T>C | | - | 1 | 0.0389 | - | - | 1.86 | 0.073 | 0.00 | 0.00 | 0.00 | 0.00 | *MTMR3* |
| rs73396830 | 22:29906213-29906213 | G | intron_variant | MODIFIER | *MTMR3* | Transcript | NM_021090.4 | protein_coding | 1/19 | NM_021090.4:c.-138+22854A>G | rs73396830 | - | 1 | 0.0389 | - | - | 0.226 | -0.367 | 0.00 | 0.00 | 0.00 | 0.00 | *MTMR3* |
| | | G | intron_variant | | *MTMR3* | Transcript | NM_153050.3 | protein_coding | 1/19 | NM_153050.3:c.-138+22854A>G | | - | 1 | 0.0389 | - | - | 0.226 | -0.367 | 0.00 | 0.00 | 0.00 | 0.00 | *MTMR3* |
| | | G | intron_variant | | *MTMR3* | Transcript | NM_153051.3 | protein_coding | 1/18 | NM_153051.3:c.-138+22854A>G | | - | 1 | 0.0389 | - | - | 0.226 | -0.367 | 0.00 | 0.00 | 0.00 | 0.00 | *MTMR3* |
| rs73396831 | 22:29906421-29906421 | T | intron_variant | MODIFIER | *MTMR3* | Transcript | NM_021090.4 | protein_coding | 1/19 | NM_021090.4:c.-138+23062C>T | rs73396831 | - | 1 | 0.0395 | - | - | 1.009 | -0.059 | 0.00 | 0.00 | 0.00 | 0.00 | *MTMR3* |
| | | T | intron_variant | | *MTMR3* | Transcript | NM_153050.3 | protein_coding | 1/19 | NM_153050.3:c.-138+23062C>T | | - | 1 | 0.0395 | - | - | 1.009 | -0.059 | 0.00 | 0.00 | 0.00 | 0.00 | *MTMR3* |
| | | T | intron_variant | | *MTMR3* | Transcript | NM_153051.3 | protein_coding | 1/18 | NM_153051.3:c.-138+23062C>T | | - | 1 | 0.0395 | - | - | 1.009 | -0.059 | 0.00 | 0.00 | 0.00 | 0.00 | *MTMR3* |
| rs112056808 | 22:29907171-29907171 | T | intron_variant | MODIFIER | *MTMR3* | Transcript | NM_021090.4 | protein_coding | 1/19 | NM_021090.4:c.-138+23812C>T | rs112056808 | - | 1 | 0.0395 | - | - | 1.207 | -0.021 | 0.00 | 0.00 | 0.00 | 0.00 | *MTMR3* |
| | | T | intron_variant | | *MTMR3* | Transcript | NM_153050.3 | protein_coding | 1/19 | NM_153050.3:c.-138+23812C>T | | - | 1 | 0.0395 | - | - | 1.207 | -0.021 | 0.00 | 0.00 | 0.00 | 0.00 | *MTMR3* |
| | | T | intron_variant | | *MTMR3* | Transcript | NM_153051.3 | protein_coding | 1/18 | NM_153051.3:c.-138+23812C>T | | - | 1 | 0.0395 | - | - | 1.207 | -0.021 | 0.00 | 0.00 | 0.00 | 0.00 | *MTMR3* |
| rs550468632 | 22:29908987-29908987 | C | intron_variant | MODIFIER | *MTMR3* | Transcript | NM_021090.4 | protein_coding | 1/19 | NM_021090.4:c.-138+25628T>C | rs550468632 | - | 1 | 0.0375 | - | - | 0.195 | -0.397 | 0.00 | 0.00 | 0.00 | 0.00 | *MTMR3* |
| | | C | intron_variant | | *MTMR3* | Transcript | NM_153050.3 | protein_coding | 1/19 | NM_153050.3:c.-138+25628T>C | | - | 1 | 0.0375 | - | - | 0.195 | -0.397 | 0.00 | 0.00 | 0.00 | 0.00 | *MTMR3* |
| | | C | intron_variant | | *MTMR3* | Transcript | NM_153051.3 | protein_coding | 1/18 | NM_153051.3:c.-138+25628T>C | | - | 1 | 0.0375 | - | - | 0.195 | -0.397 | 0.00 | 0.00 | 0.00 | 0.00 | *MTMR3* |
| rs73396841 | 22:29909799-29909799 | C | intron_variant | MODIFIER | *MTMR3* | Transcript | NM_021090.4 | protein_coding | 1/19 | NM_021090.4:c.-138+26440T>C | rs73396841 | - | 1 | 0.0395 | - | - | 0.02 | -0.837 | 0.00 | 0.00 | 0.00 | 0.00 | *MTMR3* |
| | | C | intron_variant | | *MTMR3* | Transcript | NM_153050.3 | protein_coding | 1/19 | NM_153050.3:c.-138+26440T>C | | - | 1 | 0.0395 | - | - | 0.02 | -0.837 | 0.00 | 0.00 | 0.00 | 0.00 | *MTMR3* |
| | | C | intron_variant | | *MTMR3* | Transcript | NM_153051.3 | protein_coding | 1/18 | NM_153051.3:c.-138+26440T>C | | - | 1 | 0.0395 | - | - | 0.02 | -0.837 | 0.00 | 0.00 | 0.00 | 0.00 | *MTMR3* |
| rs73396843 | 22:29910808-29910808 | G | intron_variant | MODIFIER | *MTMR3* | Transcript | NM_021090.4 | protein_coding | 1/19 | NM_021090.4:c.-138+27449T>G | rs73396843 | - | 1 | 0.0353 | - | - | 1.936 | 0.0822 | 0.00 | 0.00 | 0.00 | 0.00 | *MTMR3* |
| | | G | intron_variant | | *MTMR3* | Transcript | NM_153050.3 | protein_coding | 1/19 | NM_153050.3:c.-138+27449T>G | | - | 1 | 0.0353 | - | - | 1.936 | 0.0822 | 0.00 | 0.00 | 0.00 | 0.00 | *MTMR3* |
| | | G | intron_variant | | *MTMR3* | Transcript | NM_153051.3 | protein_coding | 1/18 | NM_153051.3:c.-138+27449T>G | | - | 1 | 0.0353 | - | - | 1.936 | 0.0822 | 0.00 | 0.00 | 0.00 | 0.00 | *MTMR3* |
| rs7291683 | 22:29911218-29911218 | T | intron_variant | MODIFIER | *MTMR3* | Transcript | NM_021090.4 | protein_coding | 1/19 | NM_021090.4:c.-138+27859C>T | rs7291683 | - | 1 | 0.0395 | - | - | 3.236 | 0.2098 | 0.00 | 0.00 | 0.00 | 0.00 | *MTMR3* |
| | | T | intron_variant | | *MTMR3* | Transcript | NM_153050.3 | protein_coding | 1/19 | NM_153050.3:c.-138+27859C>T | | - | 1 | 0.0395 | - | - | 3.236 | 0.2098 | 0.00 | 0.00 | 0.00 | 0.00 | *MTMR3* |
| | | T | intron_variant | | *MTMR3* | Transcript | NM_153051.3 | protein_coding | 1/18 | NM_153051.3:c.-138+27859C>T | | - | 1 | 0.0395 | - | - | 3.236 | 0.2098 | 0.00 | 0.00 | 0.00 | 0.00 | *MTMR3* |
| rs73396856 | 22:29911621-29911621 | A | intron_variant | MODIFIER | *MTMR3* | Transcript | NM_021090.4 | protein_coding | 1/19 | NM_021090.4:c.-138+28262C>A | rs73396856 | - | 1 | - | - | - | 1.996 | 0.0892 | 0.00 | 0.00 | 0.00 | 0.00 | *MTMR3* |
| | | G | intron_variant | | *MTMR3* | Transcript | NM_021090.4 | protein_coding | 1/19 | NM_021090.4:c.-138+28262C>G | | - | 1 | 0.0353 | - | - | 2.149 | 0.1065 | 0.00 | 0.00 | 0.00 | 0.00 | *MTMR3* |
| | | A | intron_variant | | *MTMR3* | Transcript | NM_153050.3 | protein_coding | 1/19 | NM_153050.3:c.-138+28262C>A | | - | 1 | - | - | - | 1.996 | 0.0892 | 0.00 | 0.00 | 0.00 | 0.00 | *MTMR3* |
| | | G | intron_variant | | *MTMR3* | Transcript | NM_153050.3 | protein_coding | 1/19 | NM_153050.3:c.-138+28262C>G | | - | 1 | 0.0353 | - | - | 2.149 | 0.1065 | 0.00 | 0.00 | 0.00 | 0.00 | *MTMR3* |
| | | A | intron_variant | | *MTMR3* | Transcript | NM_153051.3 | protein_coding | 1/18 | NM_153051.3:c.-138+28262C>A | | - | 1 | - | - | - | 1.996 | 0.0892 | 0.00 | 0.00 | 0.00 | 0.00 | *MTMR3* |
| | | G | intron_variant | | *MTMR3* | Transcript | NM_153051.3 | protein_coding | 1/18 | NM_153051.3:c.-138+28262C>G | | - | 1 | 0.0353 | - | - | 2.149 | 0.1065 | 0.00 | 0.00 | 0.00 | 0.00 | *MTMR3* |
| rs73396863 | 22:29913097-29913097 | A | intron_variant | MODIFIER | *MTMR3* | Transcript | NM_021090.4 | protein_coding | 1/19 | NM_021090.4:c.-138+29738T>A | rs73396863 | - | 1 | 0.0393 | - | - | 1.817 | 0.0676 | 0.00 | 0.00 | 0.00 | 0.00 | *MTMR3* |
| | | A | intron_variant | | *MTMR3* | Transcript | NM_153050.3 | protein_coding | 1/19 | NM_153050.3:c.-138+29738T>A | | - | 1 | 0.0393 | - | - | 1.817 | 0.0676 | 0.00 | 0.00 | 0.00 | 0.00 | *MTMR3* |
| | | A | intron_variant | | *MTMR3* | Transcript | NM_153051.3 | protein_coding | 1/18 | NM_153051.3:c.-138+29738T>A | | - | 1 | 0.0393 | - | - | 1.817 | 0.0676 | 0.00 | 0.00 | 0.00 | 0.00 | *MTMR3* |
| rs73396885 | 22:29924436-29924436 | A | intron_variant | MODIFIER | *MTMR3* | Transcript | NM_021090.4 | protein_coding | 1/19 | NM_021090.4:c.-137-32600C>A | rs73396885 | - | 1 | 0.0353 | - | - | 1.188 | -0.025 | 0.00 | 0.00 | 0.00 | 0.00 | *MTMR3* |
| | | A | intron_variant | | *MTMR3* | Transcript | NM_153050.3 | protein_coding | 1/19 | NM_153050.3:c.-137-32600C>A | | - | 1 | 0.0353 | - | - | 1.188 | -0.025 | 0.00 | 0.00 | 0.00 | 0.00 | *MTMR3* |
| | | A | intron_variant | | *MTMR3* | Transcript | NM_153051.3 | protein_coding | 1/18 | NM_153051.3:c.-137-32600C>A | | - | 1 | 0.0353 | - | - | 1.188 | -0.025 | 0.00 | 0.00 | 0.00 | 0.00 | *MTMR3* |
| | | A | regulatory_region_variant | | - | RegulatoryFeature | ENSR00001059246 | enhancer | - | - | | - | - | 0.0353 | - | - | 1.188 | -0.025 | - | - | - | - | - |
| rs60098050 | 22:29926719-29926719 | G | intron_variant | MODIFIER | *MTMR3* | Transcript | NM_021090.4 | protein_coding | 1/19 | NM_021090.4:c.-137-30317A>G | rs60098050 | - | 1 | 0.0353 | - | - | 6.904 | 0.5387 | 0.00 | 0.00 | 0.00 | 0.00 | *MTMR3* |
| | | G | intron_variant | | *MTMR3* | Transcript | NM_153050.3 | protein_coding | 1/19 | NM_153050.3:c.-137-30317A>G | | - | 1 | 0.0353 | - | - | 6.904 | 0.5387 | 0.00 | 0.00 | 0.00 | 0.00 | *MTMR3* |
| | | G | intron_variant | | *MTMR3* | Transcript | NM_153051.3 | protein_coding | 1/18 | NM_153051.3:c.-137-30317A>G | | - | 1 | 0.0353 | - | - | 6.904 | 0.5387 | 0.00 | 0.00 | 0.00 | 0.00 | *MTMR3* |
| rs111317522 | 22:29928460-29928460 | T | intron_variant | MODIFIER | *MTMR3* | Transcript | NM_021090.4 | protein_coding | 1/19 | NM_021090.4:c.-137-28576C>T | rs111317522 | - | 1 | 0.0353 | - | - | 1.598 | 0.0389 | 0.00 | 0.00 | 0.00 | 0.00 | *MTMR3* |
| | | T | intron_variant | | *MTMR3* | Transcript | NM_153050.3 | protein_coding | 1/19 | NM_153050.3:c.-137-28576C>T | | - | 1 | 0.0353 | - | - | 1.598 | 0.0389 | 0.00 | 0.00 | 0.00 | 0.00 | *MTMR3* |
| | | T | intron_variant | | *MTMR3* | Transcript | NM_153051.3 | protein_coding | 1/18 | NM_153051.3:c.-137-28576C>T | | - | 1 | 0.0353 | - | - | 1.598 | 0.0389 | 0.00 | 0.00 | 0.00 | 0.00 | *MTMR3* |
| | | T | regulatory_region_variant | | - | RegulatoryFeature | ENSR00001059247 | enhancer | - | - | | - | - | 0.0353 | - | - | 1.598 | 0.0389 | - | - | - | - | - |
| rs73396896 | 22:29931253-29931253 | A | intron_variant | MODIFIER | *MTMR3* | Transcript | NM_021090.4 | protein_coding | 1/19 | NM_021090.4:c.-137-25783C>A | rs73396896 | - | 1 | 0.0353 | - | - | **13.9** | 1.2343 | 0.00 | 0.00 | 0.00 | 0.00 | *MTMR3* |
| | | T | intron_variant | | *MTMR3* | Transcript | NM_021090.4 | protein_coding | 1/19 | NM_021090.4:c.-137-25783C>T | | - | 1 | - | - | - | **14.34** | 1.2938 | 0.00 | 0.00 | 0.00 | 0.00 | *MTMR3* |
| | | A | intron_variant | | *MTMR3* | Transcript | NM_153050.3 | protein_coding | 1/19 | NM_153050.3:c.-137-25783C>A | | - | 1 | 0.0353 | - | - | **13.9** | 1.2343 | 0.00 | 0.00 | 0.00 | 0.00 | *MTMR3* |
| | | T | intron_variant | | *MTMR3* | Transcript | NM_153050.3 | protein_coding | 1/19 | NM_153050.3:c.-137-25783C>T | | - | 1 | - | - | - | **14.34** | 1.2938 | 0.00 | 0.00 | 0.00 | 0.00 | *MTMR3* |
| | | A | intron_variant | | *MTMR3* | Transcript | NM_153051.3 | protein_coding | 1/18 | NM_153051.3:c.-137-25783C>A | | - | 1 | 0.0353 | - | - | **13.9** | 1.2343 | 0.00 | 0.00 | 0.00 | 0.00 | *MTMR3* |
| | | T | intron_variant | | *MTMR3* | Transcript | NM_153051.3 | protein_coding | 1/18 | NM_153051.3:c.-137-25783C>T | | - | 1 | - | - | - | **14.34** | 1.2938 | 0.00 | 0.00 | 0.00 | 0.00 | *MTMR3* |
| | | A | regulatory_region_variant | | - | RegulatoryFeature | ENSR00001059247 | enhancer | - | - | | - | - | 0.0353 | - | - | **13.9** | 1.2343 | - | - | - | - | - |
| | | T | regulatory_region_variant | | - | RegulatoryFeature | ENSR00001059247 | enhancer | - | - | | - | - | - | - | - | **14.34** | 1.2938 | - | - | - | - | - |
| rs113048784 | 22:29934106-29934106 | A | intron_variant | MODIFIER | *MTMR3* | Transcript | NM_021090.4 | protein_coding | 1/19 | NM_021090.4:c.-137-22930G>A | rs113048784 | - | 1 | 0.0353 | - | - | 1.133 | -0.035 | 0.00 | 0.00 | 0.00 | 0.00 | *MTMR3* |
| | | A | intron_variant | | *MTMR3* | Transcript | NM_153050.3 | protein_coding | 1/19 | NM_153050.3:c.-137-22930G>A | | - | 1 | 0.0353 | - | - | 1.133 | -0.035 | 0.00 | 0.00 | 0.00 | 0.00 | *MTMR3* |
| | | A | intron_variant | | *MTMR3* | Transcript | NM_153051.3 | protein_coding | 1/18 | NM_153051.3:c.-137-22930G>A | | - | 1 | 0.0353 | - | - | 1.133 | -0.035 | 0.00 | 0.00 | 0.00 | 0.00 | *MTMR3* |
| rs73396902 | 22:29935150-29935150 | G | intron_variant | MODIFIER | *MTMR3* | Transcript | NM_021090.4 | protein_coding | 1/19 | NM_021090.4:c.-137-21886A>G | rs73396902 | - | 1 | 0.0355 | - | - | 8.524 | 0.7075 | 0.00 | 0.00 | 0.00 | 0.00 | *MTMR3* |
| | | G | intron_variant | | *MTMR3* | Transcript | NM_153050.3 | protein_coding | 1/19 | NM_153050.3:c.-137-21886A>G | | - | 1 | 0.0355 | - | - | 8.524 | 0.7075 | 0.00 | 0.00 | 0.00 | 0.00 | *MTMR3* |
| | | G | intron_variant | | *MTMR3* | Transcript | NM_153051.3 | protein_coding | 1/18 | NM_153051.3:c.-137-21886A>G | | - | 1 | 0.0355 | - | - | 8.524 | 0.7075 | 0.00 | 0.00 | 0.00 | 0.00 | *MTMR3* |
| rs60751132 | 22:29935766-29935766 | A | intron_variant | MODIFIER | *MTMR3* | Transcript | NM_021090.4 | protein_coding | 1/19 | NM_021090.4:c.-137-21270G>A | rs60751132, COSV60308265 | - | 1 | 0.0353 | 0,1 | 0,1 | 0.785 | -0.112 | 0.00 | 0.00 | 0.00 | 0.00 | *MTMR3* |
| | | A | intron_variant | | *MTMR3* | Transcript | NM_153050.3 | protein_coding | 1/19 | NM_153050.3:c.-137-21270G>A | | - | 1 | 0.0353 | 0,1 | 0,1 | 0.785 | -0.112 | 0.00 | 0.00 | 0.00 | 0.00 | *MTMR3* |
| | | A | intron_variant | | *MTMR3* | Transcript | NM_153051.3 | protein_coding | 1/18 | NM_153051.3:c.-137-21270G>A | | - | 1 | 0.0353 | 0,1 | 0,1 | 0.785 | -0.112 | 0.00 | 0.00 | 0.00 | 0.00 | *MTMR3* |
| | | A | regulatory_region_variant | | - | RegulatoryFeature | ENSR00000669734 | enhancer | - | - | | - | - | 0.0353 | 0,1 | 0,1 | 0.785 | -0.112 | - | - | - | - | - |
| rs112804604 | 22:29936137-29936138 | - | intron_variant | MODIFIER | *MTMR3* | Transcript | NM_021090.4 | protein_coding | 1/19 | NM_021090.4:c.-137-20898del | rs112804604 | - | 1 | 0.0353 | - | - | 3.772 | 0.2558 | 0.00 | 0.00 | 0.00 | 0.00 | *MTMR3* |
| | | - | intron_variant | | *MTMR3* | Transcript | NM_153050.3 | protein_coding | 1/19 | NM_153050.3:c.-137-20898del | | - | 1 | 0.0353 | - | - | 3.772 | 0.2558 | 0.00 | 0.00 | 0.00 | 0.00 | *MTMR3* |
| | | - | intron_variant | | *MTMR3* | Transcript | NM_153051.3 | protein_coding | 1/18 | NM_153051.3:c.-137-20898del | | - | 1 | 0.0353 | - | - | 3.772 | 0.2558 | 0.00 | 0.00 | 0.00 | 0.00 | *MTMR3* |
| | | - | regulatory_region_variant | | - | RegulatoryFeature | ENSR00000669734 | enhancer | - | - | | - | - | 0.0353 | - | - | 3.772 | 0.2558 | - | - | - | - | - |
| rs111908292 | 22:29939914-29939914 | G | intron_variant | MODIFIER | *MTMR3* | Transcript | NM_021090.4 | protein_coding | 1/19 | NM_021090.4:c.-137-17122A>G | rs111908292 | - | 1 | 0.0353 | - | - | 3.052 | 0.1937 | 0.00 | 0.00 | 0.00 | 0.00 | *MTMR3* |
| | | G | intron_variant | | *MTMR3* | Transcript | NM_153050.3 | protein_coding | 1/19 | NM_153050.3:c.-137-17122A>G | | - | 1 | 0.0353 | - | - | 3.052 | 0.1937 | 0.00 | 0.00 | 0.00 | 0.00 | *MTMR3* |
| | | G | intron_variant | | *MTMR3* | Transcript | NM_153051.3 | protein_coding | 1/18 | NM_153051.3:c.-137-17122A>G | | - | 1 | 0.0353 | - | - | 3.052 | 0.1937 | 0.00 | 0.00 | 0.00 | 0.00 | *MTMR3* |
| rs113140876 | 22:29940180-29940180 | G | intron_variant | MODIFIER | *MTMR3* | Transcript | NM_021090.4 | protein_coding | 1/19 | NM_021090.4:c.-137-16856A>G | rs113140876 | - | 1 | 0.0353 | - | - | 4.351 | 0.3054 | 0.00 | 0.00 | 0.00 | 0.00 | *MTMR3* |
| | | G | intron_variant | | *MTMR3* | Transcript | NM_153050.3 | protein_coding | 1/19 | NM_153050.3:c.-137-16856A>G | | - | 1 | 0.0353 | - | - | 4.351 | 0.3054 | 0.00 | 0.00 | 0.00 | 0.00 | *MTMR3* |
| | | G | intron_variant | | *MTMR3* | Transcript | NM_153051.3 | protein_coding | 1/18 | NM_153051.3:c.-137-16856A>G | | - | 1 | 0.0353 | - | - | 4.351 | 0.3054 | 0.00 | 0.00 | 0.00 | 0.00 | *MTMR3* |
| rs73398629 | 22:29949551-29949551 | C | intron_variant | MODIFIER | *MTMR3* | Transcript | NM_021090.4 | protein_coding | 1/19 | NM_021090.4:c.-137-7485A>C | rs73398629 | - | 1 | 0.0353 | - | - | 4.695 | 0.3352 | 0.00 | 0.00 | 0.00 | 0.00 | *MTMR3* |
| | | C | intron_variant | | *MTMR3* | Transcript | NM_153050.3 | protein_coding | 1/19 | NM_153050.3:c.-137-7485A>C | | - | 1 | 0.0353 | - | - | 4.695 | 0.3352 | 0.00 | 0.00 | 0.00 | 0.00 | *MTMR3* |
| | | C | intron_variant | | *MTMR3* | Transcript | NM_153051.3 | protein_coding | 1/18 | NM_153051.3:c.-137-7485A>C | | - | 1 | 0.0353 | - | - | 4.695 | 0.3352 | 0.00 | 0.00 | 0.00 | 0.00 | *MTMR3* |
| | | C | regulatory_region_variant | | - | RegulatoryFeature | ENSR00001059251 | enhancer | - | - | | - | - | 0.0353 | - | - | 4.695 | 0.3352 | - | - | - | - | - |
| rs73398630 | 22:29949778-29949778 | A | intron_variant | MODIFIER | *MTMR3* | Transcript | NM_021090.4 | protein_coding | 1/19 | NM_021090.4:c.-137-7258G>A | rs73398630 | - | 1 | 0.0355 | - | - | 0.685 | -0.14 | 0.00 | 0.00 | 0.00 | 0.00 | *MTMR3* |
| | | A | intron_variant | | *MTMR3* | Transcript | NM_153050.3 | protein_coding | 1/19 | NM_153050.3:c.-137-7258G>A | | - | 1 | 0.0355 | - | - | 0.685 | -0.14 | 0.00 | 0.00 | 0.00 | 0.00 | *MTMR3* |
| | | A | intron_variant | | *MTMR3* | Transcript | NM_153051.3 | protein_coding | 1/18 | NM_153051.3:c.-137-7258G>A | | - | 1 | 0.0355 | - | - | 0.685 | -0.14 | 0.00 | 0.00 | 0.00 | 0.00 | *MTMR3* |
| | | A | regulatory_region_variant | | - | RegulatoryFeature | ENSR00001059251 | enhancer | - | - | | - | - | 0.0355 | - | - | 0.685 | -0.14 | - | - | - | - | - |
| rs60093195 | 22:29958748-29958748 | G | intron_variant | MODIFIER | *MTMR3* | Transcript | NM_021090.4 | protein_coding | 2/19 | NM_021090.4:c.-85+1660A>G | rs60093195 | - | 1 | 0.0353 | - | - | 0.783 | -0.112 | 0.00 | 0.00 | 0.00 | 0.00 | *MTMR3* |
| | | G | intron_variant | | *MTMR3* | Transcript | NM_153050.3 | protein_coding | 2/19 | NM_153050.3:c.-85+1660A>G | | - | 1 | 0.0353 | - | - | 0.783 | -0.112 | 0.00 | 0.00 | 0.00 | 0.00 | *MTMR3* |
| | | G | intron_variant | | *MTMR3* | Transcript | NM_153051.3 | protein_coding | 2/18 | NM_153051.3:c.-85+1660A>G | | - | 1 | 0.0353 | - | - | 0.783 | -0.112 | 0.00 | 0.00 | 0.00 | 0.00 | *MTMR3* |
| | 22:29962609[illegible] | T | intron_variant | | *MTMR3* | Transcript | NM_021090.4 | protein_coding | 2/19 | NM_021090.4:c.-85+5521C>T | | - | 1 | 0.0353 | - | - | 0.64 | -0.154 | 0.00 | 0.00 | 0.00 | 0.00 | *MTMR3* |

| | | | | | | | | | | | | | | | | | | | | | | | |
|---|---|---|---|---|---|---|---|---|---|---|---|---|---|---|---|---|---|---|---|---|---|---|---|
| rs58564057 | 22:29962609-29962609 | T | intron_variant | MODIFIER | *MTMR3* | Transcript | NM_153050.3 | protein_coding | 2/19 | NM_153050.3:c.-85+5521C>T | rs58564057 | - | 1 | 0.0353 | - | - | 0.64 | -0.154 | 0.00 | 0.00 | 0.00 | 0.00 | *MTMR3* |
| | | T | intron_variant | | *MTMR3* | Transcript | NM_153051.3 | protein_coding | 2/18 | NM_153051.3:c.-85+5521C>T | | - | 1 | 0.0353 | - | - | 0.64 | -0.154 | 0.00 | 0.00 | 0.00 | 0.00 | *MTMR3* |
| rs73398654 | 22:29980789-29980789 | G | intron_variant | MODIFIER | *MTMR3* | Transcript | NM_021090.4 | protein_coding | 5/19 | NM_021090.4:c.210+1737C>G | rs73398654 | - | 1 | 0.0353 | - | - | 0.248 | -0.347 | 0.00 | 0.00 | 0.00 | 0.00 | *MTMR3* |
| | | G | intron_variant | | *MTMR3* | Transcript | NM_153050.3 | protein_coding | 5/19 | NM_153050.3:c.210+1737C>G | | - | 1 | 0.0353 | - | - | 0.248 | -0.347 | 0.00 | 0.00 | 0.00 | 0.00 | *MTMR3* |
| | | G | intron_variant | | *MTMR3* | Transcript | NM_153051.3 | protein_coding | 5/18 | NM_153051.3:c.210+1737C>G | | - | 1 | 0.0353 | - | - | 0.248 | -0.347 | 0.00 | 0.00 | 0.00 | 0.00 | *MTMR3* |
| rs73398659 | 22:29986440-29986440 | A | intron_variant | MODIFIER | *MTMR3* | Transcript | NM_021090.4 | protein_coding | 5/19 | NM_021090.4:c.211-2040G>A | rs73398659 | - | 1 | 0.0353 | - | - | 0.062 | -0.631 | 0.00 | 0.00 | 0.00 | 0.00 | *MTMR3* |
| | | A | intron_variant | | *MTMR3* | Transcript | NM_153050.3 | protein_coding | 5/19 | NM_153050.3:c.211-2040G>A | | - | 1 | 0.0353 | - | - | 0.062 | -0.631 | 0.00 | 0.00 | 0.00 | 0.00 | *MTMR3* |
| | | A | intron_variant | | *MTMR3* | Transcript | NM_153051.3 | protein_coding | 5/18 | NM_153051.3:c.211-2040G>A | | - | 1 | 0.0353 | - | - | 0.062 | -0.631 | 0.00 | 0.00 | 0.00 | 0.00 | *MTMR3* |
| | | A | regulatory_region_variant | | - | RegulatoryFeature | ENSR00000669742 | enhancer | - | - | | - | - | 0.0353 | - | - | 0.062 | -0.631 | - | - | - | - | - |
| rs73398662 | 22:29987780-29987780 | G | intron_variant | MODIFIER | *MTMR3* | Transcript | NM_021090.4 | protein_coding | 5/19 | NM_021090.4:c.211-700C>G | rs73398662 | - | 1 | 0.0353 | - | - | 1.504 | 0.0256 | 0.00 | 0.00 | 0.00 | 0.00 | *MTMR3* |
| | | G | intron_variant | | *MTMR3* | Transcript | NM_153050.3 | protein_coding | 5/19 | NM_153050.3:c.211-700C>G | | - | 1 | 0.0353 | - | - | 1.504 | 0.0256 | 0.00 | 0.00 | 0.00 | 0.00 | *MTMR3* |
| | | G | intron_variant | | *MTMR3* | Transcript | NM_153051.3 | protein_coding | 5/18 | NM_153051.3:c.211-700C>G | | - | 1 | 0.0353 | - | - | 1.504 | 0.0256 | 0.00 | 0.00 | 0.00 | 0.00 | *MTMR3* |
| | | G | regulatory_region_variant | | - | RegulatoryFeature | ENSR00000669742 | enhancer | - | - | | - | - | 0.0353 | - | - | 1.504 | 0.0256 | - | - | - | - | - |
| rs73398664 | 22:29989196-29989196 | T | intron_variant | MODIFIER | *MTMR3* | Transcript | NM_021090.4 | protein_coding | 6/19 | NM_021090.4:c.293+634G>T | rs73398664 | - | 1 | 0.0355 | - | - | 3.82 | 0.2599 | 0.00 | 0.00 | 0.00 | 0.00 | *MTMR3* |
| | | T | intron_variant | | *MTMR3* | Transcript | NM_153050.3 | protein_coding | 6/19 | NM_153050.3:c.293+634G>T | | - | 1 | 0.0355 | - | - | 3.82 | 0.2599 | 0.00 | 0.00 | 0.00 | 0.00 | *MTMR3* |
| | | T | intron_variant | | *MTMR3* | Transcript | NM_153051.3 | protein_coding | 6/18 | NM_153051.3:c.293+634G>T | | - | 1 | 0.0355 | - | - | 3.82 | 0.2599 | 0.00 | 0.00 | 0.00 | 0.00 | *MTMR3* |
| | | T | regulatory_region_variant | | - | RegulatoryFeature | ENSR00001238990 | enhancer | - | - | | - | - | 0.0355 | - | - | 3.82 | 0.2599 | - | - | - | - | - |
| rs112672347 | 22:30004075-30004076 | A | intron_variant | MODIFIER | *MTMR3* | Transcript | NM_021090.4 | protein_coding | 9/19 | NM_021090.4:c.671+1083del | rs112672347 | - | 1 | 0.0353 | - | - | 6.746 | 0.5233 | 0.00 | 0.00 | 0.00 | 0.00 | *MTMR3* |
| | | A | intron_variant | | *MTMR3* | Transcript | NM_153050.3 | protein_coding | 9/19 | NM_153050.3:c.671+1083del | | - | 1 | 0.0353 | - | - | 6.746 | 0.5233 | 0.00 | 0.00 | 0.00 | 0.00 | *MTMR3* |
| | | A | intron_variant | | *MTMR3* | Transcript | NM_153051.3 | protein_coding | 9/18 | NM_153051.3:c.671+1083del | | - | 1 | 0.0353 | - | - | 6.746 | 0.5233 | 0.00 | 0.00 | 0.00 | 0.00 | *MTMR3* |
| | | A | upstream_gene_variant | | *MIR6818* | Transcript | NR_106876.1 | miRNA | - | - | | 2973 | 1 | 0.0353 | - | - | 6.746 | 0.5233 | - | - | - | - | - |
| | | A | downstream_gene_variant | | *HORMAD2-AS1* | Transcript | NR_110541.2 | lncRNA | - | - | | 4670 | -1 | 0.0353 | - | - | 6.746 | 0.5233 | - | - | - | - | - |
| rs73398683 | 22:30004230-30004230 | A | intron_variant | MODIFIER | *MTMR3* | Transcript | NM_021090.4 | protein_coding | 9/19 | NM_021090.4:c.671+1237G>A | rs73398683 | - | 1 | 0.0353 | - | - | 9.222 | 0.7828 | 0.00 | 0.00 | 0.02 | 0.00 | *MTMR3* |
| | | A | intron_variant | | *MTMR3* | Transcript | NM_153050.3 | protein_coding | 9/19 | NM_153050.3:c.671+1237G>A | | - | 1 | 0.0353 | - | - | 9.222 | 0.7828 | 0.00 | 0.00 | 0.02 | 0.00 | *MTMR3* |
| | | A | intron_variant | | *MTMR3* | Transcript | NM_153051.3 | protein_coding | 9/18 | NM_153051.3:c.671+1237G>A | | - | 1 | 0.0353 | - | - | 9.222 | 0.7828 | 0.00 | 0.00 | 0.02 | 0.00 | *MTMR3* |
| | | A | upstream_gene_variant | | *MIR6818* | Transcript | NR_106876.1 | miRNA | - | - | | 2819 | 1 | 0.0353 | - | - | 9.222 | 0.7828 | - | - | - | - | - |
| | | A | downstream_gene_variant | | *HORMAD2-AS1* | Transcript | NR_110541.2 | lncRNA | - | - | | 4516 | -1 | 0.0353 | - | - | 9.222 | 0.7828 | - | - | - | - | - |

**Supplementary Data 9. Summary of per locus association results after conditioning on top index variant**

| CHR | POS (hg38) | rsID | REF | ALT | ALT Freq | Nearest Gene | Pre-conditioning | | | N SNPs in region[a] | Post-conditioning | | | Signif.[b] |
|---|---|---|---|---|---|---|---|---|---|---|---|---|---|---|
| | | | | | | | Beta | SE | *P*-value | | Beta | SE | *P*-value | |
| 1 | 178057912 | rs111238523 | C | T | 2% | *RASAL2* | 0.058 | 0.018 | 1.76E-03 | 5008 | 0.065 | 0.018 | 4.30E-04 | |
| **2** | **422144** | **rs62107261** | **T** | **C** | **3%** | ***ALKAL2*** | **-0.095** | **0.014** | **3.83E-12** | **6640** | **-0.097** | **0.014** | **2.06E-12** | **Yes** |
| 2 | 24881806 | n/a | CCA | C | 11% | *ADCY3* | 0.042 | 0.009 | 2.94E-06 | 4187 | 0.038 | 0.009 | 2.63E-05 | |
| 3 | 185800743 | rs73061097 | T | A | 32% | *IGF2BP2* | -0.024 | 0.005 | 5.70E-06 | 4848 | -0.024 | 0.005 | 3.47E-06 | |
| 4 | 45308738 | rs80129601 | A | G | 2% | *GNPDA2* | -0.087 | 0.019 | 4.24E-06 | 6131 | -0.081 | 0.019 | 2.06E-05 | |
| 5 | 75750228 | rs258502 | G | T | 57% | *POC5* | -0.001 | 0.005 | 8.27E-01 | 6179 | -0.019 | 0.005 | 6.14E-04 | |
| 6 | 51174169 | rs984697642 | G | A | 3% | *TFAP2B* | -0.072 | 0.014 | 3.07E-07 | 5184 | -0.065 | 0.014 | 3.97E-06 | |
| 8 | 75725530 | rs16939149 | A | G | 6% | *HNF4G* | -0.039 | 0.011 | 4.29E-04 | 5692 | -0.051 | 0.011 | 8.10E-06 | |
| 11 | 27647068 | rs3838785 | A | G | 2% | *BDNF* | 0.071 | 0.020 | 3.06E-04 | 3667 | 0.080 | 0.020 | 5.76E-05 | |
| 12 | 49831467 | rs4391887 | A | G | 74% | *NCKAP5L* | 0.034 | 0.006 | 1.18E-09 | 4264 | 0.023 | 0.006 | 2.79E-04 | |
| 13 | 53367083 | rs1036958 | T | A | 52% | *OLFM4* | -0.014 | 0.005 | 4.02E-03 | 5125 | -0.018 | 0.005 | 2.70E-04 | |
| 16 | 53730708 | rs117502563 | G | A | 4% | *FTO* | 0.031 | 0.012 | 7.20E-03 | 5673 | 0.054 | 0.012 | 4.30E-06 | |
| **18** | **60361739** | **rs78769612** | **G** | **T** | **2%** | ***MC4R*** | **-0.106** | **0.019** | **3.53E-08** | **6861** | **-0.100** | **0.019** | **2.17E-07** | **Yes** |
| 19 | 47321406 | rs149173729 | C | T | 1% | *C5AR1* | 0.094 | 0.027 | 4.64E-04 | 4744 | 0.108 | 0.027 | 6.65E-05 | |
| 22 | 30035665 | rs73400621 | A | G | 5% | *MTMR3* | 0.029 | 0.012 | 1.23E-02 | 4814 | 0.042 | 0.012 | 4.80E-04 | |
| X | 32268401 | rs147568648 | A | G | 11% | *DMD* | -0.023 | 0.007 | 6.50E-04 | 4911 | -0.025 | 0.007 | 2.19E-04 | |

*CHR, chromosome; POS, position; REF, reference allele; ALT, alternative allele; Freq, frequency; SE, standard error; Signif., significant; n/a, not available*

[a] Region was defined as within ± 500kb of each index variant.

[b] Significance threshold for secondary signals ($P < 5.96 \times 10^{-7}$) was determined by Bonferroni correction for the number of variants across all regions

**Supplementary Data 10. Summary of association results after conditioning on all known index variants**

| CHR | POS (hg38) | rsID | Nearest Gene | REF | ALT | ALT Freq | Pre-conditioning | | | Post-conditioning | | |
|---|---|---|---|---|---|---|---|---|---|---|---|---|
| | | | | | | | Beta | SE | *P*-value | Beta | SE | *P*-value |
| 2 | 621558 | rs939584 | *TMEM18* | C | T | 85% | 0.0576 | 0.0068 | 1.99E-17 | 0.0392 | 0.0282 | 0.1655 |
| 3 | 186108951 | rs869400 | *ETV5* | T | G | 82% | 0.0383 | 0.0063 | 1.21E-09 | 0.0330 | 0.0225 | 0.1439 |
| **6** | **50830813** | **rs2206277** | ***TFAP2B*** | **C** | **T** | **19%** | **0.0543** | **0.0062** | **2.05E-18** | **0.0687** | **0.0182** | **1.59E-04** |
| 8 | 76068626 | rs830463 | *HNF4G* | A | G | 47% | 0.0305 | 0.0049 | 6.58E-10 | 0.0237 | 0.0058 | 4.31E-05 |
| **11** | **27657463** | **rs3838785** | ***BDNF*** | **GT** | **G** | **58%** | **-0.0303** | **0.0051** | **3.14E-09** | **-0.0324** | **0.0092** | **4.11E-04** |
| 13 | 53533448 | rs9568868 | *OLFM4* | G | T | 14% | 0.0472 | 0.0072 | 5.73E-11 | 0.1990 | 0.1881 | 0.2900 |
| 19 | 47077985 | rs28590228 | *ZC3H4* | C | T | 50% | 0.0332 | 0.0053 | 4.75E-10 | 0.0410 | 0.0219 | 0.0617 |
| **Secondary signal** | | | | | | | | | | | | |
| 18 | 60361739 | rs78769612 | *MC4R* | G | T | 2% | -0.106 | 0.019 | 3.53E-08 | -0.0722 | 0.0945 | 0.4450 |

*CHR, chromosome; POS, position; REF, reference allele; ALT, alternative allele; Freq, frequency; SE, standard error*

Known index variants curated from the following papers (PMIDs): 22344219, 22344221, 23563607, 23583978, 24094743, 24861553, 25673413, 26426971, 28391526, 28430825, 28443625, 28448500, 28552196, 28892062, 29273807, 29381148, 30108127, 30124842, 30595370, 31217584, 35399580

**Supplementary Data 11. PAINTOR results for top loci assuming one single causal variant at each locus**

Only index variants and those with posterior probability (PP )> 50% are shown. **Bold** identifies variants with > 95% PP.

| Locus rsID | CHR | POS (hg38) | rsID | REF | ALT | EAF | MAF | Gene | PAINTOR Annotation | Beta | SE | *P*-value | PVE (%) | PP | Is Index highest PP? |
|---|---|---|---|---|---|---|---|---|---|---|---|---|---|---|---|
| rs543874 | 1 | 177920345 | rs543874 | A | G | 0.2043 | 0.2043 | *SEC16B* | - | 0.0639 | 0.0060 | 1.38E-26 | 0.128 | 0.6192 | Yes |
| rs939584 | 2 | 621558 | rs939584 | C | T | 0.8518 | 0.1482 | *TMEM18* | - | 0.0576 | 0.0068 | 1.99E-17 | 0.081 | 0.0230 | Yes |
| rs10182181 | 2 | 24927427 | rs10182181 | A | G | 0.5608 | 0.4392 | *ADCY3* | - | 0.0350 | 0.0052 | 1.76E-11 | 0.051 | 0.1376 | No |
| | | 24920780 | rs6746013 | C | G | 0.5629 | 0.4371 | | genehancer | 0.3462 | 0.0052 | 2.99E-11 | 0.050 | 0.3163 | |
| rs869400 | 3 | 186108951 | rs869400 | T | G | 0.8195 | 0.1805 | *ETV5* | genehancer | 0.0383 | 0.0063 | 1.21E-09 | 0.042 | 0.6263 | Yes |
| rs12507026 | 4 | 45179317 | rs12507026 | A | T | 0.3614 | 0.3614 | *GNPDA2* | - | 0.0449 | 0.0051 | 9.55E-19 | 0.088 | 0.3051 | Yes |
| **rs2307111** | **5** | **75707853** | **rs2307111** | **T** | **C** | **0.5474** | **0.4526** | ***POC5*** | **-** | **-0.0324** | **0.0053** | **7.43E-10** | **0.043** | **0.9898** | **Yes** |
| rs2206277 | 6 | 50830813 | rs2206277 | C | T | 0.1903 | 0.1903 | *TFAP2B* | - | 0.0543 | 0.0062 | 2.05E-18 | 0.086 | 0.4895 | Yes |
| rs830463 | 8 | 76068626 | rs830463 | A | G | 0.4700 | 0.4700 | *HNF4G* | - | 0.0305 | 0.0049 | 6.58E-10 | 0.043 | 0.2278 | Yes |
| rs3838785 | 11 | 27657463 | rs3838785 | GT | G | 0.5792 | 0.4208 | *BDNF* | - | -0.0303 | 0.0051 | 3.14E-09 | 0.040 | 0.3310 | Yes |
| rs7138803 | 12 | 49853685 | rs7138803 | G | A | 0.2974 | 0.2974 | *BCDIN3D* | - | 0.0363 | 0.0054 | 1.69E-11 | 0.051 | 0.4231 | Yes |
| rs9568868 | 13 | 53533448 | rs9568868 | G | T | 0.1387 | 0.1387 | *OLFM4* | - | 0.0472 | 0.0072 | 5.74E-11 | 0.048 | 0.3631 | Yes |
| rs1421085 | 16 | 53767042 | rs1421085 | T | C | 0.2949 | 0.2949 | *FTO* | - | 0.0901 | 0.0056 | 6.11E-59 | 0.295 | 0.5887 | Yes |
| rs6567160 | 18 | 60161902 | rs6567160 | T | C | 0.2096 | 0.2096 | *MC4R* | - | 0.0525 | 0.0059 | 8.22E-19 | 0.088 | 0.7360 | Yes |
| rs28590228 | 19 | 47077985 | rs28590228 | C | T | 0.5039 | 0.4961 | *ZC3H4* | - | 0.0332 | 0.0053 | 4.76E-10 | 0.044 | 0.0945 | No |
| | | 47113120 | rs55731973 | G | T | 0.5196 | 0.4804 | | promoter, genehancer | 0.0297 | 0.0054 | 3.91E-08 | 0.034 | 0.7664 | |
| rs111490516 | 22 | 29906934 | rs111490516 | C | T | 0.0368 | 0.0368 | *MTMR3* | - | 0.0783 | 0.0133 | 4.52E-09 | 0.039 | 0.0407 | Yes |
| **rs1379871** | **X** | **31836665** | **rs1379871** | **G** | **C** | **0.4131** | **0.4131** | ***DMD*** | **-** | **0.0287** | **0.0042** | **1.35E-11** | **0.052** | **0.9980** | **Yes** |

*PAINTOR, Probabilistic Annotation INtegraTOR; CHR, chromosome; POS, position; REF, reference allele; ALT, alternative allele; EAF, effect allele frequency; MAF, minor allele frequency; SE, standard error; PVE, percent variance explained; PP, posterior probability*

**Supplementary Data 12. Top associations meeting suggestive significance ( *P* < 0.001) in PheWAS meta-analysis**

| | | | Meta | | | | | MyCode | | | | | BioMe | | | | |
|---|---|---|---|---|---|---|---|---|---|---|---|---|---|---|---|---|---|
| **PheCode** | **Phenotype Descriptions** | **Category** | **Beta** | **SE** | **OR** | ***P*** | **N** | **Beta** | **SE** | **OR** | ***P*** | **N** | **Beta** | **SE** | **OR** | ***P*** | **N** |
| 327.3 | Sleep apnea | neurological | -0.2530 | 0.0755 | 0.7765 | 8.12E-04 | 7,600 | -0.1436 | 0.1391 | 0.8663 | 3.02E-01 | 1,536 | -0.2987 | 0.0900 | 0.7418 | 8.99E-04 | 6,064 |
| 327.32 | Obstructive sleep apnea | neurological | -0.2903 | 0.0785 | 0.7480 | 2.19E-04 | 7,497 | -0.1809 | 0.1455 | 0.8345 | 2.14E-01 | 1,486 | -0.3352 | 0.0933 | 0.7152 | 3.26E-04 | 6,011 |
